# HYPER: Group testing via hypergraph factorization applied to COVID-19

**DOI:** 10.1101/2021.02.24.21252394

**Authors:** David Hong, Rounak Dey, Xihong Lin, Brian Cleary, Edgar Dobriban

**Affiliations:** Department of Statistics, The Wharton School, University of Pennsylvania, Philadelphia, PA, USA; Department of Biostatistics, Harvard T.H. Chan School of Public Health, Boston, MA, USA; Department of Statistics, Harvard University, Cambridge, MA, USA; Broad Institute of MIT and Harvard, Cambridge, MA 02142 USA

## Abstract

Large scale screening is a critical tool in the life sciences, but is often limited by reagents, samples, or cost. An important challenge in screening has recently manifested in the ongoing effort to achieve widespread testing for individuals with SARS-CoV-2 infection in the face of substantial resource constraints. Group testing methods utilize constrained testing resources more efficiently by pooling specimens together, potentially allowing larger populations to be screened with fewer tests. A key challenge in group testing is to design an effective pooling strategy. The global nature of the ongoing pandemic calls for something simple (to aid implementation) and flexible (to tailor for settings with differing needs) that remains efficient. Here we propose HYPER, a new group testing method based on hypergraph factorizations. We provide theoretical characterizations under a general statistical model, and exhaustively evaluate HYPER and proposed alternatives for SARS-CoV-2 screening under realistic simulations of epidemic spread and within-host viral kinetics. We demonstrate that HYPER performs at least as well as other methods in scenarios that are well-suited to each method, while outperforming those methods across a broad range of resource-constrained environments, being more flexible and simple in design, and taking no expertise to implement. An online tool to implement these designs in the lab is available at http://hyper.covid19-analysis.org.

## Introduction

Biological screens that identify members of a large population with a disease have become invaluable tools for disease diagnosis and surveillance. When these screens are difficult to conduct or resources are limited, finding an efficient way to conduct the screen becomes critical. As such, widespread, scalable and frequent testing is a defining challenge in combatting COVID-19 in the face of local, national and global resource constraints. Pooled testing has recently arisen as a promising efficient scientific solution to the world-wide challenge of increasing SARS-CoV-2 testing capacity^1–17^, encouraged in part by the finding that a single positive sample can be reliably detected by RT-qPCR in large pools^18^.

One approach to achieve efficiency is to design the screen in such a way that leverages structure in the tested population^19^. This idea dates back at least to the seminal work of Dorfman^20^, which proposed testing *pools* of samples when there is prior knowledge that the vast majority of samples will test negative. Dorfman testing is a two-stage approach with each individual assigned to exactly one pool. A negative test result for a pool at the first stage can be applied to all its members, eliminating the need to test them individually and potentially greatly increasing efficiency, depending on the pool size and prevalence of positive members of the population. A great strength of this approach is its simplicity and robustness in laboratory implementation. Pools are easy to form and putative positives are simply those individuals contained in a positive pool. Several early methods^4–7^ for COVID-19 pooled testing focus on Dorfman testing. However, it is well-known that Dorfman testing can have sub-optimal efficiency^9–12^; alternative designs use tests more efficiently and can thus screen more individuals, especially in the face of major resource constraints.

There has been tremendous study and progress on pooled testing (also called group testing or specimen pooling) in general. Numerous works provide statistical^21–25^, combinatorial^26–31^, as well as information and coding theoretic^32–45^ perspectives, as well as software^46;47^ to aid implementation, to name just a few. Additionally, there has been a lot of work on analyzing and optimizing group testing for various constraints and evaluation criteria^48–59^, often in the low prevalence regime. Broadly speaking, the approaches fall into three categories: i) single-stage (or nonadaptive) approaches that identify positive individuals after only one round of pooled tests by using pools with carefully designed overlaps; ii) two-stage^58–61^ approaches that declare putative positives after one round of pooled tests. The putative positives are then individually tested in the second round. Finally, iii) multi-stage^61;62^ (or adaptive/hierarchical) approaches that carry out multiple rounds of pooled tests with pools at each round chosen based on previous rounds.

Many recent methods^8–10;12–16;63^ for COVID-19 pooled testing (more than can be reviewed here) are seeking greater efficiency by splitting samples into multiple pools, and using a one-, two- or multi-stage approach. One such method is P-BEST^8^, which is a single-stage approach designed for a prevalence around 1% that splits each of 384 individuals into 48 partially overlapping pools. The pool assignments are based on a Reed-Solomon error correcting code that enables identification from the single round of tests and provides robustness against, e.g., independent PCR failures. Positive individuals are identified by running a specialized decoding algorithm based on sparse regression. A popular two-stage method is plate-based array pooling^9^, which arranges individuals into either an 8 × 12 or 16 × 24 grid (corresponding to plate sizes common in laboratory environments), then takes each column and each row to be a pool, resulting in 20 pools for 96 individuals or 40 pools for 384 individuals, respectively. Each individual is split into two pools and is a putative positive only if both those pools test positive. This approach retains some of the simplicity of Dorfman testing while being potentially more efficient. For example, negative individuals in positive pools can be identified if their other pool tests negative, avoiding the need to test them individually, whereas they would all be putative positives in Dorfman testing. Moreover, these array designs are well-suited for clinics that already store samples in these grids, especially if they have multi-channel pipettes that make row/column pooling convenient.

The hypercube method^10^ extends this idea by arranging individuals in a *q*-dimensional grid with sides of length three. The rows and columns used to form pools in the array design are generalized to multidimensional “slices” each containing 3^*q*−1^ individuals. In this design, each individual belongs to *q* of the 3*q* total pools, affording the same opportunity for efficiency gains by identifying negative individuals in positive pools by using their other pools. Moreover, hypercube is an adaptive multi-stage method. Results from each round of pooled testing are used to design further hypercube pools to test (see their paper^10^ for details), giving further opportunities for potential efficiency gains. Together, these various proposed pooling strategies offer several alternatives that address the urgent, global need for efficient screening.

However, given the global nature of the pandemic, there is a wide variety of settings with differing needs and constraints, in which the proposed combinatorial designs may have limited utility^11^. Designs that split samples into more than two or three pools, e.g., P-BEST and hypercubes, can be time-consuming and error-prone to execute by hand, making them best-suited for well-resourced labs with robotic-pipetting platforms. The decoding algorithm used by P-BEST and the multi-stage logistics of hypercubes can also make them nontrivial to implement without prior experience, expertise, and substantial lab infrastructure. Multi-stage methods can also take longer to return results, making them less suitable for time-sensitive public health settings. Moreover, many of the proposed designs are somewhat rigid and are not trivially adapted to environments that can have widely varying numbers of individuals to test, available test kits, or prevalence of positive results. Plate-based array pooling with 150 individuals, for example, might be done using a partially-filled 16 × 24 grid or perhaps two 8 × 12 grids with one or both partially-filled, but it is unclear which to choose and whether either remains efficient under variable prevalence. For SARS-CoV-2 screening in resource-limited settings, adapting to these various conditions is critical to achieving the greatest effectiveness^11^. Therefore, for this application along with broader applications of pooled screening, there remains an outstanding need for a robust, simple, flexible strategy with performance that matches (or exceeds) the effectiveness of specialized strategies, and can be applied in diverse environments without special equipment or expertise.

We propose HYPER, a two-stage pooling strategy based on the combinatorics of hypergraph factorizations. In this approach, individuals are assigned to pools by cycling through a carefully ordered sequence, given by a combinatorial construction. While the underlying mathematics is sophisticated, the resulting pools are simple to implement by hand (individuals are split at most three ways), and putative positives can be easily identified with only pencil and paper. We also provide an online tool available at http://hyper.covid19-analysis.org to facilitate implementation. The design accommodates any number of individuals while maintaining robustness, balance and efficiency. We characterize its performance under a common statistical model, as well as through realistic simulations that model both viral kinetics and epidemic spread. Our findings demonstrate that HYPER outperforms both plate-based array and P-BEST designs, which are particular instances of general array pooling and code-based designs. We found that HYPER also outperformed designs from the broad classes of balanced array and Reed-Solomon Kautz-Singleton code-based designs, even in the scenarios where those methods excel in efficiency. For SARS-CoV-2 and beyond, HYPER represents a valuable addition to the growing toolbox for performing large-scale, pooled screens.

## Results

### The need for balanced designs that are simple and flexible

A pooling design is an assignment of each of *n* individuals (or more generally, samples) to one or several of *m* pools. We seek a simple and flexible pooling design that is balanced in the following natural ways (defined formally in Supplementary Methods):

i. All individuals are assigned to the same number *q* of pools; we focus on *q* ≤ 3 for practical implementation.
ii. The *m* pools are assigned as evenly as possible, i.e., the sizes of the pools are as close as possible to equal.
iii. The 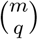 possible pool combinations are assigned as evenly as possible.

Similar but nonidentical balance conditions have been widely studied in group testing (see the references above). Here, the three conditions we require come from various naturally-arising real-life considerations that are especially relevant for COVID-19 testing. To begin, they all help make the pooling process more consistent, making robust implementation and quality control easier. For example, pool size determines how much volume to pipette from each individual in the pool. Balanced assignment of pools produces uniform pool sizes, so makes the volumes to pipette consistent across the design. Combined with uniform assignment of all individuals to *q* pools, this also makes the volume needed for all individuals consistent. Furthermore, a simple way to reduce pipetting steps in practice is to first pool individuals assigned to the same pool combination, then split this combined sample into the assigned pools. Balanced assignment of pool combinations makes the size of each of these combined samples consistent. Put together, these various forms of consistency simplify the pooling process and make it easier to add checks along the way, both of which help make it less error-prone.

Beyond making the pooling process more consistent, balance may also help make the performance of designs more consistent. For example, larger pools dilute positive samples more, which can increase the risk of false negatives. Balancing the assignment of pools can help make this reduction in sensitivity uniform across individuals. This is important for real-world testing; all individuals in the design should receive the same treatment. Similarly, balancing the assignment of pool combinations can help make efficiency more consistent by reducing the dependence of stage-two workload on which pool combinations become putative positives. This consistency in turn can help labs to better plan and efficiently allocate tests. We study how balance impacts the consistency of both sensitivity and efficiency in more detail under the COVID-19 model below.

For *q* = 1, corresponding to classical Dorfman testing, each individual needs to be assigned to a single pool. Balance can be easily achieved here by cycling through the pools until all individuals are assigned. For example, to assign *n* = 8 individuals to *m* = 6 pools (A-F), assign individual 1 to pool A, individual 2 to B, and so on, yielding the following assignments:

| Individual | 1 | 2 | 3 | 4 | 5 | 6 | 7 | 8 |
| --- | --- | --- | --- | --- | --- | --- | --- | --- |
| Pool | A | B | C | D | E | F | A | B |

All the *n* = 8 individuals are assigned to *q* = 1 pool, the *m* = 6 pools are assigned as evenly as possible (A and B are assigned twice and the rest are assigned once), and pool “combinations” are just the pools themselves when *q* = 1.

Creating maximally balanced designs for *q* > 1, and especially *q* > 2, can be much harder. A straightforward approach of listing the pools and assigning individuals to consecutive pairs (e.g., AB, CD, EF, AB, …) underutilizes the combinatorial space (for instance, it does not use AC). At the same time, cycling over all pairs sorted in lexicographic order is likely to be unbalanced in multiple ways. For instance, using all pairs in lexicographic order for *n* = 8 individuals assigned to *q* = 2 of *m* = 6 pools, yields

| Individual | 1 | 2 | 3 | 4 | 5 | 6 | 7 | 8 |
| --- | --- | --- | --- | --- | --- | --- | --- | --- |
| Pools | AB | AC | AD | AE | AF | BC | BD | BE |

which assigns A five times and F only once. As a result, this design is *not* uniformly sensitive across individuals. For example, individual 1 undergoes a five-fold dilution in pool A and a four-fold dilution in pool B, while individual 5 is diluted in pool A but not at all in pool F. Uneven dilution of viral loads can lead to differential false negative test results, leaving individual 1 with a higher risk of false negatives than individual 5. Clearly, alternative orderings could be more balanced, and the challenge is how to systematically identify these.

One approach is to generate a random design^11;64;65^. For example, one could assign each individual to *q* of *m* pools independently and uniformly at random^11^. Doing so treats both pools and pool combinations uniformly “on average”, but any given random draw may assign them very unevenly in practice, especially when *m, n, q* are not very large. For the example above, this approach draws one of the 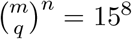 possible designs uniformly at random, so pool sizes (subjects per pool) can range all the way from zero to *n*. Likewise for pool combinations. In fact, 98.7% of the random draws in this case will not be maximally balanced, i.e., the pools or pool combinations are not assigned as evenly as possible, as can be verified by exhaustive search. Hence, it is rare for these randomly generated designs to both treat individuals uniformly, and maximally use all available pool combinations. With high probability, the random assignment design does not achieve our goal.

One could also consider searching among many random draws for a suitable design. In the small example above, it would take 180 draws to have a 90% chance of drawing at least one maximally balanced design. One might try selecting then modifying the most balanced (in some appropriate sense) from among the many draws, but such an approach is ad-hoc, involves manual tweaking, and may still not be maximally balanced. Double-pooling^64^ is an alternative random design that instead partitions the *n* individuals into *m/*2 pools two times. It generalizes to *q*-pooling by partitioning into *m/q* pools *q* times. This design produces balanced pools but does not guarantee balanced pool combinations, so again involves the possibility of manual tweaking.

An exhaustive search, on the other hand, is a systematic approach but is only practical for very small cases. The 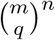 possible designs to search through is an enormous and intractable number in general. Moreover, one must repeat the process for any change in the numbers of individuals *n*, number of splits *q*, or number of pools *m*; the design is not easily adapted from one environment to another. One could attempt to precompute and store designs in advance, but this then limits us to the set of designs anticipated to be useful, again reducing flexibility. P-BEST^8^ attains balanced pools and pool combinations with a Reed-Solomon error-correcting design that requires technical expertise to adapt, and may only remain balanced in limited settings. Array designs^9^ can also be adapted to have balanced pools (by requiring the array to be a square), but they do not use all available pool combinations. They can have sub-optimal efficiency in some of our simulation settings.

As a result, though it is quite natural to desire maximally balanced designs, the problem of easily generating them is highly nontrivial. In fact, it is not initially clear that designs maximally balanced in all three ways even exist in general, let alone, an efficient way to find them.

Here we show how certain deep results from combinatorics, such as Baranyai’s theorem^66^ imply that such designs do in fact exist, under the minimal required conditions. Moreover, we demonstrate how to generate them under mild conditions using the combinatorics of hypergraph factorizations. For *q* = 3, we leverage a non-trivial number-theoretic construction due to Beth^67;68^. These tools enable us to develop a simple, flexible and efficient pooled testing strategy with maximally balanced pool designs.

### HYPER pooling method

We propose HYPER, a two-stage pooling strategy that uses maximally balanced pools built on hypergraph factorizations. Stage one consists of *pooled testing* to identify putative positives, who are tested individually in stage two. For convenience, we use H_*n,m,q*_ to denote the HYPER design with *n* individuals per batch, *m* pools, and *q* splits. We illustrate the details via a small example with *n* = 12 individuals each split into *q* = 2 out of *m* = 6 pools (Fig. 1a).

**Fig. 1.**
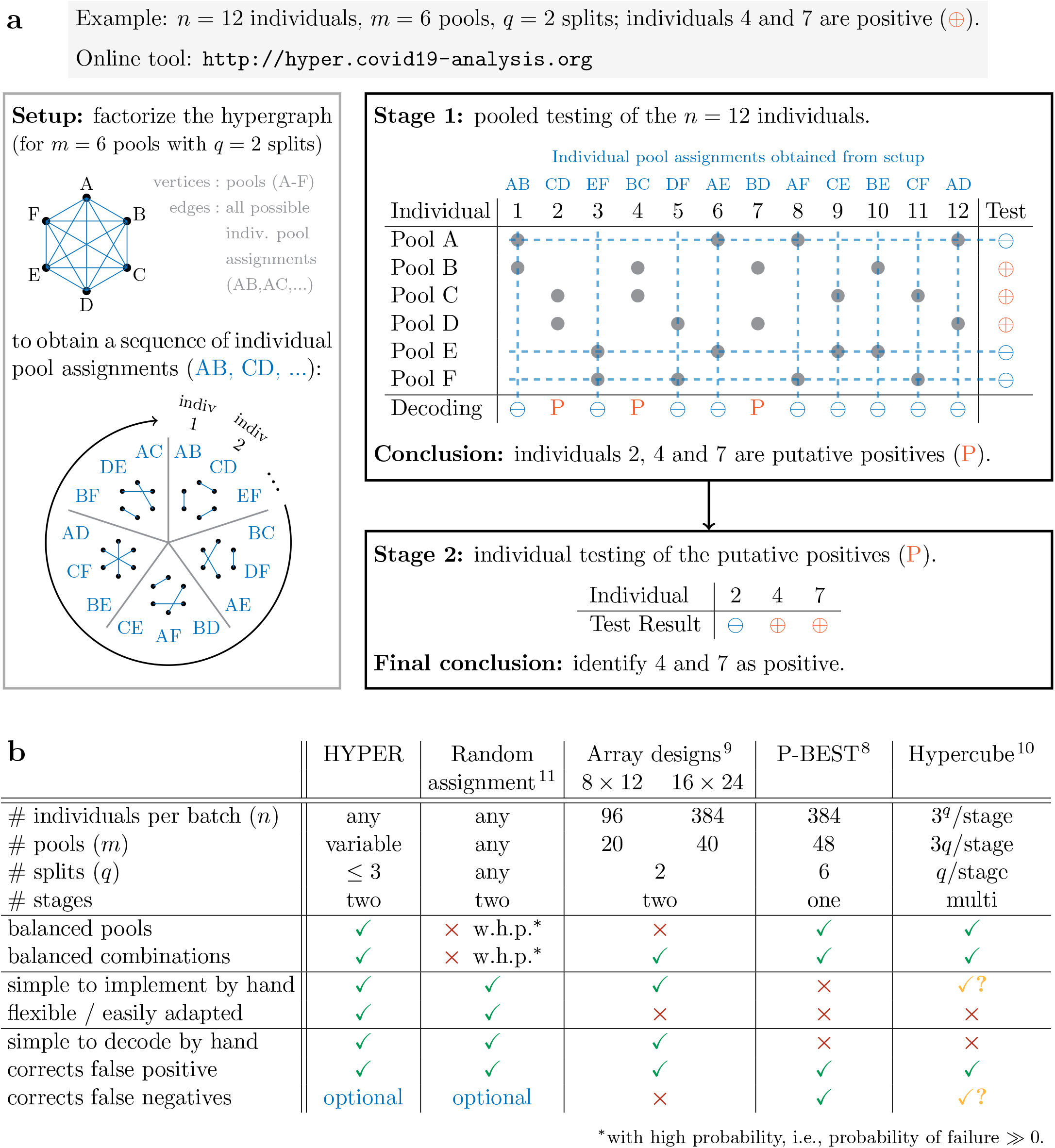
Overview of the HYPER method. **a** Illustrative example. Stage 1 tests pools that are formed by cycling through a sequence of pool assignments generated via hypergraph factorization. Putative positives are those individuals that are not in any negative pools (decoding). Stage 2 tests the putative positives individually. In this example, *n* = 12 individuals (2 of whom are actual positives) are each split into *q* = 2 of *m* = 6 pools; 3 are decoded as putative positives and both positives are successfully identified in stage 2. **b** Comparison of various features of HYPER with those of random designs^11^, plate-based array designs^9^, P-BEST^8^, and Hypercube^10^. HYPER is a maximally balanced design that is simple to implement/decode by hand, flexible and efficient.

First, to create pools for stage one, we assign each of the *n* individuals to *q* of the *m* pools by cycling through an ordering of the 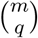 possible pool combinations generated via hypergraph factorization (as described below). If *q* = 1, this sequence of possible pool assignments would consist of the *m* pools in an arbitrary order. In our more involved example (Fig. 1a), the sequence reads AB, CD, etc., so individual 1 is assigned to pools A and B, individual 2 to C and D, and so on. In this case, we did not use all 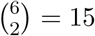 pairs in the sequence since there were only *n* = 12 individuals. However, if there were *n* > 15 individuals, we would cycle through the sequence again until all individuals were assigned. Hence, once we have the sequence, the pools are simple and flexible to form for any number of individuals *n*.

Next, we test each pool and *decode* the test results to identify putative positives. This is done by eliminating all the individuals that were in negative pools; all the individuals left over are the putative positives. Put another way, any individual that was only in positive pools is a putative positive. This is sometimes called *conservative* decoding^37^. It is easily illustrated and can be carried out with only pencil and paper, or a basic spreadsheet for larger designs (we also provide a web tool). In our example (Fig. 1a), pools A, E and F test negative so only individuals 2, 4 and 7 are putative positive. These samples are then individually tested in stage two.

Note that the conservative decoder we use (in the form described so far) does not correct for any pooled tests with false negative results, e.g., as is done in P-BEST^8^. Error correction may be incorporated by introducing a tolerance so that individuals in just one or two negative pools are still considered putative positives^11^. Doing so can improve the overall sensitivity of HYPER. However, an important source of false negatives is the dilution of viral loads below the limit of detection (which is a major concern). An individual with small viral load is hence likely to yield false negatives in most of its pools, in which case error-correction may not significantly improve sensitivity. Meanwhile, it can dramatically reduce efficiency. In our example (Fig. 1a), allowing a tolerance of one negative pool would turn individuals 1, 5, and 9-12 into putative positives. Labs must consider how to properly weigh these tradeoffs according to their specific needs. Here we will focus primarily on an error tolerance of zero, i.e., no error-correction of false negatives.

The only missing piece now is how we use hypergraph factorization to create the sequence of pool combinations for *q* = 2 and *q* = 3. Here we provide a brief overview, with further details of the construction provided in Supplementary Methods. Note that hypergraph factorization has been considered in group testing before^45^ but we apply it here in a different way. First, we require that *m* is divisible by *q*. Then, the idea is to order the 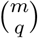 combinations so that there is no overlap among the combinations in each consecutive block of *m/q* combinations (which is in fact the formal definition of a “hypergraph factorization”, as we discuss below). In our example (Fig. 1a), the first *m/q* = 3 pairs (AB, CD, EF) do not overlap, as with the next three (BC, DF, AE), and so on. This property keeps pool sizes maximally balanced, and since each combination appears once, the combinations are balanced as well. Note that no pair is repeated and all pools contain 4 individuals. If we instead had only 11 individuals, the pools would no longer be perfectly even, since pools A and D would have only 3 individuals, but they would still be as even as possible. The same holds for any number of individuals.

Our task, therefore, is to divide the 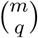 combinations into subsets of combinations, where each pool appears exactly once in each subset. As illustrated in Fig. 1a, we can think of each pool as one of *m* vertices in a *hypergraph*, where each *hyperedge* is a set of *q* of the *m* vertices (pools). Each hyperedge will correspond to a potential set of pools into which any individual sample is split. Our example has 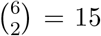 hyperedges (blue edges in Fig. 1a). Putting together all 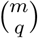 hyperedges forms the so-called complete hypergraph 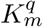 of order *q* on *m* vertices (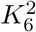 in Fig. 1a). Any subset of the hyperedges that uses all vertices exactly once is called a 1-*factor*. Restated in these terms, our task is to divide the 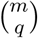 hyperedges of 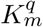 into disjoint 1-factors. This is known in combinatorics as *hypergraph factorization*. In our example (Fig. 1a), we show a factorization of 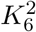 into five disjoint 1-factors, the first of which consists of hyperedges AB, CD, and EF. Hypergraph factorization has been the subject of intense study^28;29^. In particular, a deep result from combinatorics known as Baranyai’s theorem^66^ shows that factorizations *exist* under minimal conditions. Moreover, there are also efficient algorithms to construct them up to *q* = 3 under mild conditions^67–69^. More generally, and beyond what we need, the efficient construction of hypergraph factorizations is a challenging combinatorial problem.

The mathematical construction of hypergraph factorizations is somewhat sophisticated. However, the output is easy to use. We provide an online tool available at http://hyper.covid19-analysis.org that implements constructions for *q* = 2 (as long as *m* is even) and for *q* = 3 (as long as *m* is a multiple of 6 and *m* − 1 is a prime number; which covers a wide range of settings). We describe the construction details in Supplementary Methods. Using this, forming the HYPER pools and identifying putative positives becomes both simple and flexible.

### Performance under a common statistical model

We first study the performance of HYPER under a standard statistical model for group testing^20–25^ with possibly noisy tests. Specifically, we assume each individual is positive independently at random with probability *p*, where *p* is the disease prevalence. In addition, each test may be incorrect with some probability, i.e., the tests may be noisy. In particular, each test has a specificity of 1 − *α* and a sensitivity of *β*. Recall that specificity is the probability that the test returns negative given that it was supposed to (i.e., that it contained no positive individuals), and sensitivity is the probability that the test returns positive given that it was supposed to (i.e., that it contained at least one positive individual). This setting is different from the COVID-19 model we will consider in the following sections, but is also important to study since the potential applicability of HYPER extends beyond COVID-19. Here we present only our key results, with the details given in the Supplementary Methods.

For this analysis, we suppose *n* is a multiple of *m/q* so that the design has perfectly balanced pools, and let *k* = *nq/m* be the number of subjects per pool. This is not a significant restriction as *m/q* is typically a small integer in the examples we are interested in (e.g., in some simulations below we have *m* = 24 with *q* = 3). Denote *r* = 1 − *p* and 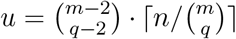. Then, we can show that the expected number 𝔼(*T*) of tests *T* used by HYPER (including the tests from the second stage) is upper bounded as

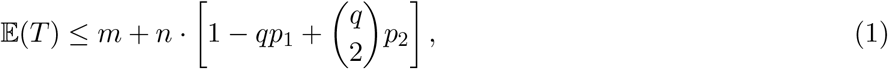

where *p*_1_ = 1 − *β* +(*β* − *α*) · *r*^*k*^ and 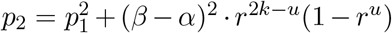. This inequality is in fact an equality when *q* = 1 or 2, in which case the above formula exactly characterizes the expected number of tests used by HYPER (Supplementary Methods). We have also derived a more general and sharper version of this upper bound that is valid for all *q* via the Dawson-Sankoff inequality^70;71^, which is a non-trivial generalization of the Bonferroni union-intersection inequalities from probability theory.

For *q* ≤ 2 and 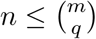, the corresponding overall average specificity (the probability that HYPER declares an individual to be negative given that the individual was negative) for HYPER is 1 − *αγ* and the overall average sensitivity (the probability that HYPER declares an individual to be positive given that the individual was positive) for HYPER is *β*^*q*+1^, where *γ* = [*β* + (*α* − *β*) · *r*^*k*−1^]^*q*^ (Supplementary Fig. 1). The false negative probability (the probability that a sample is positive given that it was declared negative) for HYPER is

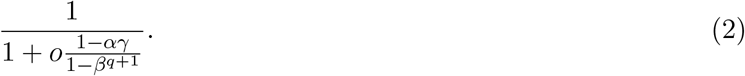

The true positive probability (the probability that a sample is positive given that it was declared positive) for HYPER is

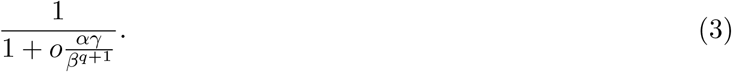

Here *o* = (1 − *p*)*/p* is the odds ratio of prevalence. Note also that the probabilities above are with respect to both the random test errors and the random positivity of the other individuals. In this noisy group testing model, sensitivity and specificity calculations have been reported in Bilder’s work for array designs^22^, but it is beyond our scope to do a full comparison. Note also that the sensitivity of HYPER under this model can be improved via error-correction of false negatives in stage one, e.g., by including individuals in only one negative pool among the putative positives^11^. However, doing so can come at the cost of efficiency, and analyzing this tradeoff is also beyond our present scope.

Focusing on the noiseless setting, i.e., test sensitivity and specificity close to 1 (*α* = 0, *β* = 1), enables some further investigation into which HYPER designs are most efficient. First, for large batches (*n* → ∞) and diminishing prevalence (*p* → 0), we can show that the optimal number of pools *m* for HYPER (with *q* = 2) is approximately *m/n* ≈ 2*p*^2*/*3^ − *p*. The corresponding approximate efficiency is 𝔼(*T*)*/n* ≈ 3*p*^2*/*3^ (including stage two tests; Supplementary Figs. 1g and 1h). This improves on Dorfman designs, for which the approximate efficiency is 𝔼(*T*)*/n* ≈ 2*p*^1*/*2^ for prevalence *p* → 0 going to zero^21^. In fact, this asymptotic efficiency matches that of three-stage Dorfman testing^21;64^. While more efficient designs for diminishing prevalence are available in the literature, they rely either on multiple stages^10;21^ or on taking *q* much larger^48–56^; each of which is outside our constraints. See Supplementary Methods for more discussion of some of these methods.

In the context of testing for widely-spread infectious disease, it is also important to consider fixed (non-diminishing) prevalence *p* > 0, i.e., the *linear regime*^35–37^. Recent works in this regime have considered two problem formulations where group testing increases efficiency. The first problem is to minimize the average number of tests used per individual by using any group testing method (i.e., any number of stages, any decoder, etc.), and is tackled, e.g., by fully-adaptive methods^36^. The second problem adds some real-life constraints by instead allowing only two-stage group testing methods with conservative decoding^37^, such as the array design and HYPER. We compared known lower bounds for these two problems with the upper bound for HYPER given above, *with m* and *q* numerically optimized for *n* = 6144 (Supplementary Fig. 2). As one would naturally expect, better efficiency (i.e., fewer tests on average) is achievable for problem 1, e.g., by using fully-adaptive methods^36^, since it is much less constrained than problem 2. For problem 2, which incorporates real-life constraints, HYPER turns out to be somewhat close to optimal. The gap between the lower bound for problem 2 and the upper bound for HYPER that arises at low prevalence is possibly due to: a) our additional constraint that *q* ≤ 3 (to aid real-life implementation), b) looseness in our upper bound for HYPER, and c) potential looseness in the lower bound^37^. Closing this gap is an exciting direction for future work.

To summarize, these results characterize the expected efficiency, sensitivity and specificity under a standard statistical model where tests have independent errors. This model captures important features of many applications beyond COVID-19 testing, and provides a useful setting to evaluate methods. Here we found that for noiseless tests, the efficiency of HYPER at low prevalence is competitive with other existing methods of comparable simplicity, and appears to be somewhat close to optimal for two-stage conservative testing with non-diminishing prevalence. Based on the specific application, the analysis can also be used to help guide the selection of HYPER designs that strike the appropriate balance of efficiency, sensitivity and specificity. For example, one might use a small number of splits *q* in settings where sensitivity loss due to independent test errors is of primary concern. On the other hand, if test errors are negligible, the best efficiency (for *q* = 2 splits) at low prevalence may be obtained by using *m/n* ≈ 2*p*^2*/*3^ − *p* pools per individual. Finally, the formulas for false negative and true positive probabilities help guide the interpretation of negative and positive results, respectively, declared by HYPER.

### Performance under a COVID-19 model

We study the performance of HYPER under the viral load based COVID-19 model of Cleary and Hay et al.^11^. It simulates: a) SARS-CoV-2 viral load kinetics in infected individuals; b) the dilution of viral loads during pooling that can lead to false negatives; and c) the evolution of infection prevalence in a large population over time during epidemic growth and decline. We focus here on a window during which the prevalence (i.e., fraction of individuals with positive viral load) increases exponentially from 0.03% to 2.46% (days 40–90 in our simulation; we consider other windows below) and individual testing has a sensitivity (i.e., fraction of positive individuals that are identified) of roughly 85% (Fig. 2). We compare HYPER with two existing state-of-the-art methods (Fig. 2a): plate-based array designs^9^ and P-BEST^8^. These methods use batches of *n* = 96 individuals (8 × 12 array; left panels) or *n* = 384 individuals (16 × 24 array and P-BEST; right panels). We consider both efficiency gain with respect to individual testing (number of individuals screened divided by average number of tests used, including stage-two tests) and average sensitivity (fraction of positive individuals identified after completion of all stages) of the methods.

**Fig. 2.**
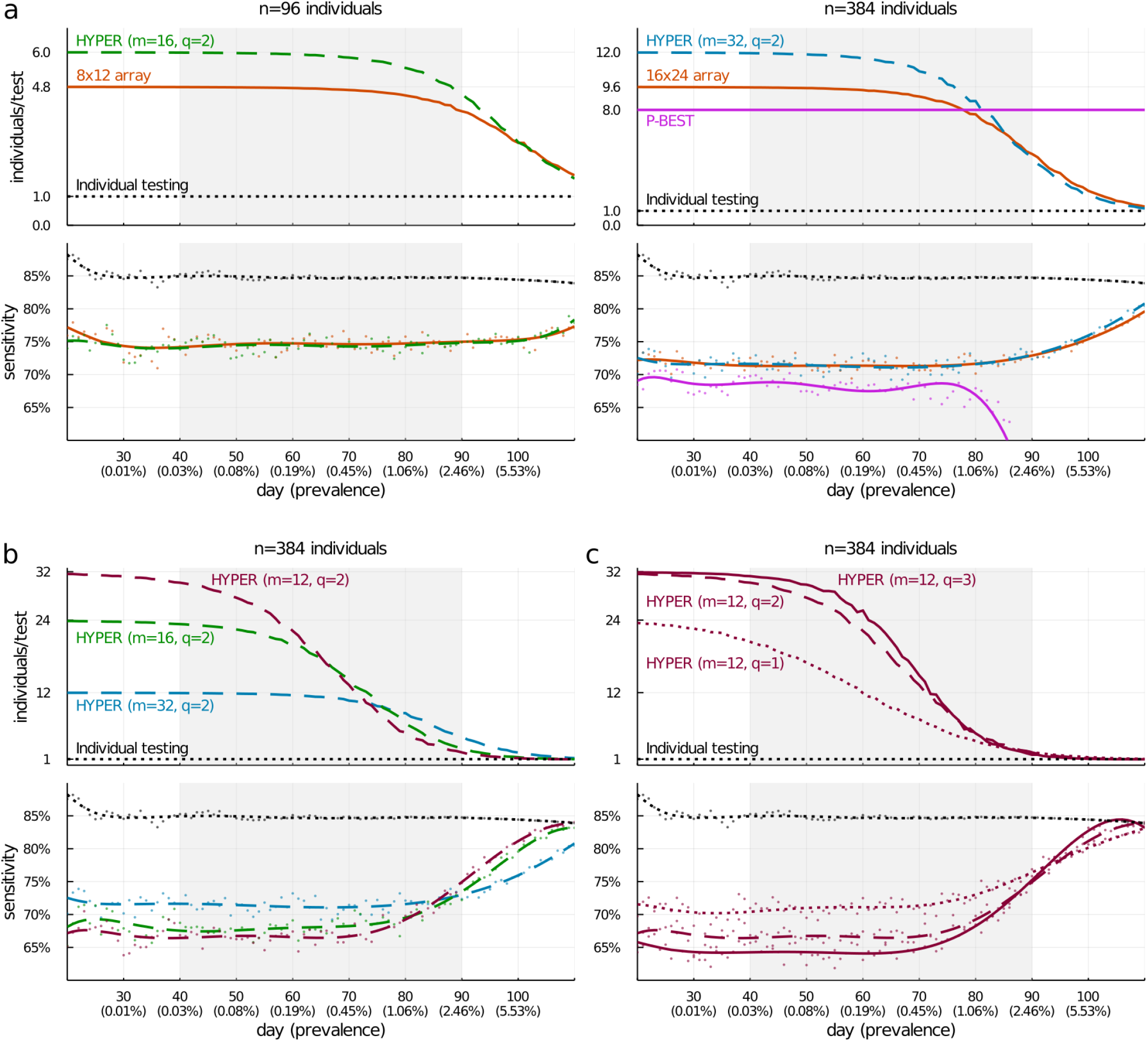
Efficiency and sensitivity of pooled testing during a simulated epidemic. Average values of efficiency (relative to individual testing) and sensitivity of a variety of pooling designs are shown for each day, with results averaged across 200,000 random trials. For sensitivity, raw averages are shown as dots with degree-8 polynomial fits overlaid as curves; the curves for efficiency depict raw averages. During the days 40–90 (highlighted), the prevalence grows exponentially from 0.03% to 2.46%. **a** Comparison of HYPER with alternative methods that use *n* = 96 individuals per batch (left panels) or *n* = 384 (right panels). The particular HYPER designs were chosen to have the same maximum pool sizes (*nq/m* = 12 for H_96,16,2_; *nq/m* = 24 for H_384,16,2_) as the array designs. Sensitivity (**a**, bottom panels) depends heavily on pool sizes, due to dilution of viral loads. **b**,**c** HYPER evaluated with varying numbers of pools (**b**, *m* = 32, 16, 12) and numbers of splits (**c**, *q* = 1, 2, 3). The designs are affected by the increasing prevalence over time to varying degrees. As prevalence increases, efficiency decreases (as more stage-two tests become necessary), while sensitivity increases (as larger viral loads begin to “rescue” small viral loads that would have been missed).

For *n* = 96 (Fig. 2a, left panels), we consider the 8 × 12 array and a HYPER design, H_96,16,2_, chosen to dilute samples a similar amount and thus potentially achieve a similar sensitivity. Our simulation shows that both methods indeed have similar sensitivity, which is about 10 percentage points lower than the sensitivity of individual testing (Fig. 2a, bottom-left panel). This is due to dilution of viral loads below the limit of detection during stage-one tests. While the array design is roughly 4.8 times more efficient than individual testing for much of the 50-day window (Fig. 2a, top-left panel), consistent with earlier studies^9^, the H_96,16,2_ design is roughly 6 times more efficient than individual testing. This corresponds to a 25% improvement in efficiency over the comparable array design, while achieving essentially the same sensitivity.

For *n* = 384 (Fig. 2a, right panels), we consider the H_384,16,2_ design, again chosen to potentially match the sensitivity of the array design (16 × 24) appropriate for this number of samples. The HYPER design is again roughly 25% more efficient than the array design (Fig. 2a, top-right panel) while having essentially equal sensitivity (Fig. 2a, bottom-right panel). In contrast to these two-stage methods, the one-stage approach of P-BEST has a constant efficiency gain of 8 times individual testing (Fig. 2a, top-right panel), but it significantly loses sensitivity around day 80 as prevalence grows (Fig. 2a, bottom-right panel). This is because the design and decoding algorithm are optimized for a prevalence around 1% and performance degrades beyond this operating point. In contrast, the sensitivity of HYPER increases around day 80 as prevalence increases. Notably, error-correcting does not appear to effectively handle the false negatives that arise here due to diluted viral loads falling below the limit of detection. P-BEST is generally the least sensitive among the three pooling strategies.

We also considered H_96,20,2_ and H_384,40,2_ designs that match the number of pools in the 8 × 12 and 16 × 24 array designs (Supplementary Fig. 3). They have efficiency essentially the same as their corresponding array designs, but with slightly higher sensitivities. For *n* = 384, we also considered HYPER designs that match the number of pools (H_384,48,2_) and the pool sizes (H_384,16,2_) of P-BEST (Supplementary Fig. 4). The H_384,48,2_ design has similar efficiency to P-BEST at low prevalence but is more sensitive. The H_384,16,2_ has similar sensitivity to P-BEST for much of the 50-day window highlighted, but is more efficient at low prevalence. As with the previous designs, the efficiency of HYPER declines as prevalence grows, eventually falling below the constant efficiency gain achieved by P-BEST around the same time that P-BEST loses sensitivity. We also compared with two random methods (random assignment and double-pooling) and a balanced square array method with holes along the diagonal (Supplementary Fig. 5). Both random methods have similar average performance to the corresponding HYPER designs. Compared with the balanced square arrays with holes, the HYPER designs (H_96,16,2_ and H_384,32,2_) use fewer pools and are roughly 25% more efficient but have correspondingly larger pools and are slightly less sensitive. Constructing balanced square arrays with the same number of pools as HYPER requires extending the method to place multiple individuals in some of the array cells (as discussed in Supplementary Methods). Comparing HYPER with this balanced array design using the same design parameters (*n, m, q*), we found that HYPER had similar or better performance, with a larger improvement in efficiency arising for more aggressive designs that used fewer pools and yielded greater efficiency at low prevalence (Supplementary Fig. 6a). We also compared with Reed-Solomon Kautz-Singleton (RS-KS) code-based designs (described in Supplementary Methods) with matching parameters, and found that HYPER again had similar or better performance with a greater improvement in efficiency when the design used fewer pools (Supplementary Fig. 6b).

Since the flexibility of HYPER allows for many designs, we also compared different HYPER designs and their various tradeoffs. Specifically, we considered various choices for the number of pools (Fig. 2b, *m* = 32, 16, 12) and the number of splits (Fig. 2c, *q* = 1, 2, 3). The average performance of the HYPER designs was generally consistent with those of the random assignment designs studied by Cleary and Hay et al.^11^. However, note that any given draw of a random design may perform better or worse based only on chance. HYPER produced the same overall performance with fixed *deterministic* designs. Specifically, the average efficiency gain of HYPER is roughly *n/m* early in the epidemic, during which low prevalence leads to pools frequently testing negative. Decreasing the number of pools *m* can significantly increase efficiency, but comes with a slight reduction in sensitivity. As the epidemic progresses, rising prevalence leads to increasingly many stage-two tests, though designs with larger *q* are more robust to this loss of efficiency. At the same time, larger *q* designs tend to be less sensitive. Overall, more efficient designs tend to be less sensitive, creating a trade-off that depends significantly on prevalence. One size does not fit all, underscoring the benefit of flexible designs like HYPER that can easily be adapted from one environment to another.

We also investigated how the balance of HYPER impacts the *consistency* of performance across individuals. In particular, we studied how the sensitivity and efficiency achieved for a single positive individual varies as a function of where that individual is placed in the design (Supplementary Fig. 7). As discussed above, uniform performance across individuals is important for real-world testing, since it can help make the treatment of individuals and the stage-two workload consistent. We compared HYPER with consecutive pooling, lexicographic pooling, random assignment pooling, double-pooling, balanced array pooling, and Reed-Solomon Kautz-Singleton (RS-KS) code-based pooling. Overall, the designs with balanced pools (consecutive pooling, double-pooling, balanced array pooling, RS-KS pooling, and HYPER) had uniform sensitivity. Designs with imbalanced pools use varying volumes from each individual and so dilute the viral loads to varying degrees, which can result in uneven sensitivity. For example, all three draws of random assignment had uneven sensitivity; for the first draw, the sensitivity was 75.7% for a positive individual in location 11 but 72.2% for location 7. The designs with balanced pool combinations (lexicographic pooling and HYPER) had uniform efficiency. A positive individual (if caught in stage one) is retested in stage two *along with any others assigned to the same pool combination*, so imbalanced pool combinations can lead to uneven amounts of stage-two testing. Consecutive pooling had uniform but lower efficiency; it used only a subset of the possible pool combinations but used them in a balanced way. Likewise, balanced array pooling and RS-KS pooling both had slightly uneven efficiency here. Each used a subset of the possible pool combinations, on which it was maximally but not perfectly balanced; in both cases some pool combinations were used twice while others were used once. HYPER, which perfectly balanced both pools and pool combinations, had uniform sensitivity and efficiency. Moreover, its median efficiency (5.68 individuals/test) and median sensitivity (74.4%) were generally among the best.

Finally, we studied how changes in test characteristics or epidemic dynamics affect performance. The first scenario we considered is tests with a limit of detection (LOD) reduced by a factor of 25 (Supplementary Fig. 8). A smaller LOD means that smaller viral loads are caught, resulting in a higher individual testing sensitivity around 95%. The sensitivity of the group testing methods increased concordantly, resulting in similar relative sensitivities to before. In general, an increase in sensitivity can lead to a decrease in efficiency, since more pools (correctly) test positive, but the impact appears to have been small here. Efficiency for the various methods was very similar to before, with HYPER enjoying essentially the same efficiency gains over individual testing. Overall, decreasing the LOD resulted in an increase in sensitivity across all the methods without much change in their relative performances.

The second scenario we considered was a population undergoing a sustained, two-wave epidemic^11^ (Supplementary Fig. 9). Namely, the simulated population has an initial wave, followed by a decline phase and subsequently another growth phase. Consistent with earlier studies^11^, sensitivity was lower for all methods (including individual testing) during the decline phase compared to the two growth phases. For example, sensitivity on day 104 (during decline) was generally lower than days 82 and 173 (during growth), even though the prevalence was around 1.0% on all three days. This difference can be explained by a general shift to the left in the distribution of nonzero viral loads on day 104 relative to days 82 and 173, due to a shift away from recent infections. As a result, positive viral loads are more likely to fall below the LOD. At the same time, the relative performance of the methods is similar to before. The HYPER designs are as sensitive as their corresponding plate-based array designs, while being roughly 20-25% more efficient for *n* = 96 and roughly 10-25% more efficient for *n* = 384. For *n* = 384, the HYPER design is generally more efficient than P-BEST (up to 50% more) until day 180 while also being generally more sensitive. After day 180, P-BEST is more efficient but at an additional cost of sensitivity. As before, prevalence appears to be one of the main factors driving efficiency. Overall, while the epidemic phases have different properties (e.g., due to differences in viral load distribution), the relative performance of HYPER designs appears to be fairly robust to these changes as well.

### Choosing a pooling method given resource constraints

In practice, decision makers must often choose a pooling method given limited capacity (on average) for daily testing and sample collection. One approach is to maximize the *number of individuals screened per day*, i.e., the number of individuals *n* per batch times the number of batches *b* that can be run per day, given the limited testing and collection capacities. This metric accounts for the impact of resource constraints. However, it does not represent the *actual* number of infected individuals that the population screen can identify. A very efficient method can screen numerous individuals but may miss all the infected ones if it lacks sufficient sensitivity. Hence, a more natural goal is to choose a design that maximizes the number of individuals screened times the average sensitivity, which we call the *effective number of individuals screened per day* or simply the “effective screening capacity” for short. A nice property of the effective screening capacity is that scaling it by prevalence measures the average number of infected individuals that are identified by the screen. This makes it especially meaningful in public health contexts, where the goal may be to find and isolate as many infected individuals as possible. Hence, in this section, we study the problem of maximizing the effective screening capacity over windows of time given resource constraints (limited amount of sample collection and testing). We evaluated the overall effective screening capacity for the epidemic window considered above (Fig. 2). We compared the best HYPER design (across a sweep of *n, m* and *q*; Supplementary Table 1) with individual testing, plate-based array designs^9^, and P-BEST^8^, all across a range of resource constraints.

We will first consider a few specific scenarios to understand the tradeoffs with each design, then consider a larger grid to get a picture of the overall trends. We begin with a testing-scarce setting (Fig. 3a) with an average testing capacity of 12 tests per day being far outstripped by an average sample collection capacity of 3072 samples per day. In this case, individual testing only screens 12 individuals and achieves an effective screening capacity of 10.2 individuals (the average sensitivity of individual testing is 84.8%, due to the presence of positive individuals with viral load below the limit of detection). The best HYPER design (maximizing the effective screening capacity, given the constraints), obtains an effective screening capacity of 122.2 individuals, roughly 12 times more than individual testing. It does so by pooling *n* = 192 individuals per batch into *q* = 2 of *m* = 6 pools with an average of *b* ≈ 0.9 batches run per day (recall that some of the testing capacity is needed for stage-two tests). Both array designs and P-BEST use more than 12 tests in a single run so do not appear here.

**Fig. 3.**
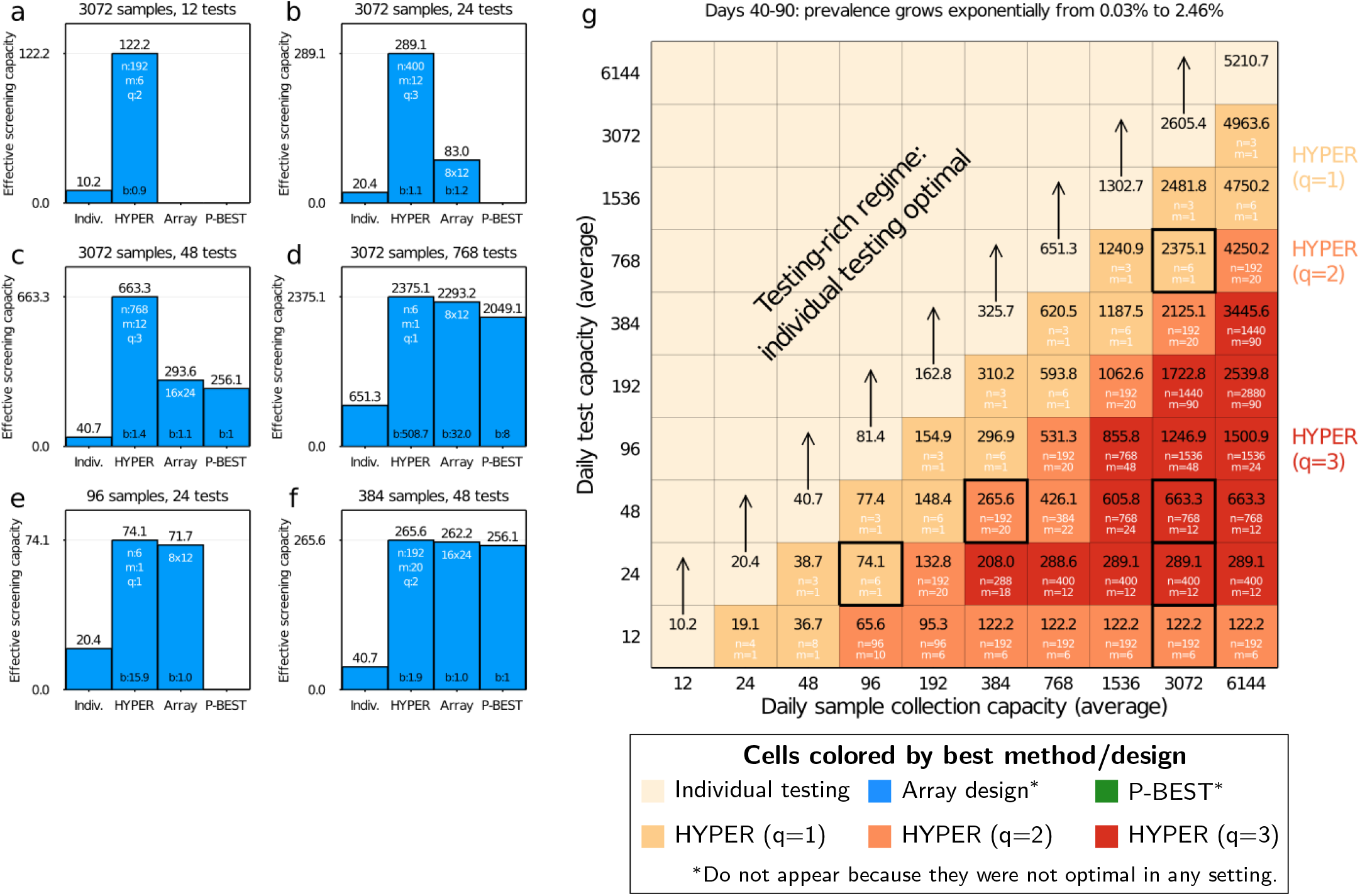
Comparison of pooling methods under resource constraints. HYPER designs (Supplementary Table 1) were evaluated together with individual testing, plate-based array designs^9^, and P-BEST^8^, across a range of sample collection and testing capacities. The basis for comparison was the average effective number of individuals screened per day across days 40-90 of the simulation (Fig. 2), during which the prevalence increases exponentially from 0.03% to 2.46%. Bar plots on the left depict the effective screening capacities (bar height) in a testing-scarce setting (**a**), followed by increasingly testing-rich settings (**b-d**), and then settings well-suited for the array designs and P-BEST (**e**,**f**) that have capacities matching their design parameters. When multiple designs for a given method were available within the constraints (i.e., various choices of HYPER designs, or a choice between the two 8 × 12 and 16 × 24 array designs), we use the most effective configuration and indicate it in white text within the appropriate bar. The average number of batches run per day is noted at the bottom of each bar. **g** Expanded comparison to a grid of sampling and testing budgets. Each cell is colored by the best method (where we separately identify HYPER designs with *q* = 1, 2, 3 splits in shades of orange / red), and shows the corresponding effective screening capacities (in black text). The best design configuration is written in white text. For HYPER, we write the number of individuals per batch *n* and the number of pools *m* for the best configuration; cell color already indicates the number of splits *q*. Note that *n* and *m* often do not match the daily sampling and testing budgets, respectively, since multiple batches can be run per day. The cases from (**a-f**) are outlined in black. See Supplementary Fig. 10 for additional details.

As testing capacity grows to 24 then 48 tests (Figs. 3b and 3c) with sample capacity unchanged, larger effective screening capacities become possible by using larger designs, including the 8 × 12 array followed by the 16×24 array and P-BEST. HYPER adapts to these settings as well, and the larger designs here are accompanied with a larger number of splits *q*. HYPER remains the most effective overall. For a testing capacity of 24 daily tests, HYPER achieves 289.1 effective individuals screened, which is over 14 times as many as individual testing (20.4 effective individuals screened) and over 3.4 times as many as the array designs (83.0 effective individuals screened). P-BEST cannot be run since it uses more tests than available, so it achieves zero effective individuals screened. For a testing capacity of 48 daily tests, HYPER achieves 663.3 effective individuals screened, which is over 16 times that of individual testing (40.7 effective individuals screened), over 2.2 times that of the array designs (293.6 effective individuals screened), and over 2.5 times that of P-BEST (256.1 effective individuals screened).

When the capacity grows to 768 tests per day (Fig. 3d), i.e., a quarter the sample collection capacity, the pooled designs remain more effective than individual testing, but now by less than 4 times. In this increasingly testing-rich regime, P-BEST and the array designs are sample-constrained and under-utilize testing resources.

P-BEST uses only *mb* = 384 of the 768 available tests, since all 3072 available samples are tested after *b* = 8 batches of *n* = 384 samples. The same is true for the 16 × 24 array design, although additional tests will also be used in stage two. The most effective HYPER design H_6,1,1_ corresponds to simple Dorfman testing, uses roughly 508 tests in the first stage, and achieves 2375.1 effective individuals screened.

Finally, we considered two settings well-tailored for the array designs and P-BEST: 96 samples with 24 tests (Fig. 3e) for which the 8 × 12 array is well-suited (recall that extra testing capacity is needed for stage 2), and 384 samples with 48 tests (Fig. 3f) for which the 16 × 24 array and P-BEST are well-suited. In particular, these are settings where the number of available individuals and pools closely match (a multiple of) the number of individuals and pools used by these designs in a single batch. This can help them maximally utilize both the testing and sample collection capacities, i.e., neither resource is under-utilized. The array designs and P-BEST performed similarly to HYPER in these favorable cases, but notably HYPER remained slightly more effective: 74.1 vs. 71.1 effective individuals screened for the first scenario, and 265.5 vs. 262.2 effective individuals screened for the second.

Expanding this analysis to a grid of sampling and testing capacities gives a broad view of overall trends. We consider a sweep with each ranging from 12 to 6144 (Fig. 3g). Note first that for all sample collection capacities, the effective screening capacity grows as testing capacity scales up, until testing capacity matches or outpaces sample collection capacity. Individually testing all samples collected is most effective from that point on. Likewise, for any given testing capacity, the effective screening capacity rises as sample collection capacity grows, eventually reaching an upper limit at which testing capacity becomes the limiting factor. Pooled testing increases this upper limit, enabling an effective number of individuals screened far beyond the actual number of available tests. For example, for a testing capacity of 96 tests per day, pooled testing achieves an effective screening capacity of up to 1500.9 individuals per day, which is over 18 times the effective screening capacity of 81.4 individuals per day achievable by individual testing.

Across the testing-constrained regime, i.e., testing capacity below sample collection capacity, HYPER out-performed both P-BEST and the array designs (Fig. 3 and Supplementary Fig. 10). As observed before (Figs. 3a to 3f), the flexibility of HYPER generally makes it more readily tailored and optimized for each setting. Dorfman testing (i.e., HYPER with *q* = 1) is most effective when testing capacity is within a quarter of the sample capacity. However, as sample capacity begins to further outstrip testing capacity, combinatorial designs that involve more individuals *n* per batch and that use more splits *q* become most effective, consistent with earlier studies of analogous random designs^11^.

The above results capture the average effectiveness of each method when deployed over a 50-day window during epidemic spread. We next investigated effectiveness on individual days, each corresponding to a different fixed prevalence. At low to moderate prevalence of 0.1% (Supplementary Fig. 11), 1.06% (Supplementary Fig. 12), or 1.36% (Supplementary Fig. 13), HYPER is consistently the most effective strategy across all settings. In an intermediate range with prevalence of 1.48% (Supplementary Fig. 14) and 2.46% (Supplementary Fig. 15) there is a subset of scenarios in which P-BEST outperforms HYPER, although the performance of each method is nearly equivalent in these settings. Outside these settings P-BEST is either not viable or substantially under-performs HYPER. At a higher prevalence of 3.15% (Supplementary Fig. 16) HYPER again performs best across all scenarios. We did not observe any scenarios in which the array designs were most effective.

The 8 × 12 and 16 × 24 plate-based array designs^9^ proposed for COVID-19 pooled testing and P-BEST^8^ are instances of the general classes of array designs and code-based designs, respectively. Hence, we also compared HYPER with general balanced array designs and Reed-Solomon Kautz-Singleton (RS-KS) code-based designs (both described in Supplementary Methods). We investigated the effectiveness of balanced array designs optimized across the same set of design parameters as HYPER (Supplementary Table 1) for prevalences of 0.1% and 1.06% (Supplementary Figs. 17 and 18). HYPER was either more effective or about as effective as the balanced arrays across the grid of resource constraints and was significantly more effective in important testing-constrained settings. In some settings, greater efficiency enabled HYPER to use more aggressive design parameters than balanced arrays (e.g., Supplementary Figs. 17a and 18a). For a prevalence of 0.1% and a capacity of 384 samples and 12 tests per day, this yielded an effective screening capacity of 231.6 individuals per day for HYPER, which was a roughly 38% improvement over balanced arrays (167.7 effective individuals screened). In other settings, HYPER was more effective with the same design parameters. For example, for a prevalence of 0.1% and a capacity of 1536 samples and 24 tests per day (Supplementary Fig. 17b), HYPER had a roughly 25% larger effective screening capacity than the balanced array (624.1 vs. 496.9 effective individuals screened) that had the same design parameters.

The RS-KS design was limited to a subset of the design parameters considered for HYPER (as discussed in Supplementary Methods and indicated in Supplementary Table 1), so we optimized this method over only those parameters. In addition to HYPER (optimized over the full set of Supplementary Table 1), we compared with a Restricted HYPER that was optimized over the same subset of parameters as allowed by RS-KS. For prevalences of 0.1% and 1.06% (Supplementary Figs. 19 and 20), both HYPER and Restricted HYPER were either more effective or about as effective as RS-KS across the grid of resource constraints and were significantly more effective in important testing-constrained settings. As with balanced arrays, HYPER and Restricted HYPER sometimes achieved greater effectiveness than RS-KS using the same design parameters (e.g., Supplementary Fig. 19b) and sometimes did so with more aggressive design parameters (e.g., Supplementary Figs. 19a and 20a). In these cases, the increase in effectiveness over RS-KS was similar to those over balanced arrays.

Note that for both the balanced array and RS-KS designs, we considered extended variants that allowed design parameters with arbitrarily many individuals (as discussed in Supplementary Methods). HYPER more significantly outperformed the non-extended versions of these designs. They do not allow more than (*m/q*)^*q*^ individuals, but the most effective design parameters in testing-constrained settings often used more individuals. One could also consider variants of the above designs formed by concatenating *k* disjoint copies of a design with *n* individuals and *m* pools to obtain a design with *kn* individuals and *km* pools. Note, however, that such designs are equivalent to running *b* = *k* batches of the original design so were already implicitly considered in the above analysis.

## Discussion

Our results demonstrate the effectiveness of a new family of pooling designs that are adaptable to any number of samples, with only mild conditions on the number of pools, while remaining maximally balanced in three senses (number of assignments per individual, pool, and combination of pools). This flexibility is critical to selecting appropriate designs under the widely varying global demands and capabilities for SARS-CoV-2 testing. In addition, the balanced nature of the designs ensures uniform treatment of samples and facilitates robust and simple implementation. Despite the simplicity of implementing HYPER, the existence and construction of the designs relies on deep mathematical results, such as Baranyai’s theorem from combinatorics^66^ and Beth’s construction from number theory^67;68^.

Our evaluation of HYPER in both a general statistical framework and a SARS-CoV-2 specific simulation can be used to guide the choice of design, depending on the setting and purpose of testing. For our general statistical setup, where each test has specificity 1 − *α* and sensitivity *β* independent of all other tests, we showed that using roughly *m/n* ≈ 2*p*^2*/*3^ − *p* pools per individual maximizes the efficiency of HYPER designs, and we derived that HYPER has sensitivity *β*^*q*^. When selecting a design for SARS-CoV-2 testing, one must also account for the impact of dilution in pooled tests (leading to false negatives), among other effects like the evolution of viral loads over time. These considerations are important for selecting designs that remain effective during epidemic spread, which is critical in the ongoing pandemic and in preparations for the future. Here, insight can be gained from our results on a realistic COVID-19 simulation^11^. For example, the sensitivity of HYPER now depends on both the number of splits *q* and the number of pools *m*. Reducing the number of pools *m* means each pool will contain more individuals, leading to more dilution and lower sensitivity. Since we also do not correct for false negatives, the sensitivity is lower than individual testing. In theory, adding error correction could increase sensitivity, but doing so is most effective when false negative PCR results are independent across tests. When most false negative results come from diluting positive samples below the LOD (as in our simulations), then error correction is not as effective (as we observed with P-BEST) because the failures are not independent. The sensitivity of HYPER is generally best when using designs with fewer splits *q* or more pools *m*. However, doing so generally reduces efficiency at low prevalence. The results illustrate a general trade-off between sensitivity and efficiency that must be balanced depending on the setting.

Given the potential application of HYPER to future epidemics, it is important to also consider changes in viral kinetics and epidemic dynamics. For example, if viral loads peak later, more false negatives might come from individuals who have been recently infected but still have a small viral load. For infection control, it is critical to identify and isolate positive individuals soon after infection to reduce the probability of transmission. The change in viral kinetics might result in more false negatives in these important cases. Moreover, the difference in sensitivity between growth and decline phases might be altered, since this difference arises from differing viral loads in the recently infected versus those late in infection. However, shifting distributions of viral loads generally affect both individual testing and pooled testing, and they appeared to do so concordantly across methods in our analysis (Supplementary Figs. 8 and 9). Another important aspect is how fast the epidemic spreads, which depends on the particular virus. If the spread is slower, prevalence may remain low for a longer time, making it possible to use HYPER designs that are very effective but only at low prevalence. These designs can gain efficiency at low prevalence by sacrificing it at high prevalence. A more stable prevalence also makes it easier to select a single design that is effective over time. For example, consider the testing-constrained setting of Fig. 3a, where the best HYPER design (H_192,6,2_) achieved an effective screening capacity of 122.2 individuals across a window of time during which the prevalence grew from 0.03% to 2.46%. This design had to strike a balance between being effective early on (at low prevalence) and later on (at high prevalence). In contrast, for a fixed low prevalence of 0.1% (Supplementary Fig. 11a), the best HYPER design (H_384,8,2_) reached an effective screening capacity of 231.6 individuals, a nearly two-fold improvement. Overall, while a future epidemic would likely require a reevaluation of all group testing methods that carefully accounts for the specific features of that epidemic, the results here illustrate a general robustness for HYPER that makes it a promising candidate.

While pooled testing can substantially increase effectiveness depending on laboratory capacity and prevalence, it is important to also consider the added logistical challenges. Notably, the gains in testing effectiveness that we demonstrate above do not account for the additional pipetting steps during pooling, or the logistical cost of temporarily storing and retrieving samples for stage two testing. However, simple (Dorfman) pooling designs are receiving increasing interest^4–7;17;72^ for real-world testing, demonstrating that these logistical challenges can be overcome in practice in a variety of settings. In comparison to Dorfman designs, more complex designs (with *q* > 1) will require up to *q* times as many pipetting steps during stage one pooling. Depending on the relative timing and cost of each step in the protocol, this may shift the relative favorability of the strategies considered above. In particular, P-BEST, with *q* = 6 or more, may become relatively unfavorable if pooling steps are expensive, while plate-based array designs, which utilize multichannel pipettes, may become more favorable. We note that we have previously validated a HYPER design (H_96,6,2_)^11^ in the laboratory, and found that with an unoptimized workflow, stage one pooling of 192 samples could be completed manually in under 90 minutes. It is therefore especially likely that gains in effectiveness may outweigh additional logistical costs in settings that are constrained by testing throughput (e.g., due to limited reagents or equipment) but have an excess of laboratory technician capacity.

An important strength of the conservative decoder we used here is its conceptual simplicity, which can help reduce the risk of mistakes in practice. Moreover, it makes it possible to quickly illustrate (Fig. 1a) and explain the method to those who may not yet be familiar with group testing. These could include labs implementing group testing for the first time, as well as patients receiving the results. Additionally, conservative decoding does not declare positives from the pooled stage one tests; positives are only declared after a positive individual test in stage two. This can further help clinicians and patients interpret and act on positive results. Put together, these benefits make conservative decoding an appealing choice in practice, though other decoders with similar features may exist and could also be of interest. Alternatively, one might consider more sophisticated decoders that may further reduce stage two tests, e.g., by identifying some definite positives directly after stage one. Some methods also have more computationally efficient decoding that can help save some time. For COVID-19 testing, we expect that preparing and running the tests will typically account for much of the time spent, making these potential savings small, but labs will need to assess this given their particular resources and constraints. Although the alternative pooling designs we evaluated here were originally published as fixed designs, potentially optimized for a fixed prevalence (P-BEST), it is possible they could be adapted to different settings.

The 8 × 12 and 16 × 24 array designs may be adapted to any grid that fits on standard (96-well or 384-well) laboratory plates. However, our results suggest that HYPER designs could in general match the sensitivity of these array designs while having greater efficiency. Indeed, the 8 × 12 and 16 × 24 array designs had sub-optimal effectiveness in every setting we considered, including those tailored to best fit each array. Moreover, we found that in every setting we considered, HYPER was either as effective as or substantially more effective than the broad class of balanced arrays. P-BEST is a specific code-based design and can in principle be adapted to different settings, although it is unclear how flexible it can be while retaining balance, and properly doing so requires some expertise and experience. To use it, one might need to not only identify and generate the appropriate Reed-Solomon code but also properly retune the decoder algorithm. Here we used the design and decoder provided online (https://github.com/NoamShental/PBEST) by the authors. As with the 8 × 12 and 16 × 24 arrays, we found that HYPER was generally as effective as or more effective than P-BEST in every setting we considered, including those tailored for P-BEST. Comparing with the broad class of Reed-Solomon Kautz-Singleton (RS-KS) code-based designs, we found that HYPER was also as effective as or substantially more effective than this broad set of code-based designs in every setting we considered.

So far we have limited HYPER to *q* ≤ 3. This has the advantage of reducing the additional logistical burden (and potential for error) that comes with splitting samples into more pools. Moreover, the efficient construction of hypergraph factorizations is highly nontrivial for *q* > 3. However, higher *q* can have several advantages. For example, individuals in the same hyperedge (i.e., assigned to the same combination of pools) are identified as putative positives together as a block even if only one of them is actually positive. Using a higher *q* can significantly increase the number of hyperedges 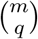, reducing the number of individuals sharing a single hyperedge. Results for HYPER here also indicated that high *q* designs can be highly effective when the sample capacity significantly outstrips testing capacity, consistent with earlier studies of analogous random designs^11^. Likewise, greater efficiency can be obtained by using a multi-stage approach with more than two stages, which is also more logistically challenging. In practice, one must weigh these opportunities for greater effectiveness against the increased complexity. Such designs may be especially promising for labs with access to robotic pipetting platforms.

To conclude, we present a simple, efficient and flexible pooled testing strategy that can be easily tailored and implemented without specialized expertise or equipment. To further facilitate implementation, we provide an online tool available at http://hyper.covid19-analysis.org that makes it easy to generate and carry out designs for a broad range of settings.

## Data Availability

Simulated data can be regenerated using the accompanying code.

## Data availability

Data can be regenerated using the accompanying code.

## Code availability

Code is available at https://github.com/dahong67/hyper-group-testing.

## Acknowledgements

D.H. was supported by the Dean’s Fund for Postdoctoral Research of the Wharton School and NSF BIGDATA grant IIS 1837992. R.D. and X.L. were supported by a grant from the Partners in Health. E.D. was supported in part by NSF BIGDATA grant IIS 1837992.

## Author contributions

All authors contributed to the design of the study and to discussions of all aspects. D.H., R.D., and E.D. performed the theory development and analysis under the general statistical model. D.H., X.L., and B.C. performed and analyzed the simulations under the COVID-19 model. R.D. and X.L. developed the interactive online tool. D.H., B.C., and E.D. wrote the first draft of the manuscript. All authors reviewed and edited the manuscript.

## Supplementary Methods

### Maximally balanced designs

We seek designs that are maximally balanced in three ways. Here we illustrate each way through some examples:

1. Assign all individuals to the same number *q* of pools

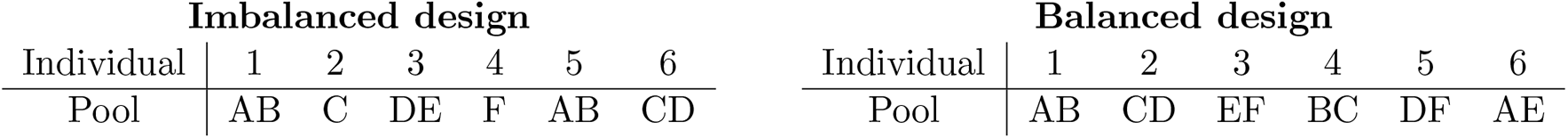 In the imbalanced design, all individuals are in two pools except individuals 2 and 4, who are in one pool. The balanced design assigns all individuals to two pools.
2. Assign the *m* pools as evenly as possible

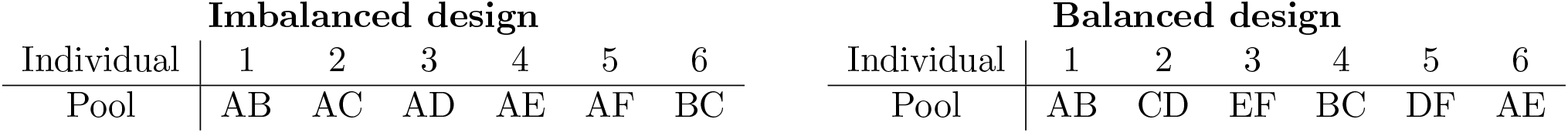 In the imbalanced design (lexicographic pooling), pool A has five individuals, pools B and C have two, and pools D-F have only one. All pools in the balanced design have two individuals.
3. Assign the 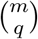 possible pool combinations as evenly as possible

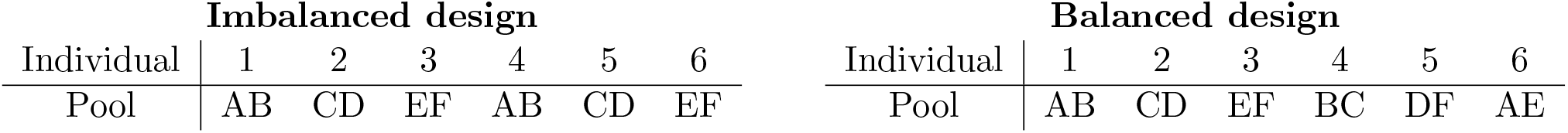 In the imbalanced design (consecutive pooling), pool combinations AB, CD and EF are each used twice but none of the other combinations (e.g., AC) are used.

Formally, *maximally balanced* designs can be defined as follows:

#### Definition 1.

*Let P* ∈ {0, 1}^*m*×*n*^ *denote a pooling design with n individuals and m pools, where P*_*ij*_ = 1 *means that pool i contains individual j and P*_*ij*_ = 0 *otherwise. We say P is maximally balanced if for some q*

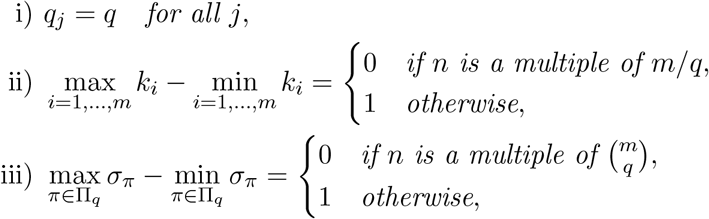

*where* 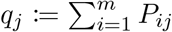 *is the number of pools individual j is assigned to*, 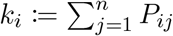 *is the size of pool i, σ*_*π*_ *is the number of individuals assigned to the pool combination π, and* Π_*q*_ := {*π* ⊆{1, …, *m*} : |*π*| = *q*} *is the set of all possible combinations of q pools chosen from* {1, …, *m*}.

### Balanced array designs with holes

The plate-based array designs^9^ proposed for COVID-19 testing use 8 × 12 and 16 × 24 arrays. This corresponds to plate sizes common in laboratory environments, making these choices convenient in practice. However, it results in imbalanced pools (Fig. 1b) because the row pools are larger than the column pools. To address this, one could instead use square arrays, though these may no longer corresponds to common physical plate sizes in the lab. In these cases, the arrays become primarily a conceptual tool for constructing the design. In particular, taking *k* = *m/*2, one could use a *k* × *k* array with *n* = *k*^2^ = (*m/*2)^2^ individuals. This design has balanced pools since all pools contain *k* individuals. However, this is somewhat inflexible since *n* must then be a perfect square. One way to address this is to partially fill a larger array design than needed, i.e., consider a square array with holes.

For example, to replace the 8 × 12 array, which covers *n* = 96 individuals, one could use a 10 × 10 array with four holes. Note, however, that the holes must be placed carefully to preserve maximal balance of the pools. For example, if the holes are all placed on the first row, then that row pool will have four fewer individuals than the rest of the row pools. Instead, the holes should be placed along diagonals of the array. In this way, the pools in the array stay maximally balanced.

Taking this approach for general *n* corresponds to filling the array along the diagonals. For *k* = 3, e.g., individuals may be incrementally filled into a 3 × 3 array as follows:

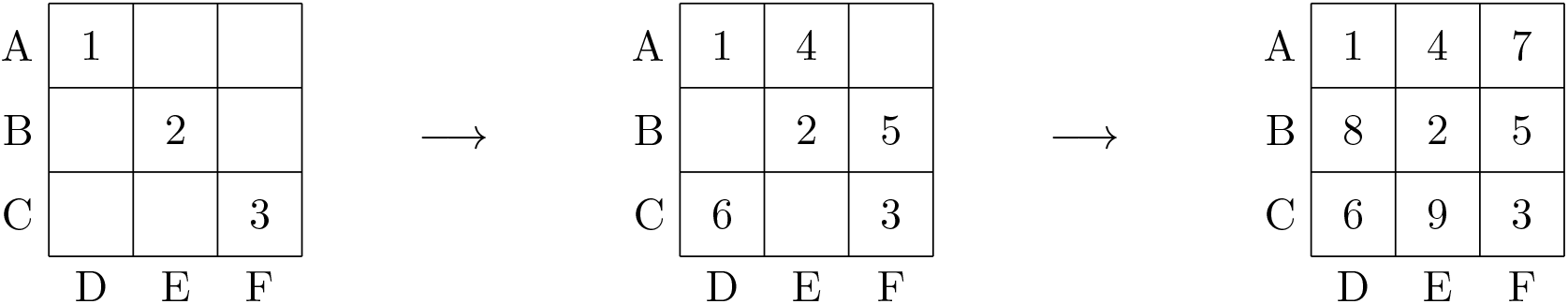

Namely, we assign individuals to cells (and the corresponding pools) as follows:

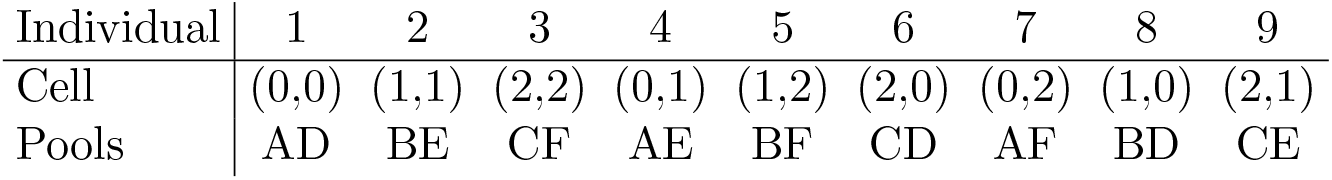

| Individual | 1 | 2 | 3 | 4 | 5 | 6 | 7 | 8 | 9 |
| --- | --- | --- | --- | --- | --- | --- | --- | --- | --- |
| Cell | (0,0) | (1,1) | (2,2) | (0,1) | (1,2) | (2,0) | (0,2) | (1,0) | (2,1) |
| Pools | AD | BE | CF | AE | BF | CD | AF | BD | CE |

i.e., the sequence of assignments to cells are generated for general *k* as

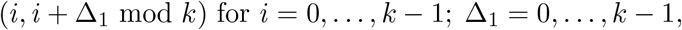

and the pool assignments are given by the rows and columns.

This approach is limited to producing designs with at most *k*^2^ individuals. However, it can be extended to allow for more individuals, i.e., to create designs with *n* > *k*^2^, by going back to the start and placing multiple individuals in each cell. Namely, individual *k*^2^ + 1 is placed in the same cell as individual 1, individual *k*^2^ + 2 is with individual 2, and so on. This parallels how HYPER handles 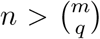 individuals. Here it produces a design with maximally balanced pools. Pool combinations are not maximally balanced since some combinations are unused while others are assigned twice, but they are maximally balanced among those that are used.

The design can also be extended to three-dimensional arrays that assign each individual to *q* = 3 pools. For *k* = 3, e.g., we have the following 3 × 3 × 3 array with pools along each slice (pool B shaded in to illustrate):

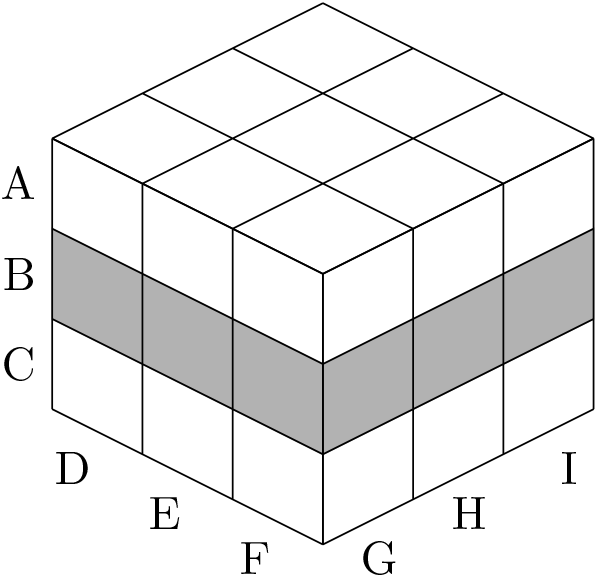

The sequence of assignments to cells are now generated as

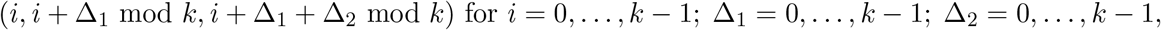

yielding the following pool assignments for *k* = 3:

| Individual | 1 | 2 | 3 | 4 | 5 | 6 | 7 | 8 | 9 |
| --- | --- | --- | --- | --- | --- | --- | --- | --- | --- |
| Cell | (0,0,0) | (1,1,1) | (2,2,2) | (0,1,1) | (1,2,2) | (2,0,0) | (0,2,2) | (1,0,0) | (2,1,1) |
| Pools | ADG | BEH | CFI | AEH | BFI | CDG | AFI | BDG | CEH |
| Individual | 10 | 11 | 12 | 13 | 14 | 15 | 16 | 17 | 18 |
| Cell | (0,0,1) | (1,1,2) | (2,2,0) | (0,1,2) | (1,2,0) | (2,0,1) | (0,2,0) | (1,0,1) | (2,1,2) |
| Pools | ADH | BEI | CFG | AEI | BFG | CDH | AFG | BDH | CEI |
| Individual | 19 | 20 | 21 | 22 | 23 | 24 | 25 | 26 | 27 |
| Cell | (0,0,2) | (1,1,0) | (2,2,1) | (0,1,0) | (1,2,1) | (2,0,2) | (0,2,1) | (1,0,2) | (2,1,0) |
| Pools | ADI | BEG | CFH | AEG | BFH | CDI | AFH | BDI | CEG |

Analogous to the *q* = 2 balanced arrays, this approach is limited to producing designs with at most *k*^3^ individuals but can be extended by cycling back through the pool assignments as is done in HYPER.

These extensions make it possible to generate array designs with parameters matching those considered for HYPER (Supplementary Table 1), allowing for further comparison with HYPER.

### Reed-Solomon Kautz-Singleton code-based designs

We consider the celebrated Kautz-Singleton^32^ construction that converts a *b*-nary code (often a Reed-Solomon^73^ code) into a binary design matrix. This design has recently been shown to be order optimal for the “probabilistic group testing problem” under certain assumptions^74^.

For the reader’s convenience, here we detail the steps involved in constructing the Reed-Solomon Kautz-Singleton (RS-KS) design used in this paper in our notation. The design is parameterized by (*n, m, q*), together with an additional parameter *f* used in the construction. The first step is to construct a *m/q*-nary matrix from a Reed-Solomon code as follows:

1. Let *b* := *m/q*, and let *F*_*b*_ be the finite field of size *b* (so *b* must be a prime power).
2. Let *x* be a generator of *F*_*b*_, i.e., *F*_*b*_ = {0, 1, *x*, …, *x*^*b*−2^}, with *x*^*b*−1^ = 1.
3. Let 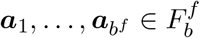 be an enumeration of 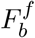, e.g., in lexicographic order:

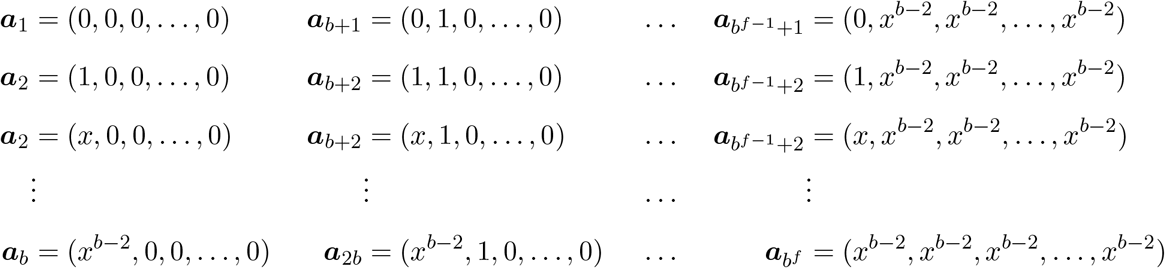
4. Form the *q* × *b*^*f*^ matrix with *b*-nary entries:

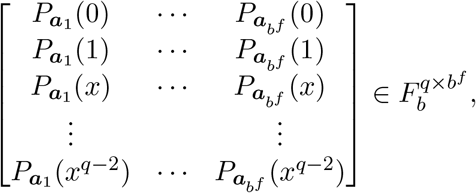

where 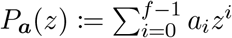 for any 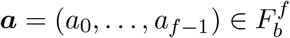 and any *z* ∈ *F*_*b*_. This requires that *q* ≤ *b* (so the evaluation is at distinct elements), and *f* ≤ *q* (so it is injective; we need this to be a meaningful code).

The second step, the Kautz-Singleton construction, is to convert the *b* = *m/q*-nary matrix into a binary design matrix by replacing each letter of the code by a binary column vector {0, 1}^*b*^ with a one-hot encoding, i.e.,

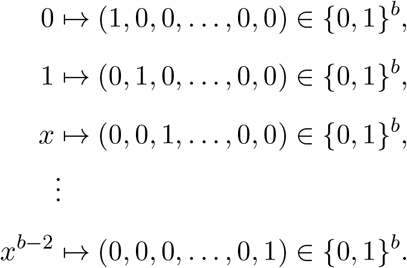

This gives a *qb* × *b*^*f*^ binary design matrix with *q* ones in each column. Since *m* = *qb*, the size of the matrix is equivalently *m* × (*m/q*)^*f*^.

Note that this approach is limited to producing designs for which

- *f* ≤ *q*.
- *n* = (*m/q*)^*f*^
- *m/q* ≥ *q*, i.e., *m* ≥ *q*^2^, and
- *m/q* is a prime power.

Note that the requirement that *f* ≤ *q*, means that for the choices *q* = 2 and *q* = 3 we consider, the relevant designs are: (*q* = 2, *f* = 2), (*q* = 3, *f* = 2), and (*q* = 3, *f* = 3).

For *n* ≤ (*m/q*)^*f*^, we consider truncating the design by taking the first *n* columns to obtain the desired *m* × *n* binary design matrix. The *m* rows correspond to the *m* pools, the *n* columns correspond to the *n* individuals, and the *q* ones in each column identify to which pools each individual is assigned.

For *n* > (*m/q*)^*f*^, we consider extending the design in a straightforward way by recycling the pool assignments, i.e., by concatenating the design matrix with copies on the right until it has at least *n* columns total. Namely, individual (*m/q*)^*f*^ + 1 is assigned the same pools as individual 1, individual (*m/q*)^*f*^ + 2 is assigned the same pools as individual 2, and so on. This parallels how HYPER handles 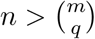 individuals.

The resulting design parameters from Supplementary Table 1 that can be handled by this construction are indicated there by asterisks (∗). We found that for all these designs, the pools were maximally balanced and the pool combinations were maximally balanced among those that were used. When *m/q* is not a prime power, we considered using a prime power *b* larger than *m/q* in the construction then dropping *qb* − *m* rows to obtain *m* final rows, but doing so produced columns with less than *q* ones (i.e., those individuals were not placed in *q* pools). This could be addressed by removing those columns, but in the cases we tried, this led to the pools being unbalanced.

#### Example

We work out the construction for *n* = 9, *m* = 6, *q* = 2, *f* = 2 as an illustrative example. Here, *b* = 3 so we have *F*_3_ = {0, 1, 2}, i.e., (*Z*_3_, +, ·), with generator *x* = 2. Next, we have the enumeration

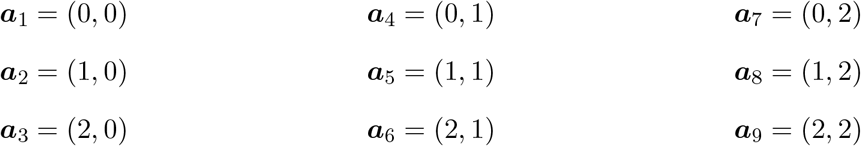

which yields the following *b*-nary matrix

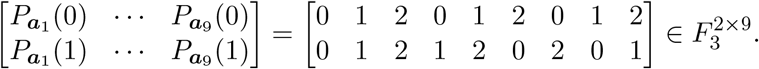

Converting to a binary matrix finally yields the following design:

|  | 1 | 2 | 3 | 4 | 5 | 6 | 7 | 8 | 9 |
|---|---|---|---|---|---|---|---|---|---|
| A | 1 | 0 | 0 | 1 | 0 | 0 | 1 | 0 | 0 |
| B | 0 | 1 | 0 | 0 | 1 | 0 | 0 | 1 | 0 |
| C | 0 | 0 | 1 | 0 | 0 | 1 | 0 | 0 | 1 |
| D | 1 | 0 | 0 | 0 | 0 | 1 | 0 | 1 | 0 |
| E | 0 | 1 | 0 | 1 | 0 | 0 | 0 | 0 | 1 |
| F | 0 | 0 | 1 | 0 | 1 | 0 | 1 | 0 | 0 |

Note that this has balanced pools and maximally balanced pool combinations, but does not use all pool pairs (e.g., the pool pair AB is unused).

### Simulation under a COVID-19 model

We performed simulations studies using the COVID-19 model of Cleary and Hay et al.^11^. The model first simulates viral loads for a large population of *n*_pop_ = 12, 500, 000 individuals across *d*_pop_ = 357 days during which the epidemic grows then declines. It captures the evolution of both: a) viral loads within each individual, i.e., within-host viral kinetics, and b) infection prevalence in the overall population. See Cleary and Hay et al.^11^ for a detailed description. The main output we use is a matrix 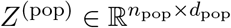 of the population viral loads, where 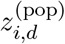 is the viral load of individual *i* on day *d*.

Next, the model simulates pooled testing to determine the average efficiency gain (with respect to individual testing) and average sensitivity for each day. For the reader’s benefit, we detail the process here. For HYPER designs, i.e., H_*n,m,q*_, the simulation proceeds for each trial *r* of day *d* as follows:

1. Draw *n* individuals uniformly at random from the population. Let *z*_1_, …, *z*_*n*_ be their viral loads that day. That is, draw *n* indices *k*_1_, …, *k*_*n*_ uniformly at random from the set {1, …, *n*_pop_} (with replacement), and let 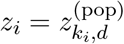. Put another way, 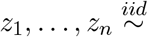 Uniform 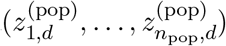.
2. Generate the *sampled* viral load for each of the *m* pools ℐ _1_, …, ℐ _*m*_ ⊆{1, …, *n*} as follows:

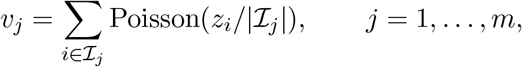

where | ℐ _*j*_| is the size of pool *j*, i.e., the number of individuals assigned to it.
3. Compute stage-one pooled testing results:
  - if *v*_*j*_ > LOD then pool *j* tests positive, where the LOD (limit of detection) we use is 100.
  - otherwise, pool *j* tests negative with probability 0.99 (i.e., the false positive rate of PCR results is 1%).
4. Select putative positives as those individuals that are not in any negative pools.
5. Compute stage-two individual testing results for the putative positives: putative positive individual *j* tests positive if *z*_*j*_ > LOD and tests negative otherwise.
6. Declare individuals identified by HYPER as those that tested positive in stage-two.
7. Record the following for the current trial *r* and day *d*:
  - the number of true positive individuals identified by HYPER: 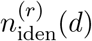,
  - the number of tests expended: *T* ^(*r*)^(*d*) = *m* + number of tests used in stage-two.
  - the number of true positive individuals seen: 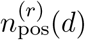 = number of individuals with viral load > 0,

For each day, we repeat this for 500 initial trials, then continue until either at least 2,500 true positive individuals have been seen or a total of 200,000 trials have elapsed (including the initial 500). This is to reduce experimental noise. Denoting *R* to be the total number of trials run, we then compute the following averages across trials

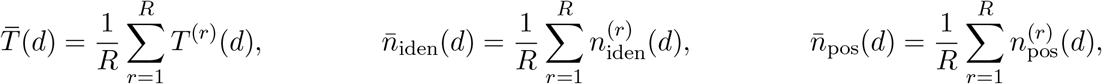

then finally compute the average efficiency gain and average sensitivity for day *d* as follows:

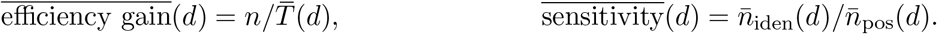

Note that step 2 in the simulation above captures dilution due to pooling, since each individual’s viral load gets divided by the pool size. The Poisson distribution models the “arrival” of viral particles when the small volume is pipetted from each swab. Note also that step 5 models the individual testing of stage two as having no false positives. Doing so simplifies the simulation without meaningfully affecting our conclusions (e.g., the most effective pooling designs, which do not depend substantially on stage two specificity). We do include false positives in stage one, since the overall efficiency depends on the specificity there. The parameters were chosen to match earlier modeling studies^11;75–77^.

For the 8 × 12 and 16 × 24 plate-based array designs^9^, the simulation proceeds in the same way except for step 2, where the corresponding array pools are used instead. Recall that the array method is a two-stage method like HYPER. For P-BEST^8^, which is a one-stage method, steps 1-3 are the same (except that step 2 now uses the P-BEST pools). Steps 4-6 are replaced by running the P-BEST decoder to identify individuals. For this, we followed the example (including its tuning parameters) provided online by the authors at https://github.com/NoamShental/PBEST/blob/f7ffebe6c7021ee40167239210806c5a1319f81e/mFiles/example_PBEST.m. Finally, since P-BEST has no second stage of validation tests, the number of tests expended is always *T* ^(*r*)^(*d*) = *m* = 48.

Fig. 2 plots the average efficiency gains and average sensitivities of the various methods for each day in a 90-day window of epidemic growth. Here we included individual testing, which has a constant average efficiency gain of 1 (unity) since it is the baseline. Its average sensitivity on day *d* is equal to

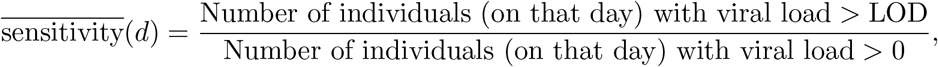

since individual testing identifies those individuals with viral load > LOD, and true positive individuals are those with viral load > 0 (as before). The average sensitivities of the various methods appeared to generally have significant experimental noise. So, Fig. 2 plots the raw averages (i.e., 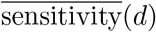 as dots along with a degree-8 polynomial curve fitted to 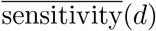 v.s. log_10_ *p*(*d*) across the plotting window of days *d* = 20, …, 110, where *p*(*d*) is the prevalence on day *d*.

In Fig. 2a, we compared HYPER designs H_96,16,2_ and H_384,32,2_ with their counterpart array designs and P-BEST. For the HYPER designs, the numbers *n* of individuals per batch were chosen to match the array designs and P-BEST. The numbers *m* of pools were chosen so that the corresponding pool sizes *nq/m* match the maximum pool sizes of the array designs (12 for the 8 × 12 array and 24 for the 16 × 24 array). Fig. 2b compares HYPER designs H_384,32,2_, H_384,16,2_, and H_384,12,2_ that have varying numbers of pools. Fig. 2c compares HYPER designs H_384,12,1_, H_384,12,2_, and H_384,12,3_ that have varying numbers of splits.

### Comparison of pooling methods under resource constraints

We used the simulations above to evaluate the various methods (individual testing, HYPER, plate-based array designs, P-BEST) under resource constraints and over time. We considered two forms of resource constraints: i) a limited daily sample collection budget, and ii) a limited daily testing budget. We let both range from 12 to 6144, forming the grid of resource-constrained scenarios shown in Fig. 3g and Supplementary Fig. 10, with a few selected scenarios highlighted in Figs. 3a to 3f. These figures evaluate average performance of the various methods when deployed across days 40-90 of the simulation. Supplementary Figs. 11 to 16 repeat the analysis (using the same set of scenarios) for individual days, namely days 53, 80, 83, 84, 90, and 93. Hence, we will focus on describing Fig. 3 and Supplementary Fig. 10; Supplementary Figs. 11 to 16 are similar.

In each scenario, we evaluated each method by its *effective screening capacity*. As discussed in the main text, this performance metric measures how many individuals the method can screen under the resource constraints, with a correction applied to account for the associated sensitivity. To measure performance over time, we also consider averaging across a chosen set of days 𝒟. Fig. 3 and Supplementary Fig. 10 consider days 40-90, so 𝒟 = {40, …, 90} there. Supplementary Figs. 11 to 16 examine individual days, which corresponds, e.g., to 𝒟 = {53} in Supplementary Fig. 11. To compute average effective screening capacity, we first determine the number of batches *b*(*d*) that can be run on each day *d*, and its corresponding average 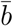:

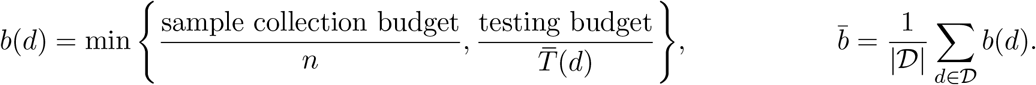

If 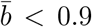 batches per day, i.e., fewer than 0.9 batches can be run per day on average, then the method is considered infeasible within the resource constraints. Then we set the method to have an average effective screening capacity of 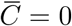. Setting the above threshold at 0.9 captures an assumed flexibility to use fewer or more tests across days. Otherwise, if 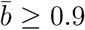, we compute the effective screening capacity *C*(*d*) for each day *d* and its corresponding average 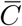 as follows:

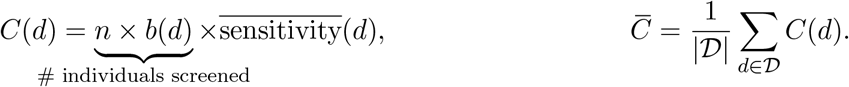

Figs. 3a to 3f show the average effective capacities for the considered methods as bars, with the corresponding average number of batches noted at the bottom of each bar. Multiple configurations are available for both the array method (the 8 × 12 and 16 × 24 array designs) and HYPER (various choices of *n, m* and *q*). For these methods, we select the most effective among all configurations, i.e., the configuration with the highest average effective screening capacity 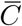. For HYPER, in particular, we optimized over the configurations listed in Supplementary Table 1. The chosen configuration is noted at the top of each bar in Figs. 3a to 3f.

Supplementary Fig. 10 shows the bar graphs for the full range of resource-constrained scenarios considered. Fig. 3g summarizes these findings by showing only which method was best (where we distinguish different choices of *q* in HYPER), the corresponding average effective screening capacity, and the corresponding configuration.

### HYPER pool designs from hypergraph factorization

As we illustrated in Fig. 1a, HYPER assigns individuals to pools by cycling through a sequence of pool assignments given by hypergraph factorization. Namely, for *q* = 2 splits and *m* = 6 pools labelled A-F, we cycled through the 15 possible pairs of pools in the order: AB, CD, EF, BC, DF, AE, BD, AF, CE, BE, CF, AD, BF, DE, AC. Namely, individual 1 was assigned to pools A and B, individual 2 to C and D, and so on; after individual 15, we return to the beginning of the sequence and cycle through again. For *n* = 18 individuals, this would result in the following pool assignments:

| Individual | 1 | 2 | 3 | 4 | 5 | 6 | 7 | 8 | 9 | 10 | 11 | 12 | 13 | 14 | 15 | 16 | 17 | 18 |
| --- | --- | --- | --- | --- | --- | --- | --- | --- | --- | --- | --- | --- | --- | --- | --- | --- | --- | --- |
| Pools | AB | CD | EF | BC | DF | AE | BD | AF | CE | BE | CF | AD | BF | DE | AC | AB | CD | EF |

For use in a lab protocol, it can sometimes be helpful to re-order these assignments so that individuals assigned to the same pair of pools appear consecutively:

| Individual | 1 | 2 | 3 | 4 | 5 | 6 | 7 | 8 | 9 | 10 | 11 | 12 | 13 | 14 | 15 | 16 | 17 | 18 |
| --- | --- | --- | --- | --- | --- | --- | --- | --- | --- | --- | --- | --- | --- | --- | --- | --- | --- | --- |
| Pools | AB | AB | CD | CD | EF | EF | BC | DF | AE | BD | AF | CE | BE | CF | AD | BF | DE | AC |

This is useful if we plan to first combine the samples from individuals 1 and 2, then split that combined sample into pools A and B. Likewise for individuals 3 and 4, as well as 5 and 6, in this case. To avoid confusion, we emphasize that we have simply re-ordered the pool assignments to show the repeated ones one after the other. Thus, the table does not show AB, CD and EF as the first three pairs, but rather AB is repeated twice, then CD is repeated twice, and so on.

### Hypergraph factorization

The sequence of pool assignments used in HYPER comes from hypergraph factorization. Here we describe the key ideas (and algorithms) for factorizations and the underlying theoretical results, in parallel with the application to group testing.

Suppose we are given a number *m* of vertices, which correspond to pools in group testing. We consider the *complete hypergraph* 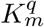 of order *q* ≤ *m* on these *m* vertices, which is simply the collection of all *q*-*hyperedges*, i.e., subsets of size *q* of the *m* vertices. For *q* = 2, this is the complete graph on *m* vertices, which can be drawn as all edges connecting *m* vertices.

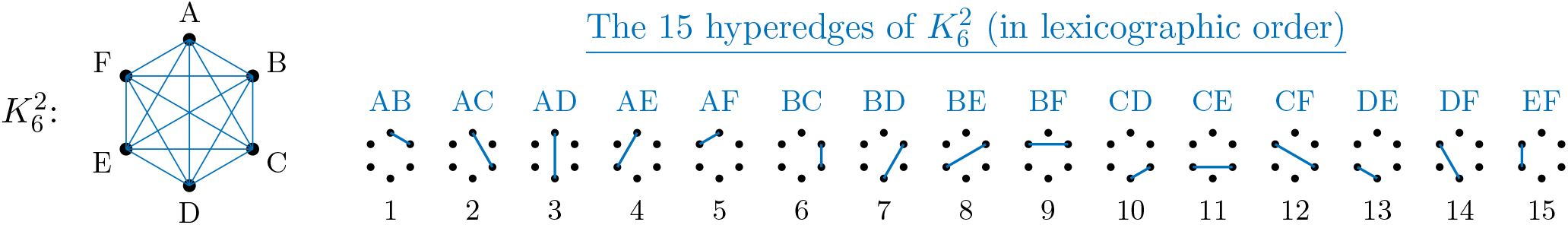

Drawing the corresponding complete hypergraph for *q* = 3 is harder, but one can quickly visualize the hyperedges as triangles connecting *q* = 3 vertices:

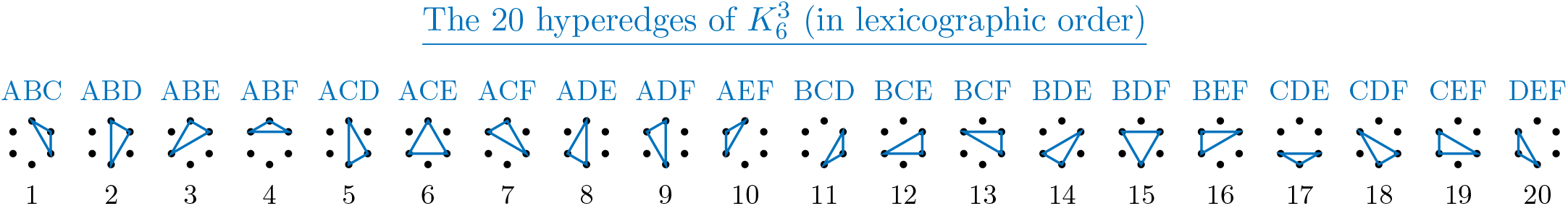

In general, there are 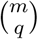 hyperedges in the complete hypergraph 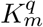, given by all subsets of size *q* of the *m* vertices. In group testing, the hyperedges correspond to samples: each sample is placed into the pools contained in the subset of pools determined by a hyperedge.

### Connection to group testing

For HYPER group testing, we are interested in assigning samples to pools in a simple and balanced manner. The notion of balance corresponds to using each pool an equal number of times; or as close as possible to this. This can be achieved quite directly in simpler cases, but requires more work in more complex cases. Consider now the simplest case, where *q*, the number of pools into which each sample is placed divides the total number of pools. Under this number-theoretic condition, for any *m* pools, we can clearly split them into *m/q* non-overlapping subsets/hyperedges of size *q*, and thus for a set of *m/q* samples, we can use each of the *m* pools exactly once, achieving perfect balance. The above partitioning hyperedges are called a 1-*factor* of the hypergraph. For example, for *q* = 2 and *q* = 3 (both with *m* = 6) we could have:

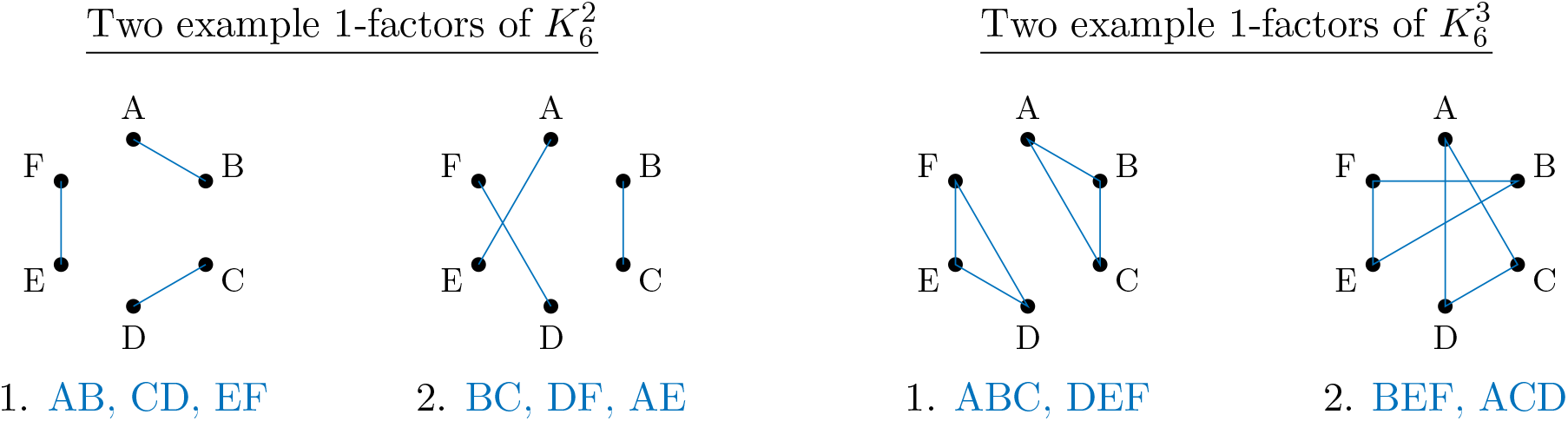

### Baranyai’s theorem

While this method to achieve balance is clear for *n* = *m/q* samples, it is less clear if and how something similar can be achieved for more samples. In fact, a celebrated result in combinatorics, *Baranyai’s theorem*^66^, states that the complete hypergraph can be *factorized*, in the sense that the collection of 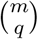 hyperedges can be split/partitioned into 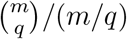 non-overlapping and different 1-factors (each of which containing *m/q* hyperedges), such that each *q*-hyperedge appears in exactly one of the partitions. For group testing, this means that perfect balance can be achieved whenever the number of splits *q* divides the total number of pools *m* (e.g., for a two-pool split, we need an even number of pools). In theory, this solves exactly the problem we need.

Baranyai’s theorem guarantees the existence of the desired designs, but does not provide efficient algorithms for constructing them. In fact, at the current time, general constructions seem to be known only for *q* = 2 and *q* = 3. For relatively small *q, m* one can certainly attempt to use brute-force enumeration to find appropriate designs. However, in this work we will focus on efficient and general constructions.

### Hypergraph factorization for *q* = 2

For *m* even, we use the following efficient method for constructing a factorization of 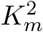. Here we follow the description in Section VII-5.5 of Colbourn and Dinitz^30^ and illustrate it using *m* = 6 as an example; see also page 595 of Beth et al.^28^. The construction begins with the following starter:

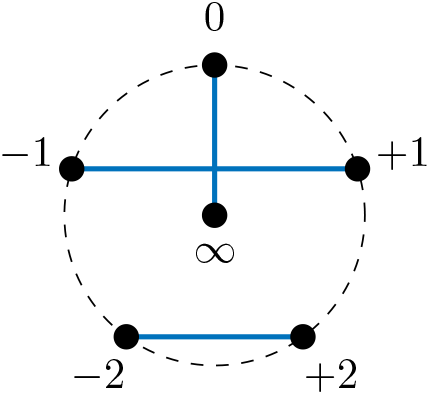

Namely, vertices *V* = {−*u*, …, +*u*, ∞} where *u* = *m/*2−1, i.e., ℤ_*m*−1_ ∪ {∞}, with the starter 1-factor formed by edges {(0, ∞), (−1, +1), …, (−*u*, +*u*)}. The remaining 1-factors are then generated by “rotating the diagram” as follows:

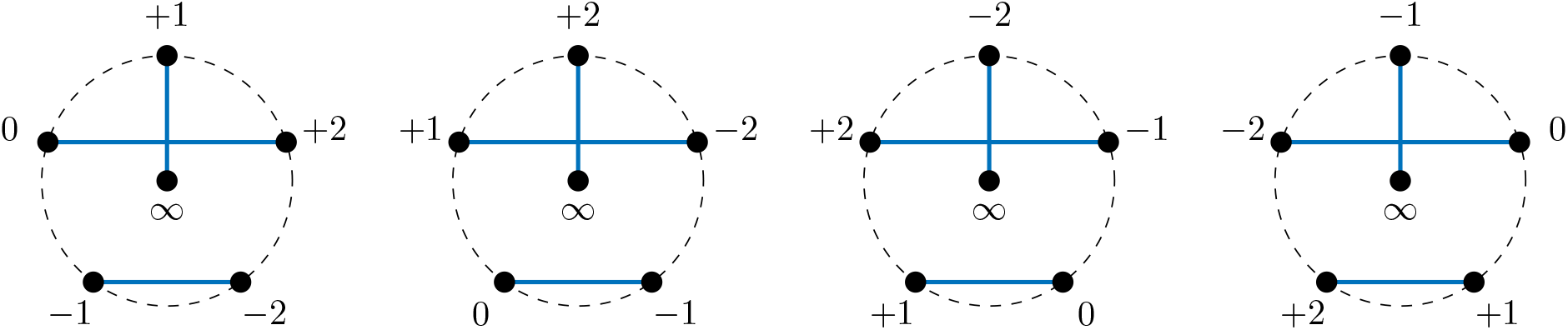

This yields *m* − 1 many 1-factors in total, that taken together form the desired hypergraph factorization of 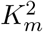. To connect this back to the pools, simply relabel the vertices *V* using the pool names. For example, using

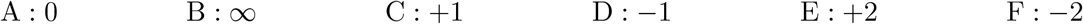

for the above yields the following 1-factors:

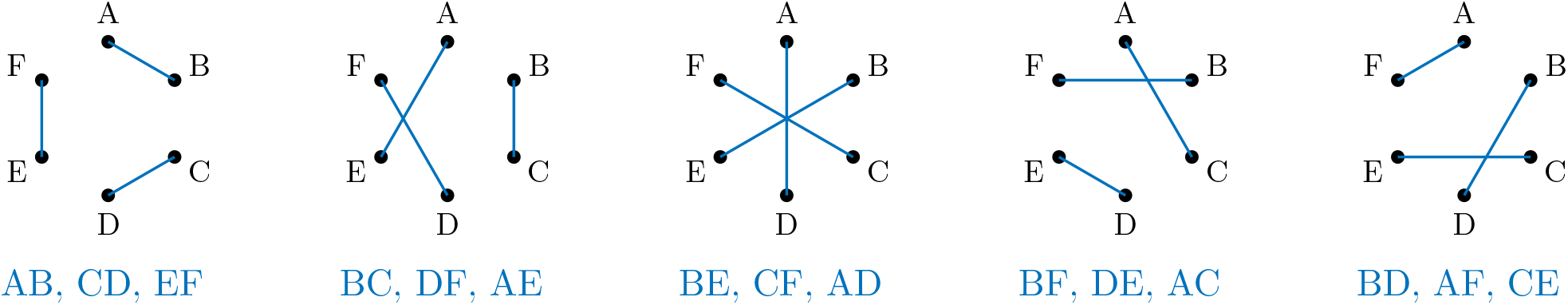

### Hypergraph factorization for *q* = 3 via Beth’s construction

We will leverage the non-trivial number-theoretic construction for *q* = 3 due to Thomas Beth^67;68^, which is guaranteed to work when *m* = 6*k* for some integer *k* (so the total number of pools is divisible by six), and *r* = 6*k* − 1 is a prime number (that is, a number that is not divisible by any other number between 1 and *r*). In what can be viewed as a lucky coincidence, it so happens that most of the designs that we are interested in enjoy these properties: e.g., for pool sizes of *m* = 6, 12, 24, 48, each are divisible by six, and further we have that *r* = 5, 11, 23, 47 are prime numbers.

#### Algebraic background

We follow the description in Beth^67^ (which appears to be difficult to access online; the construction is also presented in the thesis^68^, Section 3.1, and also referenced in Tamm^69^). The construction works as follows. Consider the finite field (or Galois field) of the prime order *r, GF* (*r*) = ℤ*/r*ℤ= *F*_*r*_. This is defined as the set of numbers {0, 1, …, *r* − 1}, with addition, multiplication, and division by nonzero elements all defined modulo *r* (i.e., the result is always the residue after dividing by *r*). To this field, we append the symbol ∞, as the result of division by zero, so that 1*/*∞ = 0. We also define *c* + ∞ = *c* · ∞ = ∞ for all *c* ≠ 0 ∈ *F*_*r*_. This constitutes the so-called “projective line” *PG*(1, *r*), with the “point” ∞ at infinity.

#### Beth’s construction

Now, Beth considers the fractional linear map *π* : *PG*(1, *r*) → *PG*(1, *r*) given by *π*(*x*) = −(1 + *x*)*/x*. Here, 1 denotes the additive unit of the field, while addition and division are taken modulo *r*. A key observation is that *π* is a fixed-point-free map of order three; that is, it maps *x* → *π*(*x*) → *π* ◦ *π*(*x*) → *π* ◦ *π* ◦ *π*(*x*) = *x*, such that all intermediate values are distinct. Thus, these *orbits* of *π* are sets of size three that partition *PG*(1, *r*). Let *O* = {*A*_*i*_, *i* = 1, …, (*r* + 1)*/*3} be the partition of *PG*(1, *r*) into orbits (and note that the size of *PG*(1, *r*) is *r* + 1, hence there are (*r* + 1)*/*3 orbits).

Let also *ω* be a primitive element of *F*_*r*_, that is an element such that *ω*^*j*^ ≠ 1, for any *j* = 1, 2, …, *r* − 2. Then, Beth’s result^67;68^ states that the partitions induced by multiplying and translating *O* by specific values *λ, g* as *λ* · *O* + *g* form a 1-factorization of *F*_*r*_. Here *λ* · *O* + *g* means that we take each of the hyperedges *A*_*i*_ ∈ *O*, and transform their elements affinely into *λ* · *A*_*i*_ + *g*, thus obtaining another hyperedge. Specifically, *λ* needs to take the values of the powers of *ω* given by *λ* = *ω*^*j*^, *j* = 1, …, (*r* − 1)*/*2, and *g* can take any value in *F*_*r*_.

The key for us is that this construction can be evaluated very efficiently, by simply iterating over the orbits of *π* and the values *λ, g*.

#### Example: *r* = 5

For *m* = 6 pools, *r* = *m* − 1 = 5 is a prime number and Beth’s construction applies. We go through the construction in this setting as an illustrative example.

1. For *r* = 5, we have *F*_*r*_ = {0, 1, 2, 3, 4} and *PG*(1, *r*) = {0, 1, 2, 3, 4, ∞}.
2. We compute orbits of *π* by repeatedly applying *π* as follows:

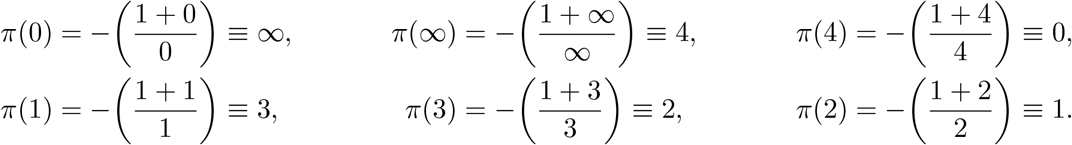 So we have the two orbits *O* = {{0, ∞, 4}, {1, 3, 2}} that partition *PG*(1, *r*).
3. We find a primitive element of *F*_*r*_ by looking at powers

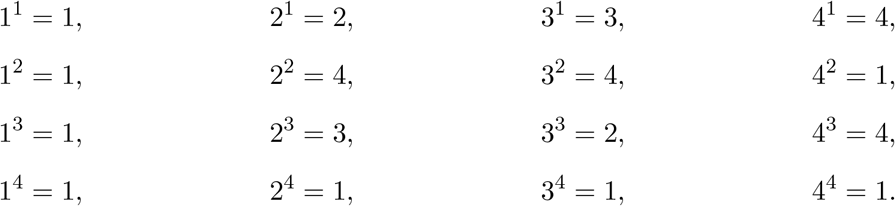 So we can choose *ω* ∈ {2, 3}. We will (arbitrarily) choose *ω* = 2.
4. Finally, we loop through *λ* and *g*. For *ω* = 2, *λ* loops through {2, 4}.

| | $\lambda = 2$ | $\lambda = 4$ |
| --- | --- | --- |
| $g = 0$ | $\{0, \infty, 3\}$<br>$\{2, 1, 4\}$ | $\{0, \infty, 1\}$<br>$\{4, 2, 3\}$ |
| $g = 1$ | $\{1, \infty, 4\}$<br>$\{3, 2, 0\}$ | $\{1, \infty, 2\}$<br>$\{0, 3, 4\}$ |
| $g = 2$ | $\{2, \infty, 0\}$<br>$\{4, 3, 1\}$ | $\{2, \infty, 3\}$<br>$\{1, 4, 0\}$ |
| $g = 3$ | $\{3, \infty, 1\}$<br>$\{0, 4, 2\}$ | $\{3, \infty, 4\}$<br>$\{2, 0, 1\}$ |
| $g = 4$ | $\{4, \infty, 2\}$<br>$\{1, 0, 3\}$ | $\{4, \infty, 0\}$<br>$\{3, 1, 2\}$ |

Note that each cell in the resulting output table forms a partition of $\{0, 1, 2, 3, 4, \infty\}$ as desired. Looping over the cells and using the partitions to form pools yields the desired design.

Note that each cell in the resulting output table forms a partition of {0, 1, 2, 3, 4, ∞} as desired. Looping over the cells and using the partitions to form pools yields the desired design.

### General background on design theory

To understand our designs, and how they fit into the broader context, it is valuable to introduce some basic concepts from design theory. See, e.g., Beth et al.^28;29^ for excellent introductions, and we will follow notation and terminology from those references.

For us, a design *I* is a collection of points *V* and blocks *B*, and an assignment of some points to some blocks. In group testing, the points correspond to samples, and the blocks correspond to pools. The terminology of points and blocks is meant to be evocative of geometry, and indeed designs are closely connected to finite geometries such as affine and projective planes. Intuitively, points can sometimes be viewed as geometric points, while blocks can be viewed as lines. Points will be denoted with lowercase letters such as *p*, while blocks will be denoted with upper case letters such as *B*. The fact that point *p* is associated with (or incident on) block *B* is denoted as *pIB*.

Designs are called *q*-hypergraphs, if the size of the set of points incident on each block *B* is *q*, i.e., (*B*) = {*p* ∈ *V* : *pIB*} has size *q*. For group testing, this means that each sample is assigned to *q* pools. We are thus interested in *q*-hypergraphs, for small values of *q*, such as 2, 3.

A partition of a design is a disjoint union of its blocks into parts. A parallel class of a design is a collection of blocks such that each point is incident on exactly one block. This is analogous of the geometric idea that parallel lines do not intersect, and so if we partition the space into parallel lines, then each point belongs to exactly one such line. A design is called resolvable if it has a partition into parallel classes (a.k.a 1-factorization, parallelism or resolution).

For group testing, a resolution means that in each part of the partition of the blocks, we use each pool exactly once. Since our goal is to use pools in a balanced way, this is precisely what we want. Thus, from a design theory perspective, we are interested exactly in resolutions of *q*-hypergraphs. There is a great amount of work on existence and constructions of such resolutions, see, e.g., Ch VIII of Beth et al.^28^ and references therein.

One classical strategy is the permutation group action approach. Here, we start with a collection {*B*_*j*_}, *j* = 1, …, *J* of base blocks (say of size *q*), viewed as subsets of the set of pools [*m*] = {1, …, *m*} where *m* is the number of pools, and a subgroup *G* of the permutation group *S*_*m*_ on *m* elements. If the action of *G* on *B*_*j*_ ⊂ [*m*] *j* = 1, …, *J* leads to *J* non-overlapping orbits, then there are classical conditions under which the collection of these orbits forms a *t*-design (i.e., a hypergraph such that all *t*-subsets of points are incident on the same number of blocks), see Theorem III.8.2 on p. 207 of Beth et al.^28^. A more specific technique is the difference cycle method, which is an application of the permutation group action method when the cyclic group *G* = ℤ_*l*_ acts on the vertices *V* = ℤ_*l*_ by translation. This approach leads to resolutions of the complete hypergraph for both *q* = 2 and *q* = 3, see Section VIII.8 of Beth et al.^28^. These are precisely the algorithms that we use.

### Performance characterization under a common statistical model

Here we study the performance of HYPER under the common statistical model where each individual is positive independently at random with probability *p* and each test may be incorrect with some probability, i.e., the tests may be noisy. In particular, we suppose each test has a specificity of 1 − *α* and a sensitivity of *β*.

### An upper bound on the expected number of tests in the noiseless case

We first consider the noiseless case (*α* = 0, *β* = 1), and give an upper bound on the expected number of tests used by HYPER (including stage two tests). To get this result, we leverage the Dawson-Sankoff inequality^70^, a nontrivial refinement of the Bonferroni union-intersection inequalities, which we use in the form given by Galambos^71^. We will later relax the assumption made here that tests have perfect sensitivity and specificity.

#### Theorem 1.

*Consider HYPER with any number of samples n, number of pools m, number of pools per sample q, such that n is a multiple of m/q, and let k* = *nq/m be the pool size. Let p be the prevalence level, and suppose that each sample is positive independently with probability p. Let T be the number of tests used by HYPER (including stage two tests), which is a random variable. For any positive integer l* ≥ 2, *with* 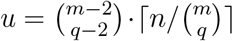, *the expected number of tests is upper bounded by*

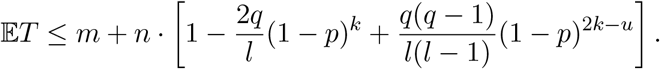

*The bound becomes an equality when (A) q* = 1, *(B)*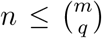 *and q* = 2; *and with taking l* = 2 *in both cases, or when (C) n is a multiple of* 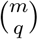. *For general q, the optimal choice for the parameter l is bounded by*

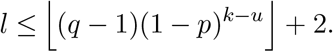

*Proof of Theorem 1*. Let *R*_*i*_, *i* = 1, …, *n* be the indicator of the event that we need to re-test individual/sample *i*. Using the standard approach of calculating 𝔼*T*, we find that the number of tests required is equal to *m* (one for each pool), plus any retests required. Hence,

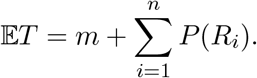

Now, for our decoder, *R*_*i*_ happens precisely when all groups containing *i* are positive. Let *T*_*i*_, *i* = 1, …, *n* be the indicator that the *i*th sample is positive. Let *G*_*j*_ ⊂ {1, …, *n*} be the samples contained in pool *j*, for *j* = 1, …, *m*.

We use a refined version of the Bonferroni inequality known as the Dawson-Sankoff inequality^70^ to bound *P* (*R*_*i*_). First let us recall the familiar Bonferroni union-intersection inequalities. Consider events *A*_1_, …, *A*_*N*_, and for all *j* ∈ {1, …, *N*}, define

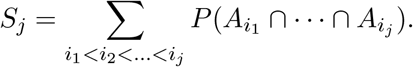

Then, the well-known Bonferroni inequalities state that for even *h* ∈ {1, …, *N*},

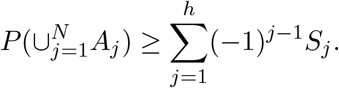

By construction, there are *q* pools containing sample *i*. Without loss of generality, we can assume by relabeling the pools that their indices are 1, …, *q*. Then, the samples contained in them are *G*_1_, …, *G*_*q*_; which implicitly depend on *i*, but this is not displayed for notational simplicity. In the Bonferroni inequality, we set *N* = *q*, and for *j* = 1, …, *q*,

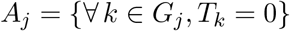

be the event that all samples contained in the *j*-th pool containing sample *i* test negative. As above, these implicitly depend on *i*, but this is not displayed for notational simplicity. By definition, the *i*-th sample is not retested, so *R*_*i*_ does not happen, precisely when none of the samples contained in the pools 1, …, *q* to which sample *i* belongs to are positive. Equivalently, 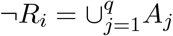. Then for any even integer *h*

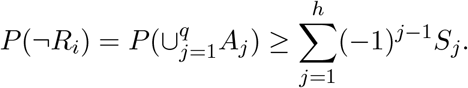

Taking *h* = 2, we thus find

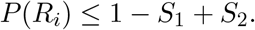

*P* (*R*_*i*_) ≤ 1 − *S*_1_ + *S*_2_.

We can get sharper results with the Dawson-Sankoff inequality^70^. In the form given by Galambos^71^, this states that for any integer *l* ≥ 2,

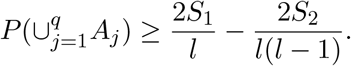

Now we can write 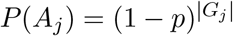 and for *j* ≠ *j*^*′*^, we have 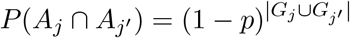.

It remains to bound |*G*_*j*_|. Here hypergraph factorization designs are useful, because they try to balance |*G*_*j*_|. In each consecutive block of *m/q* samples, they use each of the *m* pools exactly once. Thus, in each *G*_*j*_, there is at most one new sample. Recall that *k* is the number of consecutive blocks of samples of size *m/q*, and we assumed that *k* = *nq/m* is an integer. Based on the above, we have |*G*_*j*_| = *k*.

Moreover, |*G*_*j*_ ∪ *G*_*j′*_ | = |*G*_*j*_| + |*G*_*j′*_ | − |*G*_*j*_ ∩ *G*_*j′*_ |, and the intersection *G*_*j*_ ∩ *G*_*j′*_ has size at most 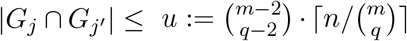. The reason is that, since hypergraph designs are maximally balanced, they only intersect at most 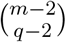 times in every consecutive block of 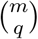 samples. These intersections correspond to the number of ways of choosing the remaining *q* − 2 pools out of the remaining *m* − 2. Hence, *P* (*A*_*j*_) = (1 − *p*)^*k*^ and for *j* ≠ *j*^′^, *P* (*A*_*j*_ ∩ *A*_*j′*_) ≤ (1 − *p*)^2*k*−*u*^. Thus, *S*_1_ ≥ *q*(1 − *p*)^*k*^ and 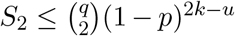. In addition, we have equality for *S*_2_ when either (A) *q* = 1, in which case the intersection is empty and *u* = 0, or 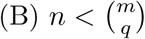 and *q* = 2 (in which case we know that the groups intersect in exactly *u* = 1 sample, which is the sample defining them) or (C) *n* is a multiple of 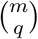 (in which case we know that the number of intersections is exactly given by *u* for each pair of groups). This leads to the desired result.

As is known^70;71^, the optimal choice for *l* is *l* = ⌊2*S*_2_*/S*_1_⌋ + 2. We can approximate the optimal choice using the calculations from the proof as

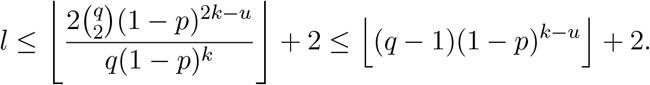

This finishes the proof.□

Let us denote *r* = 1 − *p*. When 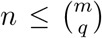, an optimal hypergraph design is obtained by minimizing the number of per-person tests *E*(*m, q*) = 𝔼*T/n*, i.e.,

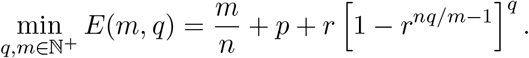

This formula is exact for *q* = 1 and *q* = 2. One can check that we recover the bound from above with equality for these cases when 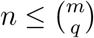 (so *u* = 0 for *q* = 1 and *u* = 1 for *q* = 2), and by taking *l* = 2. In more detail, we can write (replacing above *k* = *nq/m*, and using *r* = 1 − *p*)

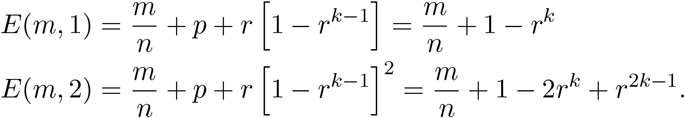

Moreover, by substituting into Theorem 1 *u* = 0 for *q* = 1 and *u* = 1 for *q* = 2, and by taking *l* = 2; the bounds given there for 𝔼*T/n* (denote them 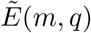) become

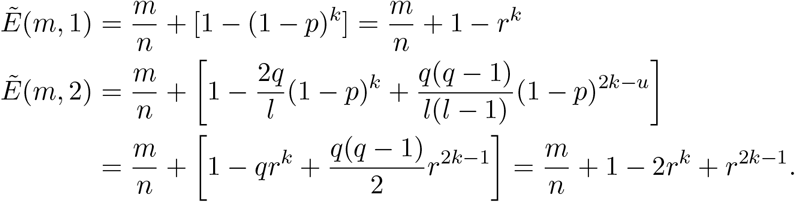

This shows that the upper bounds are sharp for *q* = 1, 2.

However, for a larger *q*, this formula corresponds to the so-called *locally tree-like* approximation in the graphical model corresponding to the observation model, when computing the probability of *R*_*i*_.

### Optimal efficiency of HYPER for *q* = 2 in the noiseless diminishing prevalence case

Our next result characterizes the optimal efficiency of hypergraph designs for *q* = 2 for diminishing *p* → 0.

#### Proposition 2

(Optimal efficiency of HYPER for *q* = 2)

*In the noiseless large n case (i*.*e*., *α* = 0, *β* = 1, *and n* → ∞*), the optimal efficiency of HYPER with q* = 2 *is approximately E*^*^ ≈ 3*p*^2*/*3^ *in the limit p* → 0 *with m/n* ≈ 2*p*^2*/*3^ − *p*.

These results are valid in the regime of *p* where 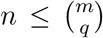, which restricts *p* to be larger than a certain threshold. When *p* → 0, we expect the optimal *m* to decrease in this limit, eventually leading to 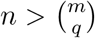. In practice, this means that the formula is valid for a larger range of *p* when *n* is larger.

In comparison, the optimal efficiency for Dorfman testing (*q* = 1) is approximately 2*p*^1*/*2^ for small *p* (see, e.g., Finucan^21^). Thus hypergraph designs improve over Dorfman designs. The same asymptotic efficiency *E*^*^ ≈ 3*p*^2*/*3^ is also achieved by three-stage-testing^21^ as well as double-pooling^64^. However, our proposal is a two-stage deterministic approach. In comparison, both the hierarchical testing approaches proposed in Finucan^21^ and Mutesa et al.^10^ attain an asymptotic efficiency *E*^*^ ≈ *ep* ln(1*/p*) as *p* → 0, which is asymptotically more efficient; but it requires multi-stage tests that we avoid.

More generally, we note that there is a lot of work on optimality of group testing, for various cases, e.g., two-stage and multi-stage algorithms, adaptive and non-adaptive algorithms, in worst case or average case, etc^48–53^. As *p* → 0, these works and others discuss a universal lower bound of order Θ(*p* ln(1*/p*)) on the efficiency. However, the best efficiency (and the algorithms that achieve it) depends on the specific rate at which *p* → 0. In particular, under the same statistical model as in our paper, Mezard and Toninelli^51^ construct certain tests where each sample is placed into *q* = ⌈ln(1*/p*)*/* ln 2⌉ pools, and show that these attain asymptotic efficiency *p* ln(1*/p*). Coja-Oghlan et al.^54^ constructs a 2-stage algorithm with asymptotically optimal efficiency, requiring *q* = *m* ln(2)*/*(*np*). Gebhard et al.^55^ discusses similar proposals for the noisy case. In our work, the constraints we work with do not allow *q* to grow with *p* → 0. Scarlett^56^ proposes a 4-stage algorithm with asymptotically optimal efficiency. Our work is also related to *d*-disjunctive superimposed matrices for pooled testing, which work without errors when the number of positives is at most *d* and there are no false positives^32;33^. Sharp bounds on the size of *d*-disjunctive matrices were constructed in Erdös et al.^31^. Classic error-correcting codes such as Reed-Solomon codes were suggested for group testing dating back at least to Kautz and Singleton^32^.

*Proof of Proposition 2*. Taking the limit as *n* → ∞ with *m/n* = *y* fixed, where *α* = 0 and *β* = 1, we obtain

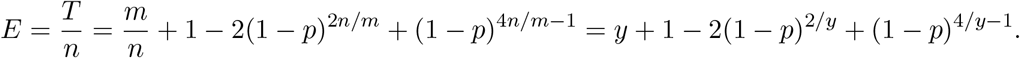

To optimize, we differentiate *E* with respect to *y*, obtaining

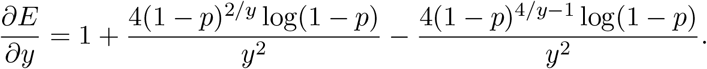

The optimal *y* can be obtained by solving 0 = *∂E/∂y* for *y* in terms of *p*. We approximate this solution in the limit *p* → 0 by taking the leading two terms of the Puiseux expansion (around *p* = 0) of the degree four Taylor approximation of *∂E/∂y* (with respect to *p* = 0). This has one branch corresponding to a real solution yielding *y*^⋆^ ≈ 2*p*^2*/*3^ − *p*.

Substituting *y* = 2*p*^2*/*3^ − *p* into *E* and computing a Taylor approximation yields

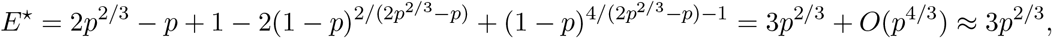

completing the derivation.□

### Generalization to the noisy case

We next study the noisy case, where each of the pooled tests can have false positives and false negatives. Our first result gives an exact formula for the expected number of tests for *q* = 1, 2, and moreover, also gives formulas for the false positive and false negative rates for each individual test.

#### Theorem 3

(Performance of hypergraph factorization: noisy case)

*Consider hypergraph factorization designs in a noisy observation model. Suppose n is a multiple of m/q. Suppose q* = 1 *or q* = 2, *and* 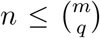. *Let k* = *nq/m and r* = 1 − *p. The expected number of tests has the following exact form:*

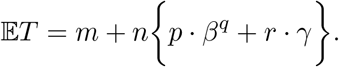

*Here γ* = [*β* + (*α* − *β*) · *r*^*k*−1^]^*q*^. *Denoting the odds ratio as o* = (1 − *p*)*/p the true negative and true positive probabilities for each individual sample’s status T*_*i*_ *and the test results* 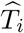 *are, respectively*,

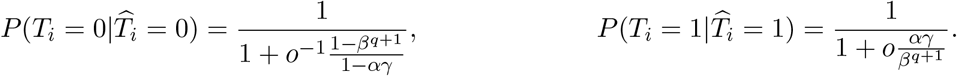

Our second result gives a more generally applicable upper bound on the expected number of tests, that is valid for all *q*.

#### Theorem 4

(Performance bound for hypergraph factorization: noisy case, upper bound)

*Consider hypergraph factorization designs in a noisy observation model with any number of samples n, number of pools m, number of pools per sample q, such that n is a multiple of m/q, and let k* = *nq/m*. *Let* 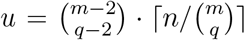. *Let p be the prevalence level, and suppose that each sample is positive independently with probability p. Suppose the pools and the re-tests have sensitivity* 1 − *α (where α is the level of each test), and specificity (or power) β. For any positive integer l* ≥ 2, *the expected number of tests is upper bounded by*

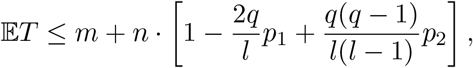

*Where*

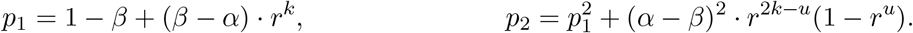

*Here r* = 1 − *p. The optimal choice of l, minimizing the upper bound, is achieved by l* = ⌊ (*q* − 1)*p*_2_*/p*_1_⌋ + 2. *Proof of Theorems 3 and 4*. We will follow, to some extent, the notation and assumptions from Bilder’s works (see, e.g., Bilder^22^). Let *Ĥ*_*j*_ be the binary result of testing group *j*, and *H*_*j*_ be the true status of the *j*-th group. By definition, 1 − *α* is the sensitivity of each grouped test, i.e., 1 − *α* = *P* (*Ĥ*_*j*_ = 0|*H*_*j*_ = 0) (we use this notation in convention with the notion of *α* for the level of a test in hypothesis testing); and *β* is the specificity (or power) of each grouped test, i.e., *β* = *P* (*Ĥ*_*j*_ = 1|*H*_*j*_ = 1). Moreover, we assumed that each test outcome is independent. We have

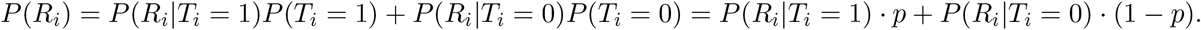

The key is to determine the probabilities *P* (*R*_*i*_|*T*_*i*_ = *t*). Now *R*_*i*_ happens if and only if each of the groups containing *i* have a positive status, i.e., for all *i* ∈ *G*_*j*_, we have *Ĥ*_*j*_ = 1:

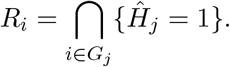

Since *q* = 1, 2, and 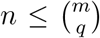, these groups are non-overlapping outside of *i*, and thus, their probabilities are independent conditional on *i*. We can thus write

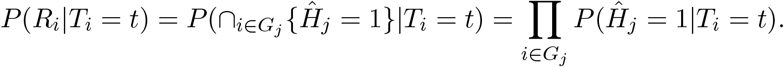

Next, we can condition on *H*_*j*_ for each term, to write

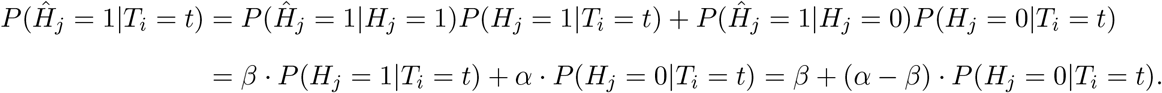

Moreover, since 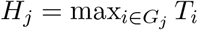, we have

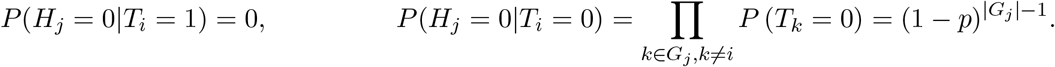

Working our way back up, we can substitute these above to find

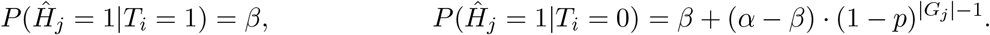

Letting *n*_*i*_ = |{*i* : *i* ∈ *G*_*j*_}| be the number of groups that *i* belongs to, we find

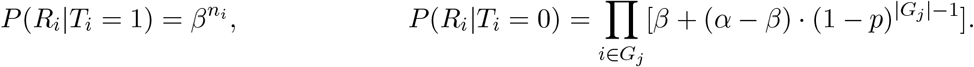

Finally,

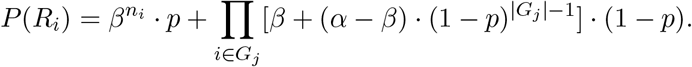

Hence

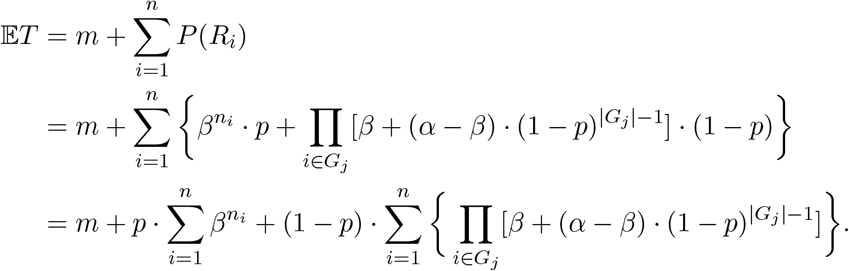

Recall that for the hypergraph designs, |*G*_*j*_| ≤ ⌈*n/*(*m/q*) ⌉, and if *n* is an integer multiple of *m/q* as assumed here, then |*G*_*j*_| = *nq/m*. Moreover, by construction, *n*_*i*_ = *q*. Hence we find

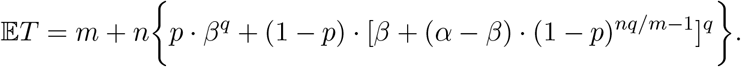

This gives the desired formula for the expected number of tests. Next we derive the per-test false negative and positive rates. Let 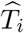 be the indicator that the *i*-th sample is declared positive. This happens if all groups containing *i* are positive in the first round, and then the result of a second independent test 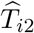 is also positive. Recall that *T*_*i*_ is the indicator that the *i*th sample is positive, and *P* (*T*_*i*_ = 1) = *p*.

We are interested in the probabilities 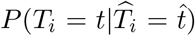, for 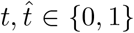. For 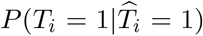 this denotes the true positive probability; while 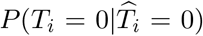 this denotes the true negative probability. Using Bayes’ rule, we can write

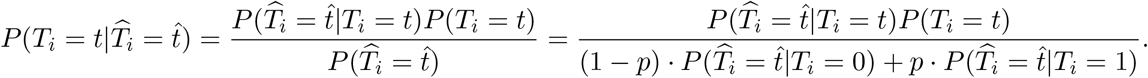

Thus the probabilities reduce to determining 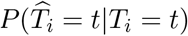. We can write, assuming the result of the re-test is independent of the original tests, and assuming the re-test has the same operating characteristics as any grouped test, that

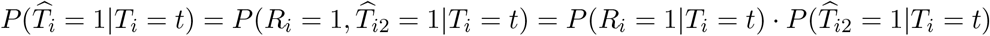

We have 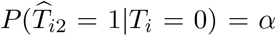 and 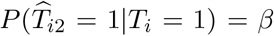. Hence, using our previous results and denoting 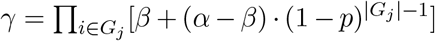, we have 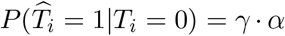 and 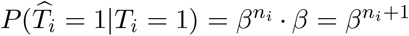. Working our way back,

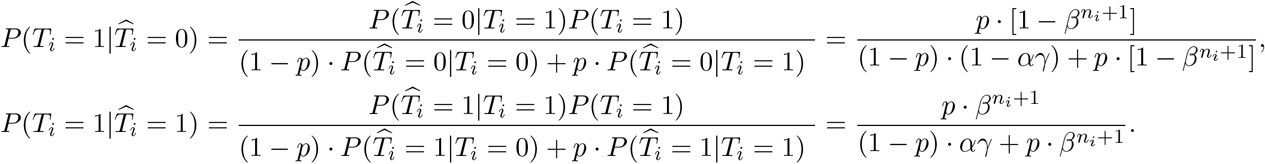

Recall that for the hypergraph designs, if *n* is an integer multiple of *m/q*, then *γ* = [*β* +(*α* − *β*) · (1 − *p*)^*nq/m*−1^]^*q*^ and *n*_*i*_ = *q*. Hence, denoting the odds ratio as *o* = (1 − *p*)*/p* we find that the true negative and true positive probabilities are, respectively,

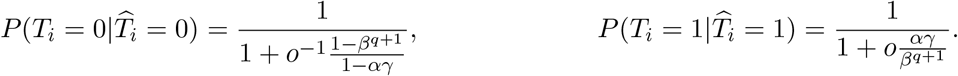

This finishes the proof of Theorem 4. Now, we proceed to Theorem 3. This follows in a similar way to the previous Theorem 1, but with more involved calculations. As before, the Dawson-Sankoff inequality^70^, in the form given by Galambos^71^, states that for any integer *l*,

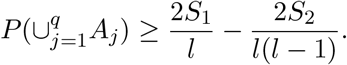

Taking *A*_*j*_ = {*Ĥ*_*j*_ = 0}, we find that the above equals 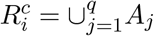. Now *S*_1_ = ∑*P* (*A*_*j*_), *S*_2_ = ∑_*j<g*_ *P* (*A*_*j*_ ∩*A*_*g*_). Thus it is enough to give a lower bound for *P* (*Ĥ*_*j*_ = 0) and an upper bound for *P* (*Ĥ*_*j*_ = *Ĥ*_*g*_ = 0) for all *j ≠ g*.

We can condition on *H*_*j*_ to write

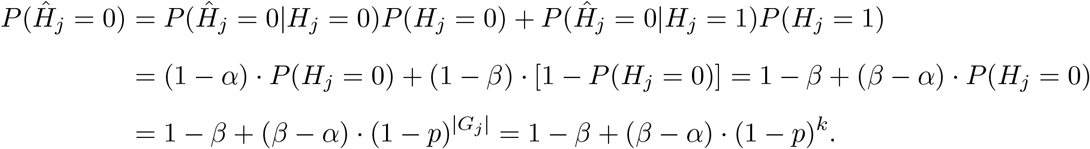

In the last line, we have used that *n* is a multiple of *m/q*.

Similarly, we can calculate for *j* ≠ *k*, noting that |*G*_*j*_| = |*G*_*g*_| = *k*, and denoting |*G*_*j*_ ∩ *G*_*g*_| = *v*, so that |*G*_*j*_ ∪ *G*_*g*_| = 2*k* − *v*,

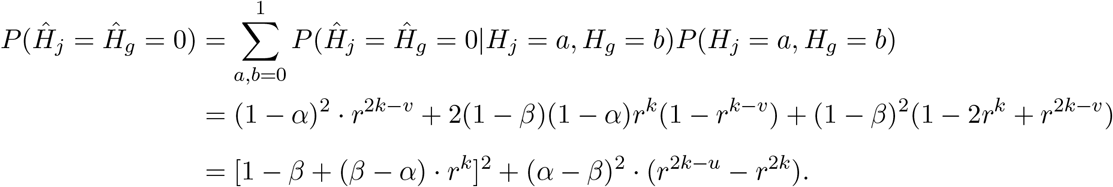

As discussed before, due to the construction of hypergraph factorization designs, we have 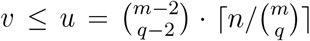. Since the above expression is monotonically increasing in *u*, we can also conclude that we can upper bound it by replacing *v* with *u*. This finishes the proof. □

---

**Supplementary Figure 1:**
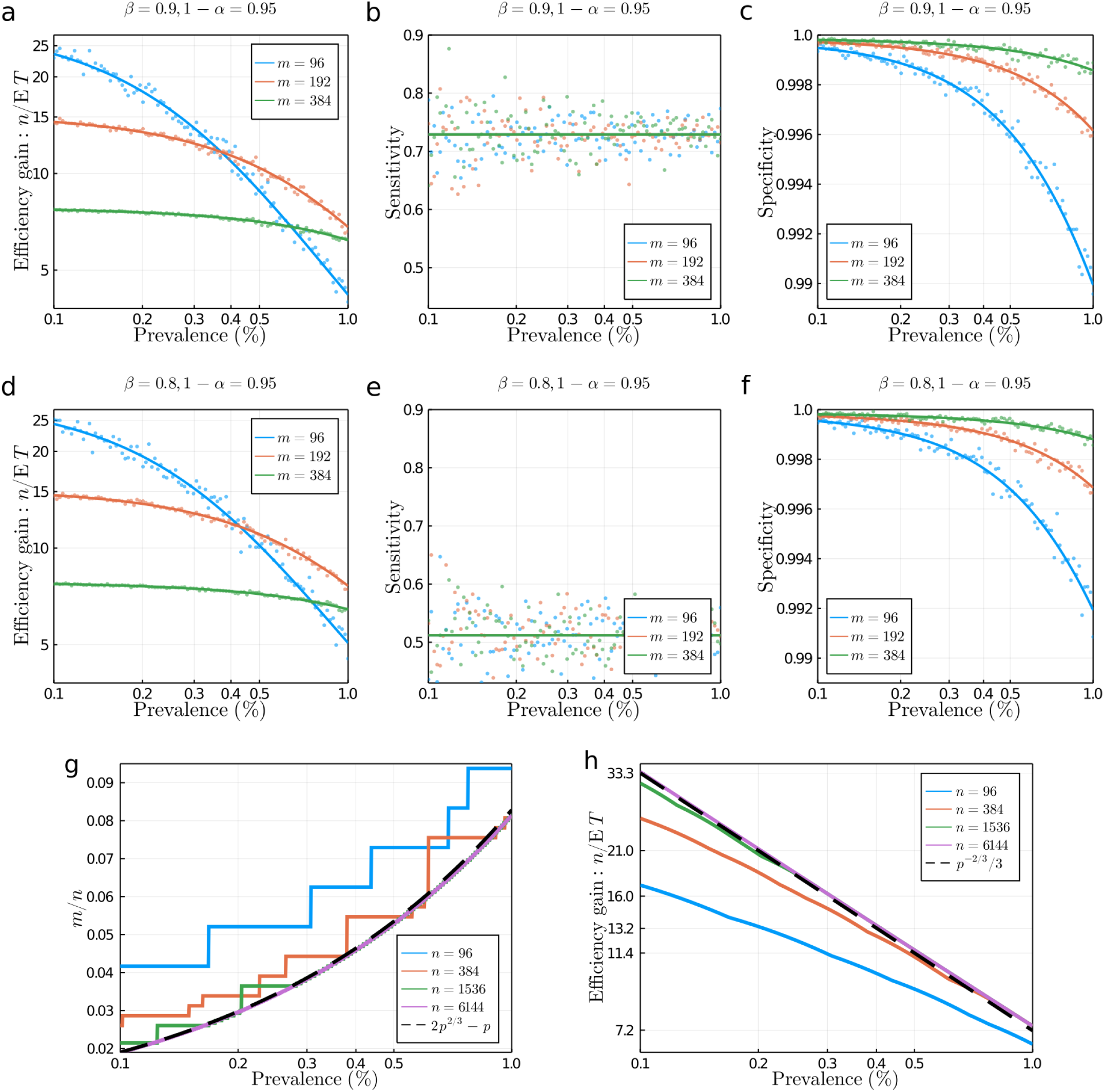
Performance under a common statistical model. We consider the overall efficiency (**a**), sensitivity (**b**), and specificity (**c**) of three HYPER designs (three choices of *m*, all with *n* = 3072 and *q* = 2) under a general statistical model with noisy tests having sensitivity *β* = 90% and specificity 1 − *α* = 95%. The theoretical predictions (solid curves) agree well with simulation results (scatter plots). For a lower sensitivity *β* = 80% (but same specificity 1 − *α* = 95%), the efficiency (**d**) is slightly better (since more positives are missed), the sensitivity (**e**) is lower, and the specificity (**f**) is slightly improved (at larger prevalence). For noiseless tests (*β* = 1 − *α* = 1), we study the optimal choice (**g**) of *m* for HYPER with *q* = 2 in the limit of large batches (*n* → ∞) with diminishing prevalence (*p* → 0). As *n* grows from *n* = 96 to *n* = 6144, the value of *m* optimizing the efficiency (found by exhaustive search) approaches the theoretical limit of *m/n* ≈ 2*p*^2*/*3^ − *p* for small prevalence *p*. Likewise, the corresponding optimal efficiency (**h**) for HYPER with *q* = 2 approaches 𝔼*T/n* ≈ 3*p*^2*/*3^ for small prevalence *p*. The approximation improves for increasingly small *p* as *n* grows.

**Supplementary Figure 2:**
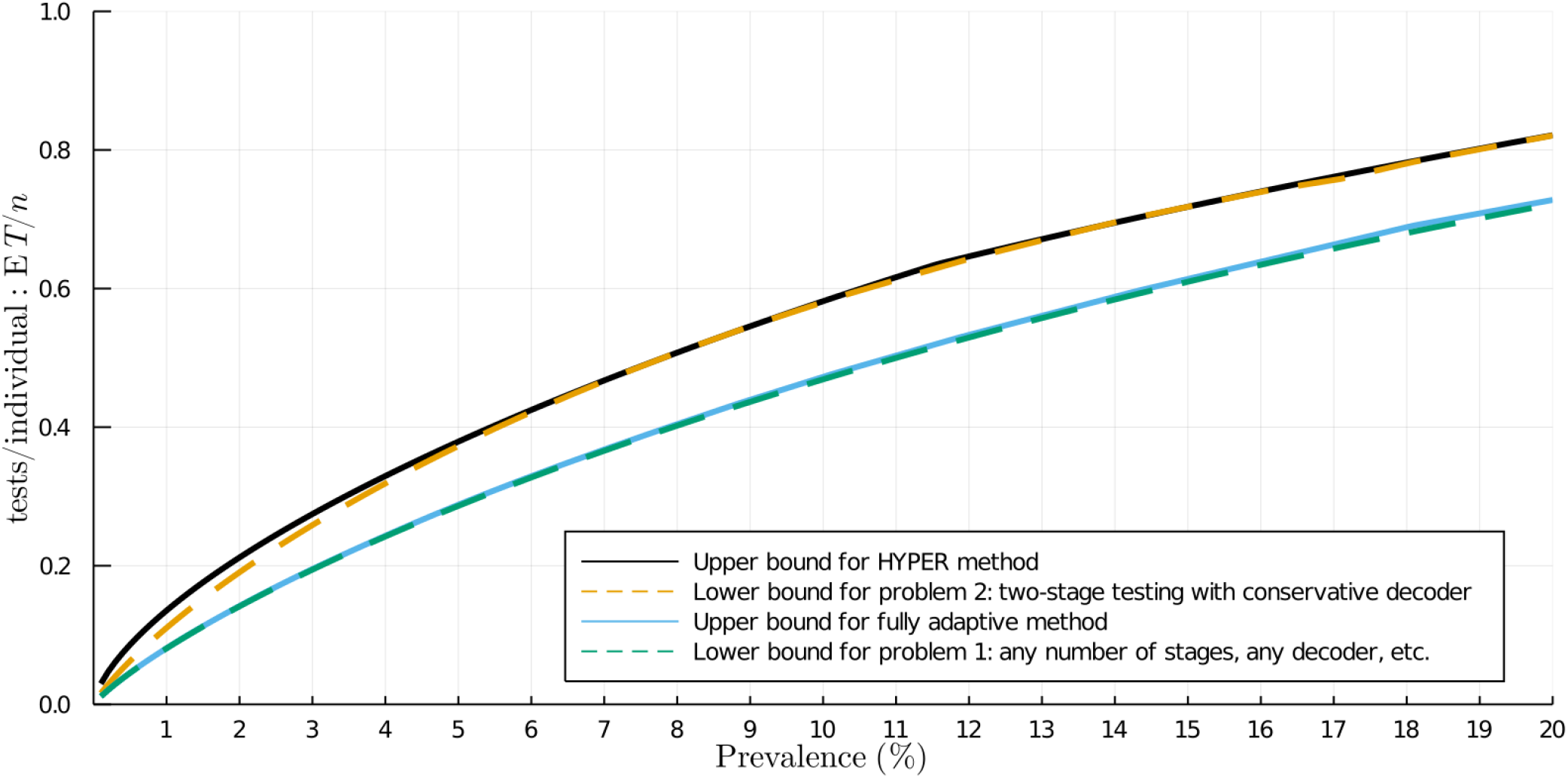
Comparison of two design problems and their bounds. We compare two design problems. Problem 1 is to minimize the average number of tests used per individual by using any group testing method (i.e., any number of stages, any decoder, etc.). This problem is tackled, e.g., by fully-adaptive methods. Problem 2 is to minimize the average number of tests used per individual but by using two-stage group testing methods with a conservative decoder^37^. This is the problem that HYPER tackles. Namely, we consider methods with a first stage of pooled tests followed by a second stage where putative positives are tested individually. This class of methods is of great practical importance. Using only two stages reduces the time needed to receive results, which is crucial in public health settings. Moreover, carrying out more stages can be difficult in practice due to the added logistical burden (especially for adaptively chosen pooled tests). Using a conservative decoder also helps in practice since it avoids more complicated reasoning, e.g., to identify definite positives by process of elimination. This figure compares known lower bounds for the two problems in the noiseless large *n* setting (i.e., *α* = 0, *β* = 1, and *n → ∞*) for a range of fixed prevalences *p* (sometimes called the *linear regime*^36^). As one would naturally expect, the lower bound (counting bound^36^) for problem 1 is lower than the lower bound^37^ for problem 2 since it is less constrained. For problem 1, fully-adaptive methods are nearly optimal, as was already known^36^. For problem 2, HYPER appears to be fairly close to optimal. The gap visible for smaller prevalences is likely due to: a) our additional constraint that *q* ≤ 3 (to aid real-life implementation), b) looseness in our upper bound for HYPER, and c) potential looseness in the lower bound^37^.

**Supplementary Figure 3:**
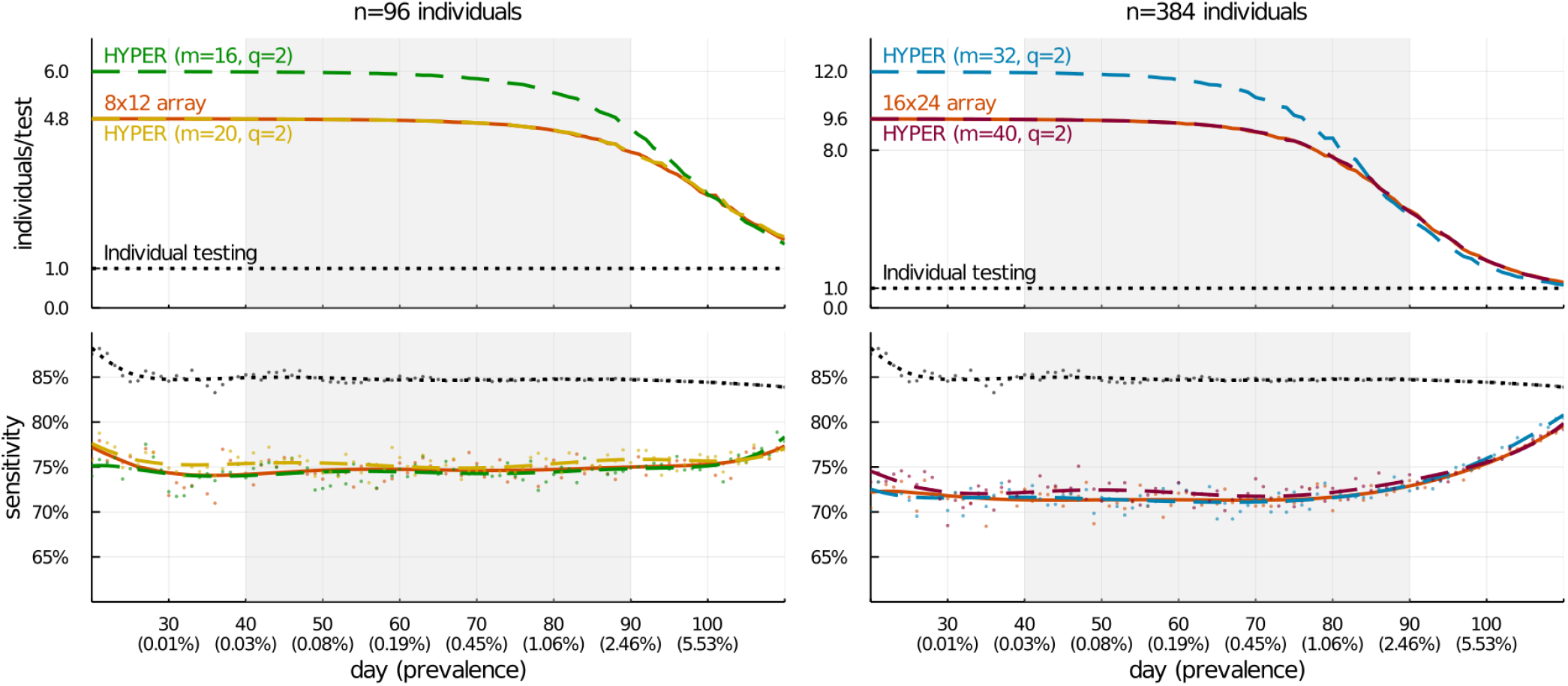
Comparison of plate-based array designs with additional HYPER designs. We expand the comparison of HYPER and array designs in Fig. 2. Previously, the HYPER designs were chosen to have the same maximum pool sizes (*nq/m* = 12 for H_96,16,2_; *nq/m* = 24 for H_384,16,2_) as the array designs. Here we also include HYPER designs that are instead chosen to have the same number of pools (*m* = 20 for the 8 × 12 array and *m* = 40 for the 16 × 24 array). These HYPER designs have efficiency nearly the same as their array counterparts. However, they have a slightly higher sensitivity, likely due to their more balanced pools.

**Supplementary Figure 4:**
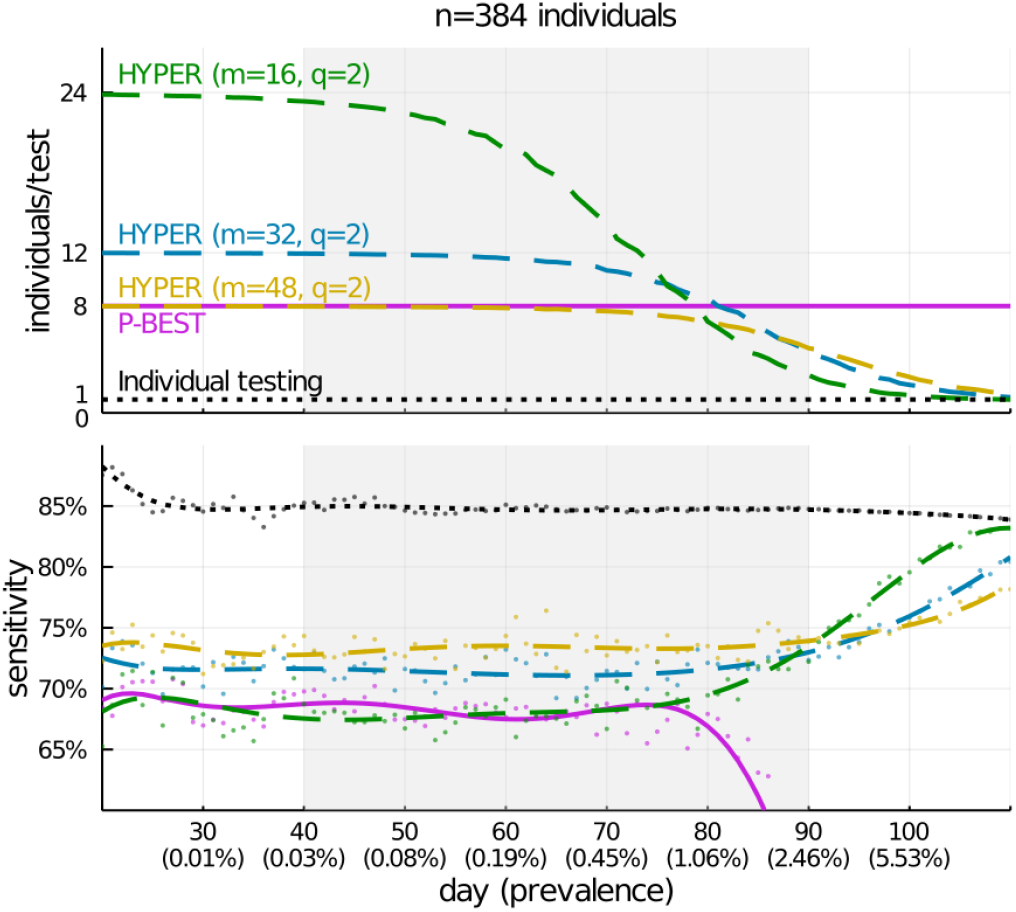
Comparison of P-BEST with additional HYPER designs. We expand the comparison of HYPER and P-BEST in Fig. 2. To the previous H_384,32,2_ HYPER design, we add a H_384,48,2_ design (with the same number of pools *m* = 48 as P-BEST) and a H_384,16,2_ design (with same pool size *nq/m* = 48 as P-BEST). Since H_384,48,2_ has the same number of pools as P-BEST, its efficiency at low prevalence is similar. However, its pools are one-third in size helping it achieve a higher sensitivity. The H_384,16,2_ has the same size pools as P-BEST, and a comparable sensitivity for much of the 50-day window highlighted. However, it has one-third as many pools giving it an initial efficiency roughly three times higher. As before (Fig. 2), the efficiency of HYPER declines for both designs as prevalence grows, eventually falling below the constant efficiency gain achieved by P-BEST around day 80. Likewise, as before, the sensitivity of HYPER grows around the same time, while P-BEST significantly loses sensitivity.

**Supplementary Figure 5:**
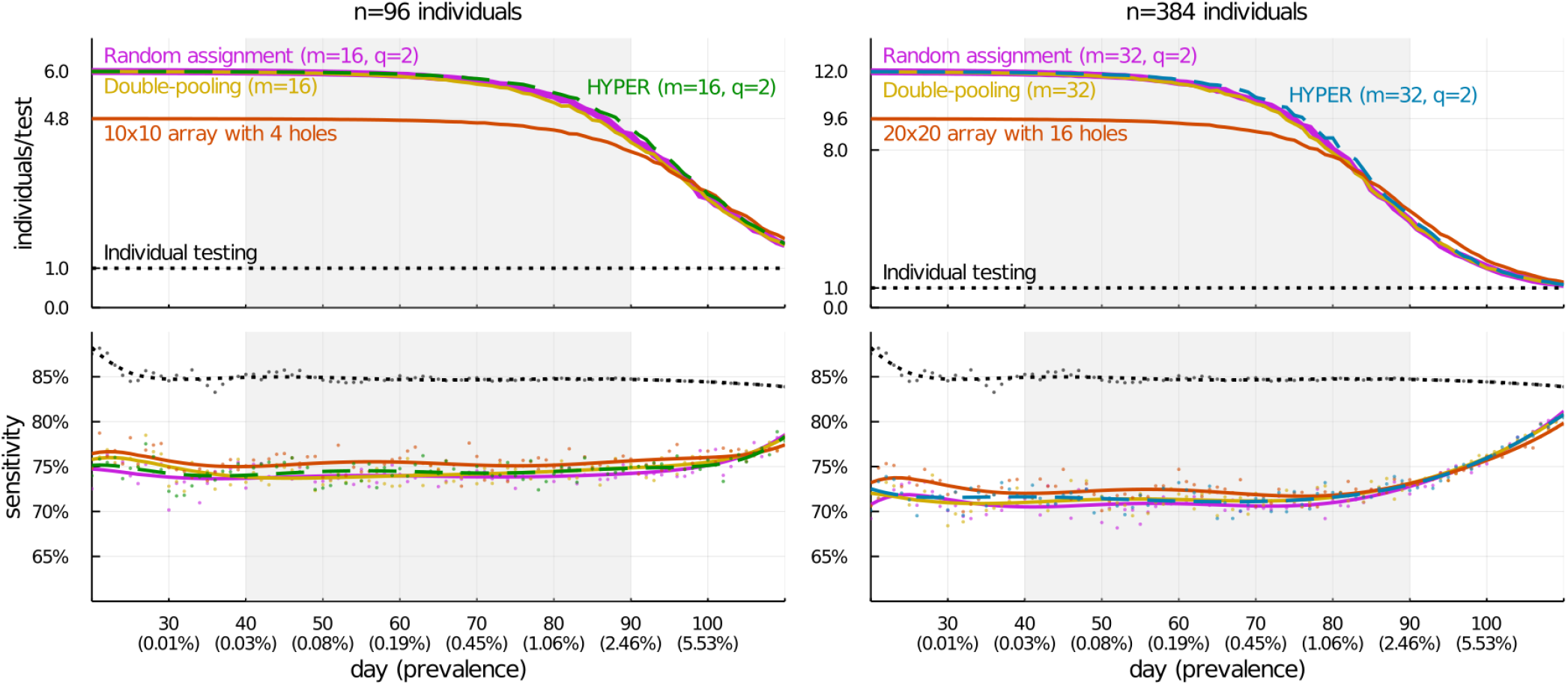
Comparison with additional designs. We consider the simulated epidemic from Fig. 2, and compare HYPER (as in Fig. 2a) with some additional methods: random assignment (each individual is assigned to *q* of *m* pools independently and uniformly at random), double-pooling (individuals are randomly partitioned into *m/q* pools *q* = 2 times), and balanced arrays with holes (pools are rows and columns of a square array where some number of cells along the diagonal are empty). As before, average values of efficiency (relative to individual testing) and sensitivity of the various pooling designs are shown for each day, with results averaged across 200,000 random trials. For sensitivity, raw averages are shown as dots with degree-8 polynomial fits overlaid as curves; the curves for efficiency depict raw averages. For both *n* = 96 and *n* = 384 individuals per batch, the average efficiency and sensitivity of both random assignment and double-pooling are generally similar to their corresponding HYPER designs. However, it turns out that these random designs behave less consistently than HYPER. Their performance depends on which individual happens to be positive, which is undesirable from a laboratory standpoint. This aspect is obscured by the averages in this figure, and we investigate it separately in Supplementary Fig. 7. Compared with the balanced array designs with holes, HYPER uses fewer pools and is roughly 25% more efficient for much of the 50-day window highlighted. HYPER also has correspondingly larger pool sizes, and is slightly less sensitive. Note that the 10 × 10 and 20 × 20 arrays here are, respectively, the smallest square arrays that can accommodate *n* = 96 and *n* = 384 individuals without placing multiple individuals in the same array cell. Forming balanced arrays with the same number of pools as HYPER (i.e., *m* = 8 and *m* = 16) requires extending the design to assign multiple individuals to some array cells, and we consider these designs in Supplementary Fig. 6.

**Supplementary Figure 6:**
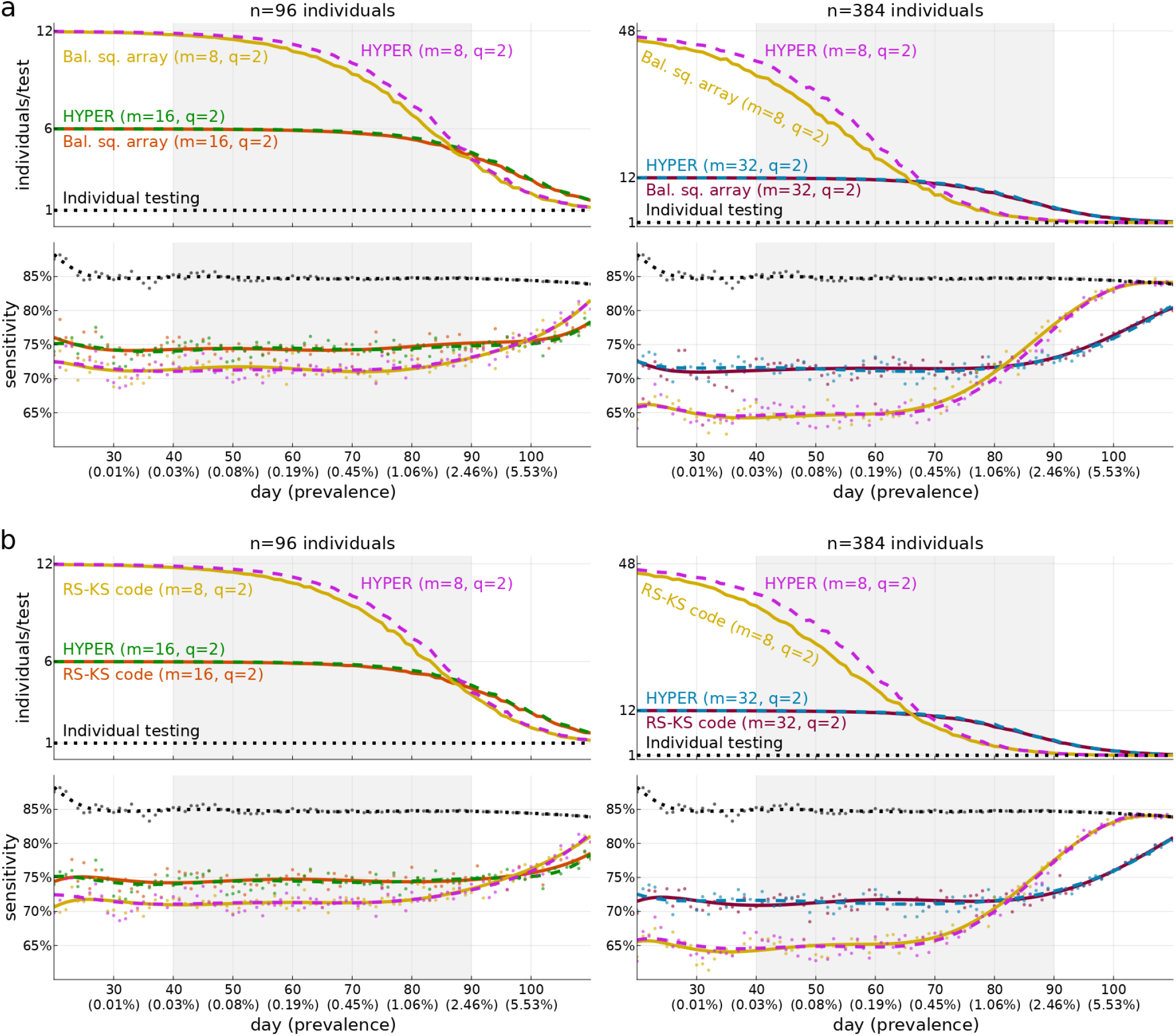
Comparison with balanced array and code-based designs with matching parameters. We repeat the comparison of Supplementary Fig. 5 but with balanced array (**a**) and Reed-Solomon Kautz-Singleton (RS-KS) code-based (**b**) designs using the same pooling parameters (number of individuals *n*, number of pools *m*, pools per individual *q*) as the HYPER designs. The two designs are described in the Supplementary Methods (balanced arrays designs on pg. S2, RS-KS code-based designs on pg. S3). For both designs, matching the pooling parameters requires recycling pool assignments in a similar way as is done in HYPER. Without doing so, both balanced square arrays and RS-KS designs with *q* = 2 can accommodate no more than (*m/*2)^2^ with *m* pools. The sensitivity of these methods closely matched those of HYPER when the pooling parameters were the same. Sensitivity depends significantly on the number of individuals per pool, which is *nq/m* for all three methods since they all have balanced pools. HYPER had either similar or better efficiency than both methods, with a larger improvement arising for more aggressive pooling parameters that used fewer pools and yielded greater efficiency at low prevalence.

**Supplementary Figure 7:**
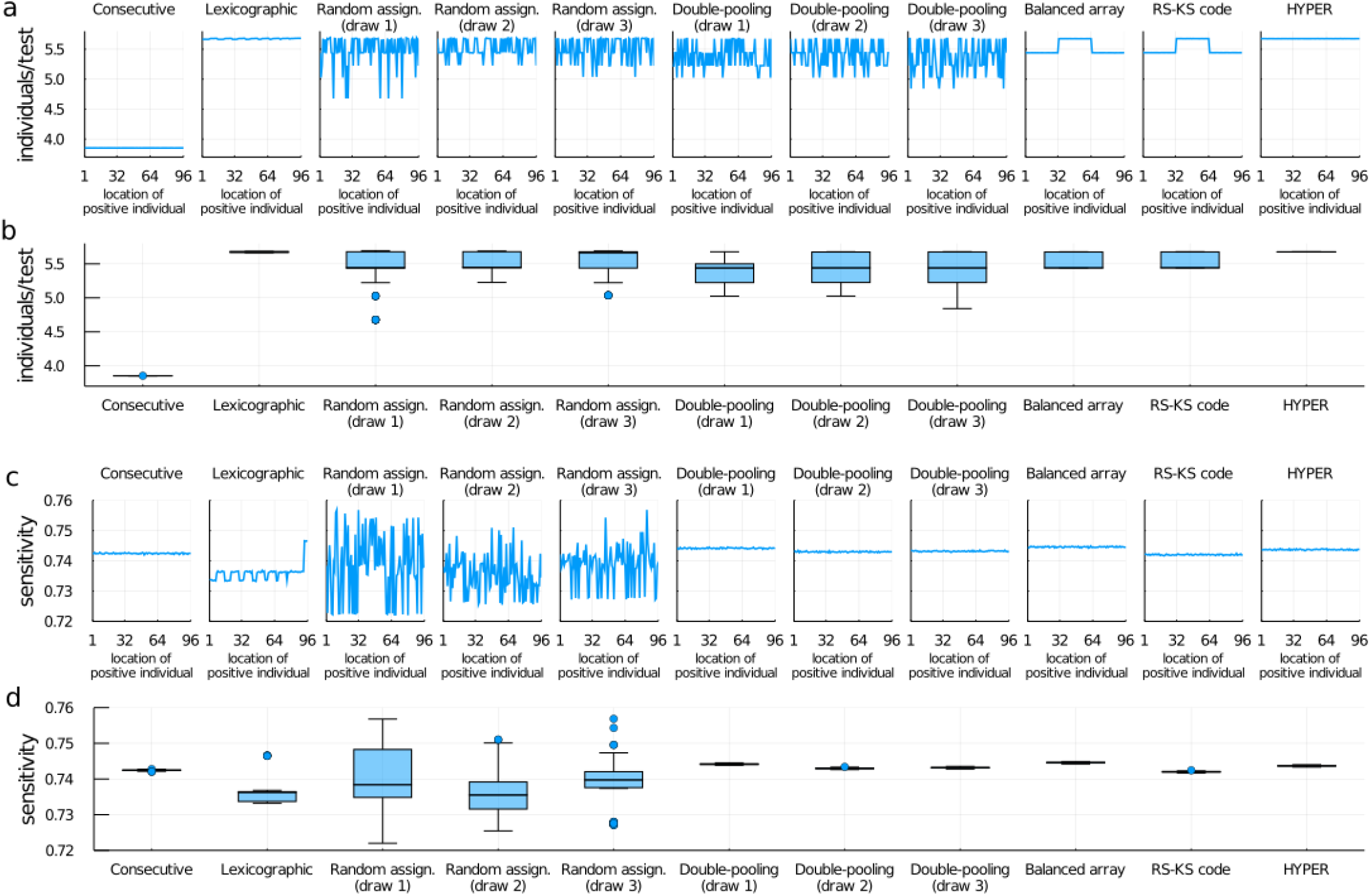
Impact of imbalanced designs. We investigate how balance affects how much efficiency and sensitivity vary depending on where positive individuals happen to fall. We consider designs with *n* = 96 individuals per batch, *m* = 16 pools and *q* = 2 splits, and we suppose exactly one individual is positive in each batch. That individual’s viral load is drawn from the distribution of nonzero viral loads on day 80 of the simulated epidemic from Fig. 2 (for which the prevalence is roughly 1.06%); everyone else has zero viral load. We consider placing the positive individual in each of the *n* slots of the batch (i.e., as individual 1, individual 2, etc.), and compare the average values (from 100,000 trials) for efficiency (**a**, relative to individual testing) and for sensitivity (**c**) as functions of the location. Consecutive pooling (AB, CD, EF,…), only uses a subset of the 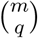 possible pool combinations and was generally less efficient. It does, however, have balanced pools yielding uniform sensitivity regardless of where the positive individual fell. Lexicographic pooling (AB, AC, AD, …) has balanced pool combinations yielding uniformly high efficiency. However, it has unbalanced pools resulting in uneven sensitivity. Random assignment pooling (each individual assigned to *q* of *m* pools uniformly at random) has varying performance depending on which particular design gets drawn, and often has unbalanced pool combinations and pools. As a result, all three draws had uneven efficiency and sensitivity. Double-pooling (individuals randomly partitioned into *m/q* pools *q* = 2 times), guarantees balanced pools but often has unbalanced pool combinations. As a result, all three draws had uniform sensitivity but uneven efficiency. Note that uneven efficiency means that the stage two workload is more unpredictable, which can make the logistics of large-scale screening more difficult. Balanced array pooling (pg. S2) and Reed-Solomon Kautz-Singleton (RS-KS; pg. S3) code-based pooling both had balanced pools here, and each was maximally balanced on the (*m/*2)^2^ pool combinations it used. However, neither was perfectly balanced on the pool combinations it used; in both cases, some pool combinations were used twice while others were used once. Both had uniform sensitivity and slightly uneven efficiency. In contrast, HYPER has both balanced pool combinations and balanced pools. As a result, it had both uniformly high efficiency *and* uniform sensitivity. Moreover, its median efficiency (5.68 individuals/test) and median sensitivity (74.4%) were generally among the best (**b**,**d**). The boxplots (**b**,**d**) each include a center line (median), box limits (upper and lower quartiles), whiskers (1.5x the interquartile range), and points (outliers).

**Supplementary Figure 8:**
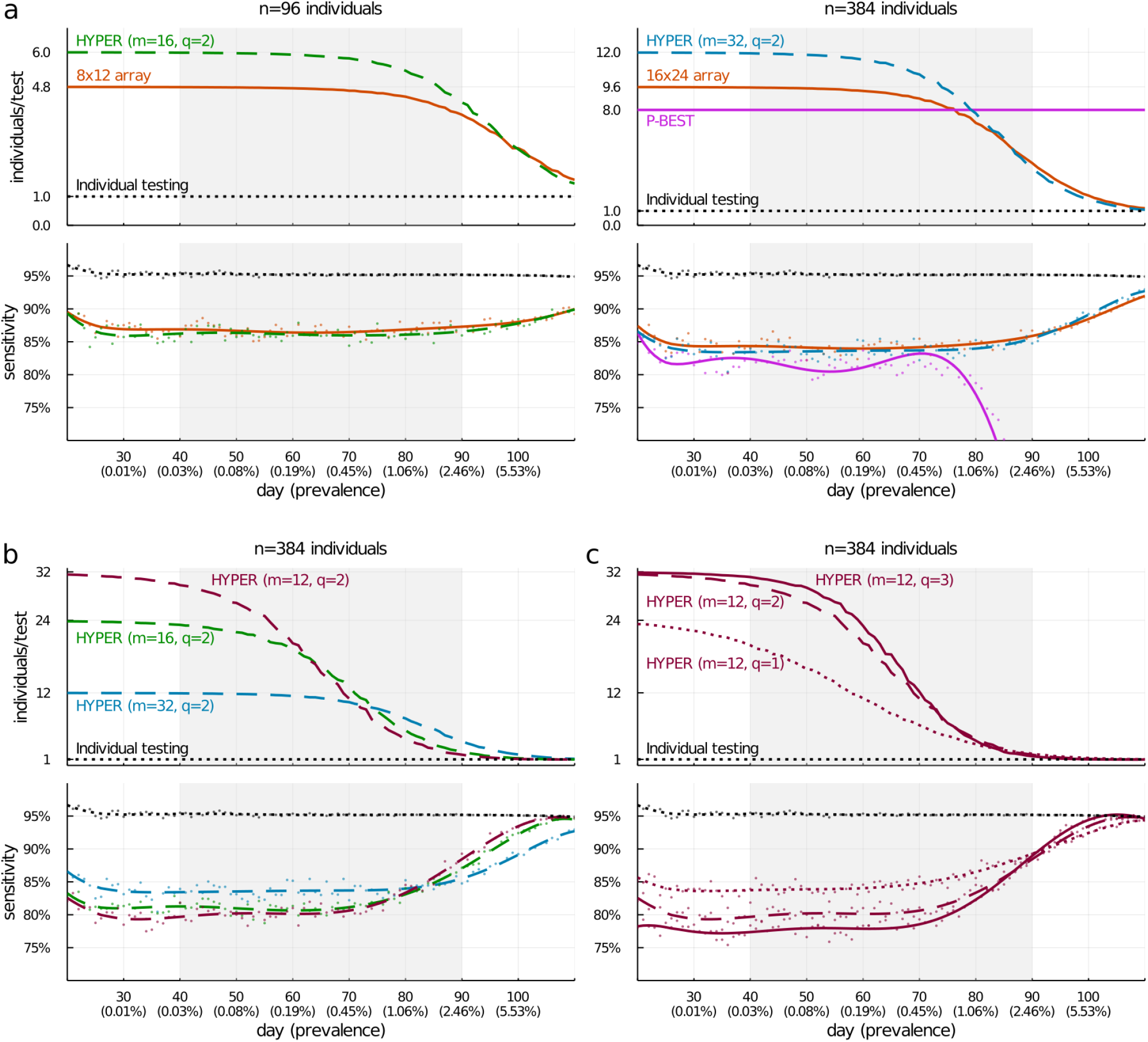
Efficiency and sensitivity of pooled testing during a simulated epidemic (Fig. 2) repeated for a 25-fold reduction in the limit of detection (from 100 to 4). With this reduction in the limit of detection (LOD), smaller viral loads are detected resulting in a higher individual testing sensitivity of roughly 95%. The group testing methods have a corresponding increase in sensitivity, with their relative sensitivity remaining similar to before. For example, the H_96,16,2_ design and the 8 × 12 array design have a 0.54 percentage point difference in sensitivity averaged across days 40-90 here, compared with a 0.24 percentage point difference before (Fig. 2). The group testing methods also have lower sensitivity than individual testing again due to the dilution of viral loads during the pooled testing of stage one. The gap is now slightly smaller; e.g., the sensitivity of the H_96,16,2_ design is roughly 9 percentage points lower here, compared with a roughly 10 percentage point difference before (Fig. 2). Efficiency for the various methods is very similar to before (Fig. 2), with HYPER enjoying essentially the same gains in efficiency over individual testing.

**Supplementary Figure 9:**
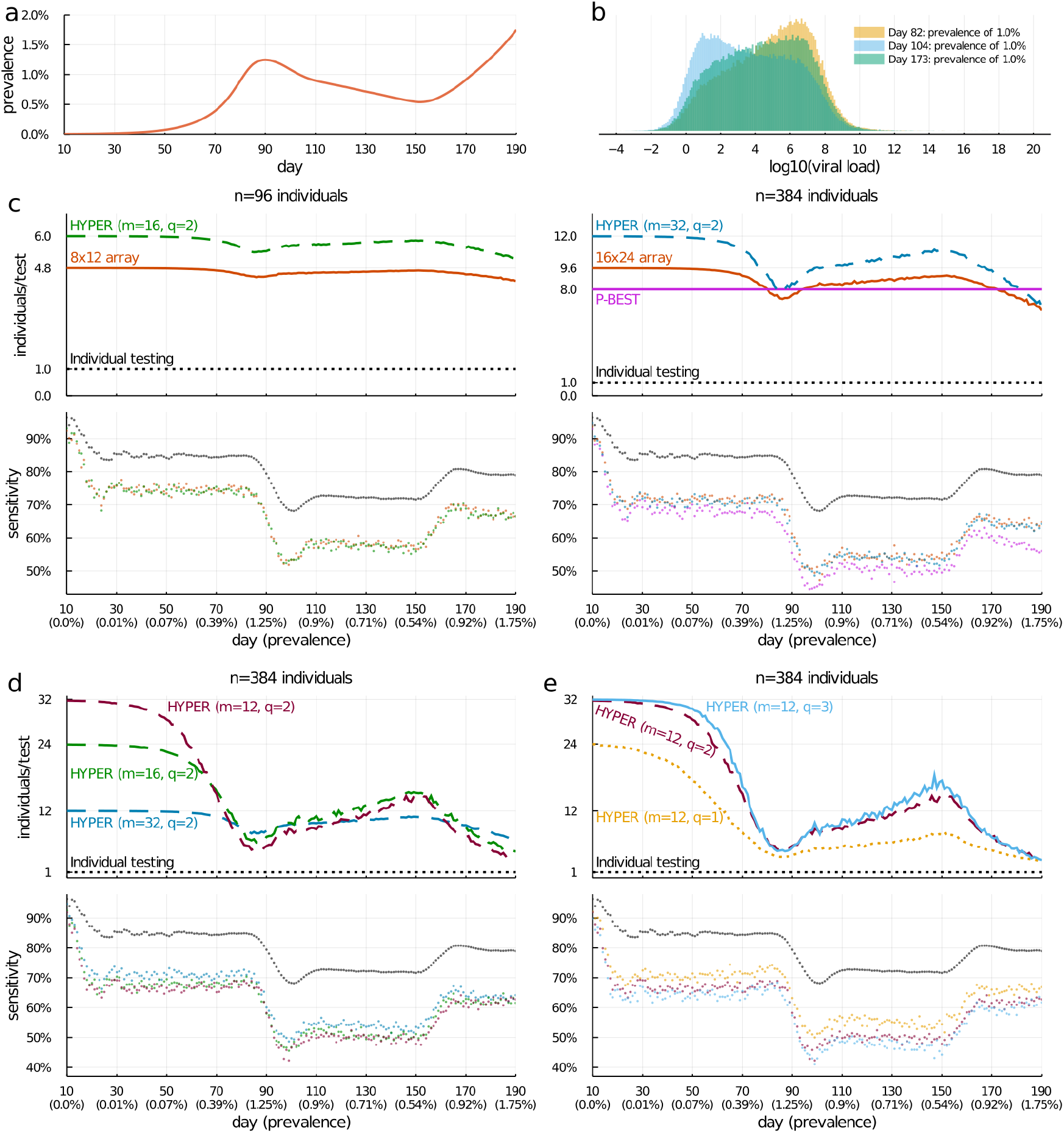
Efficiency and sensitivity of pooled testing during a simulated epidemic (Fig. 2) repeated with a two-wave epidemic. To evaluate a later (post-exponential) phase, we repeated our simulation but with a population undergoing a sustained, two-wave epidemic (obtained from Cleary and Hay et al.^11^). The simulated population was generated from an SEIR model with transmission rate modified at two time points: *R*_0_ was initiated at 2.5 at day 0, decreased to 0.8 at day 80, and subsequently increased to 1.5 at day 150. The result is an epidemic with an initial wave, followed by a decline phase and subsequently another growth phase (**a**). We compare the efficiency and sensitivity of the same methods as before (**c**,**d**,**e**). Consistent with earlier studies^11^, sensitivity is generally lower for all methods (including individual testing) during the decline phase compared to the two growth phases. For example, compare day 104 (during decline) with days 82 and 173 (during growth), which all had prevalence around 1.0%. The difference in sensitivity can be explained by observing that the distribution of nonzero viral loads (**b**) on day 104 is shifted to the left relative to days 82 and 173, due to a shift away from recent infections. Hence, positive viral loads are more likely to fall below the limit of detection and get missed. At the same time, the relative performance of the methods is similar to before. The chosen HYPER designs are as sensitive as their corresponding array designs, while being roughly 20-25% more efficient for *n* = 96 and roughly 10-25% more efficient for *n* = 384. For *n* = 384, the HYPER design is generally more efficient than P-BEST (up to 50% more) until day 180 while also being generally more sensitive. After day 180, P-BEST is more efficient but at an additional cost of sensitivity.

**Supplementary Figure 10:**
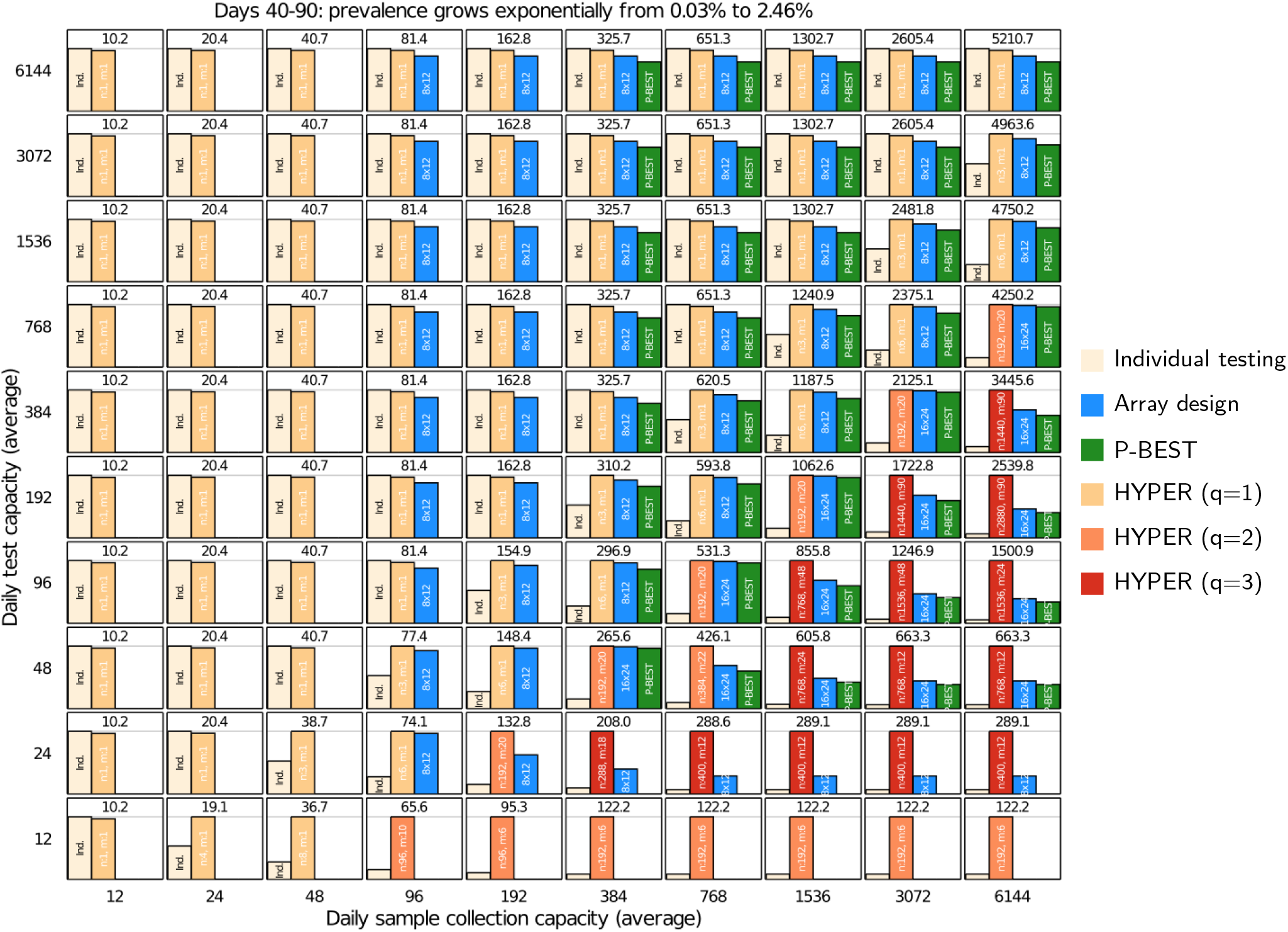
Detailed comparison corresponding to Fig. 3. Each cell shows the effective number of individuals screened given constrained sample collection and testing capacities, for individual testing, HYPER (best *n* and *m* noted in white with *q* indicated by bar color), plate-based array designs^9^ (better design between 8 × 12 and 16 × 24 noted in white), and P-BEST^8^. The best effective screening capacity for each cell is shown in black.

**Supplementary Figure 11:**
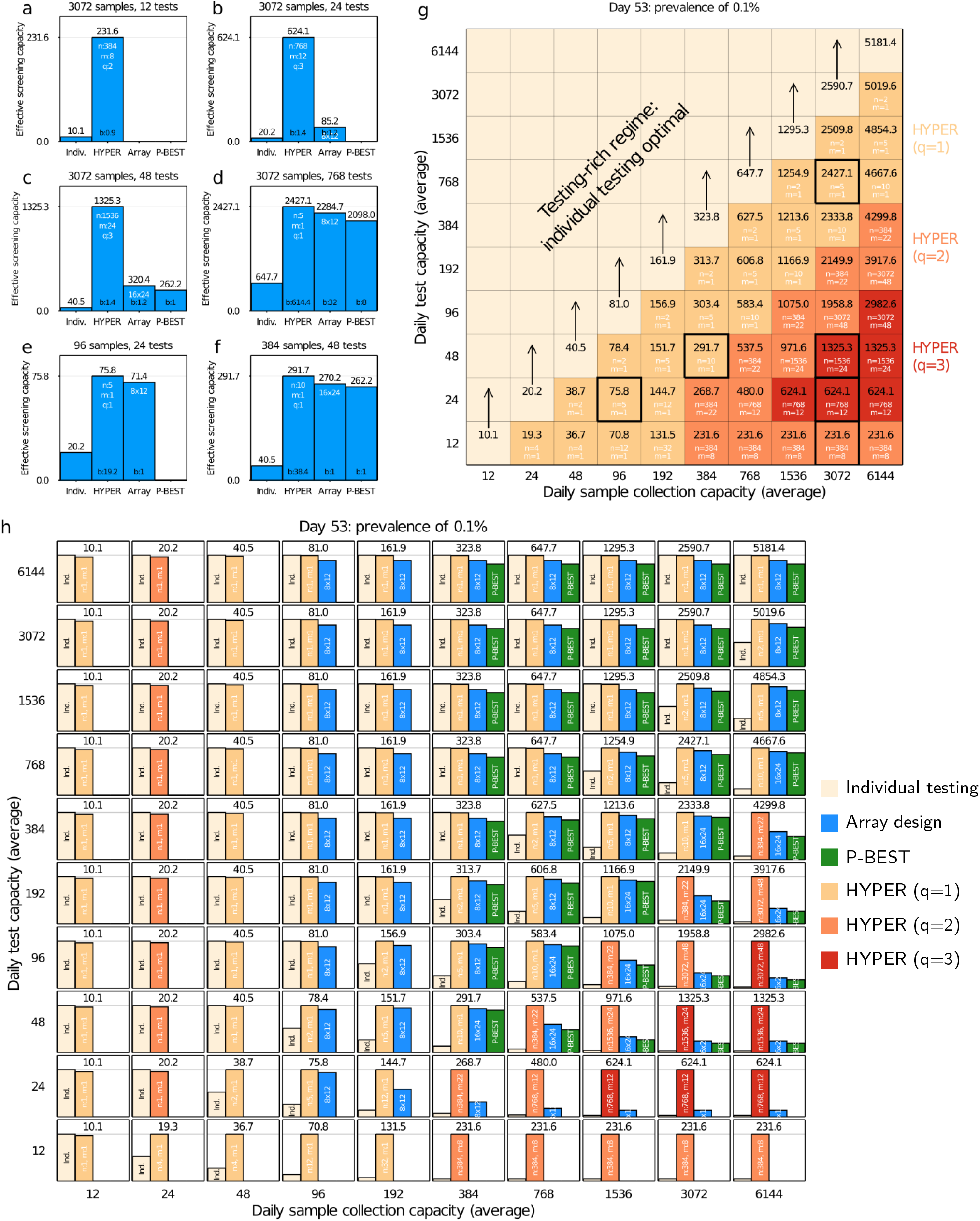
Comparison of pooling methods under resource constraints (Fig. 3 and Supplementary Fig. 10) repeated for day 53 (prevalence of 0.1%). At this low prevalence, HYPER is the most effective across the grid of resource constraints and can be significantly so. The low prevalence setting also favors simple designs (low *q*) across more of the test-constrained regime.

**Supplementary Figure 12:**
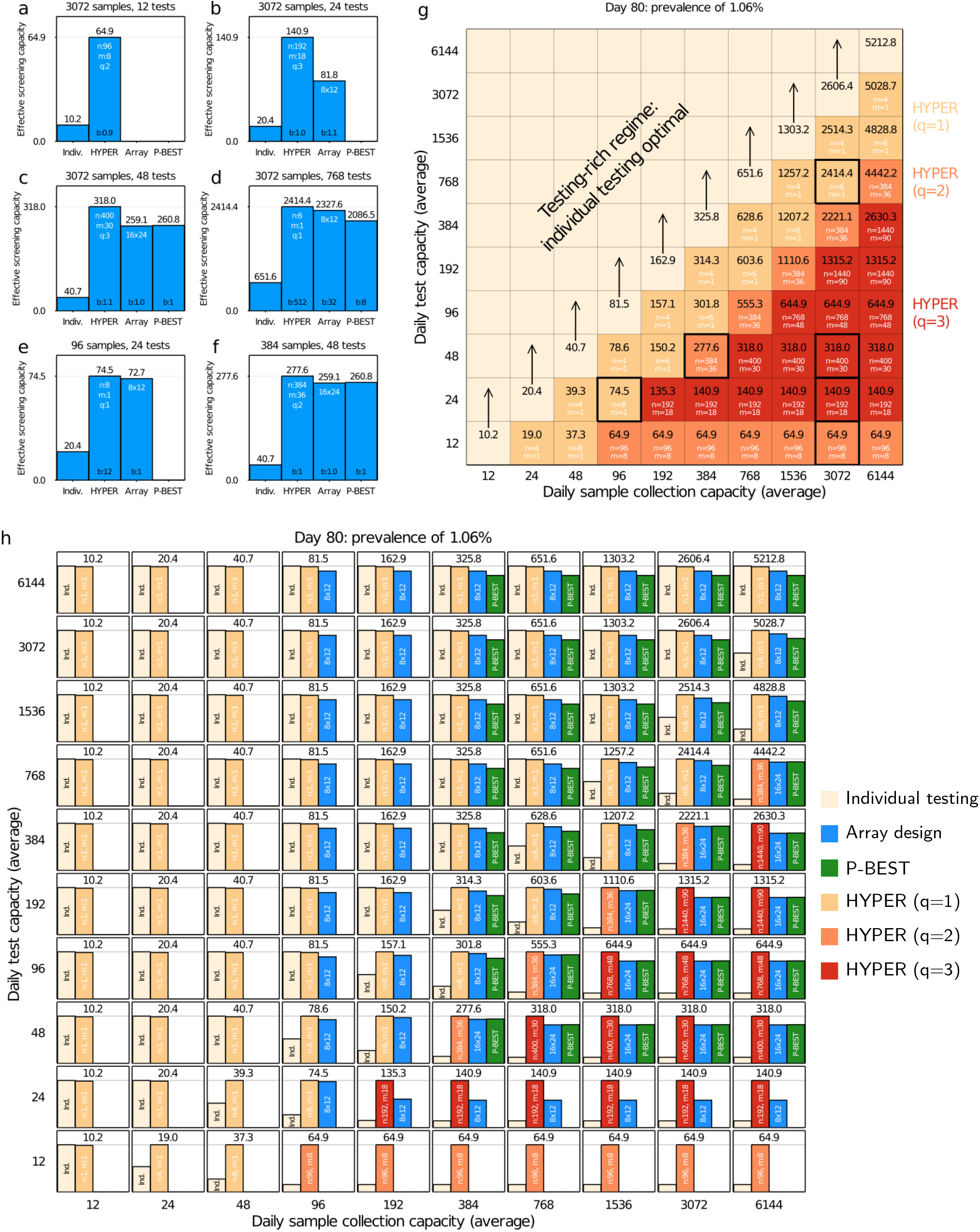
Comparison of pooling methods under resource constraints (Fig. 3 and Supplementary Fig. 10) repeated for day 80 (prevalence of 1.06%). At this moderate prevalence, HYPER remains best across all scenarios. Compared to Supplementary Fig. 11, which had lower prevalence, designs with higher *q* are more often the most effective.

**Supplementary Figure 13:**
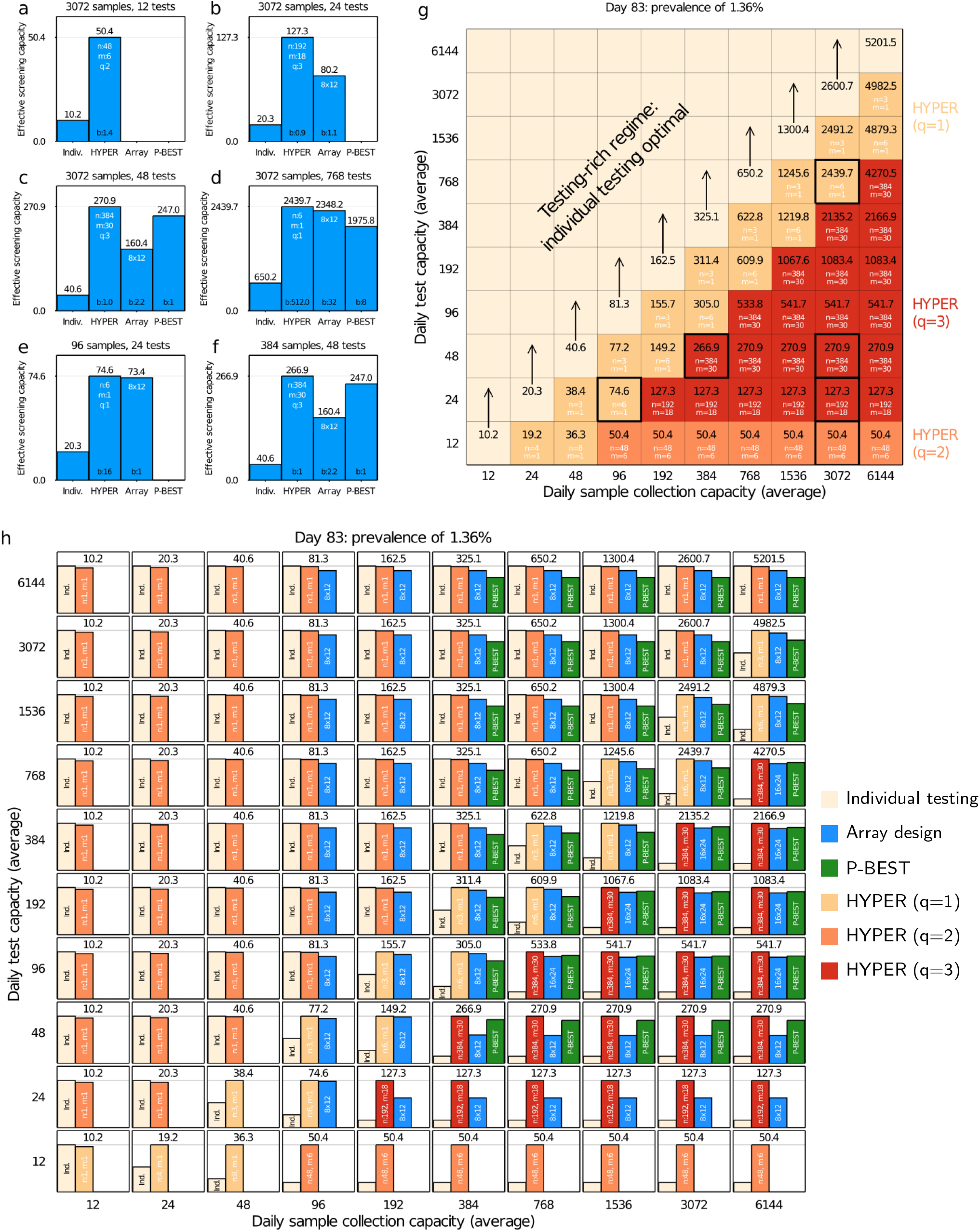
Comparison of pooling methods under resource constraints (Fig. 3 and Supplementary Fig. 10) repeated for day 83 (prevalence of 1.36%). At this prevalence, HYPER remains best across all scenarios. Compared to Supplementary Fig. 12, designs with higher *q* are even more often the most effective.

**Supplementary Figure 14:**
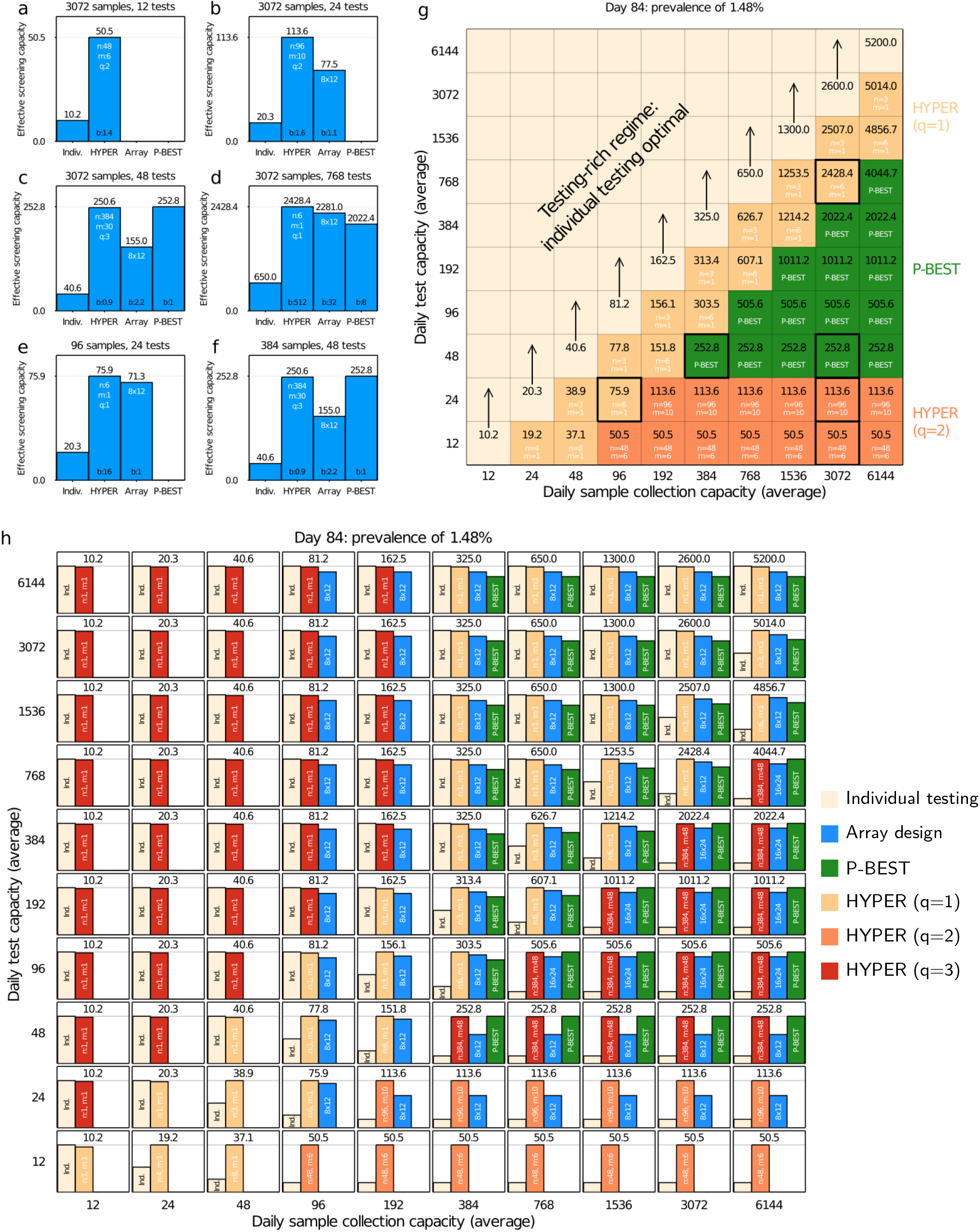
Comparison of pooling methods under resource constraints (Fig. 3 and Supplementary Fig. 10) repeated for day 84 (prevalence of 1.48%). At this intermediate prevalence, there is a subset of scenarios in which P-BEST outperforms HYPER. However, HYPER is still nearly as effective in these scenarios. Outside these settings P-BEST is either not viable or substantially under-performs HYPER.

**Supplementary Figure 15:**
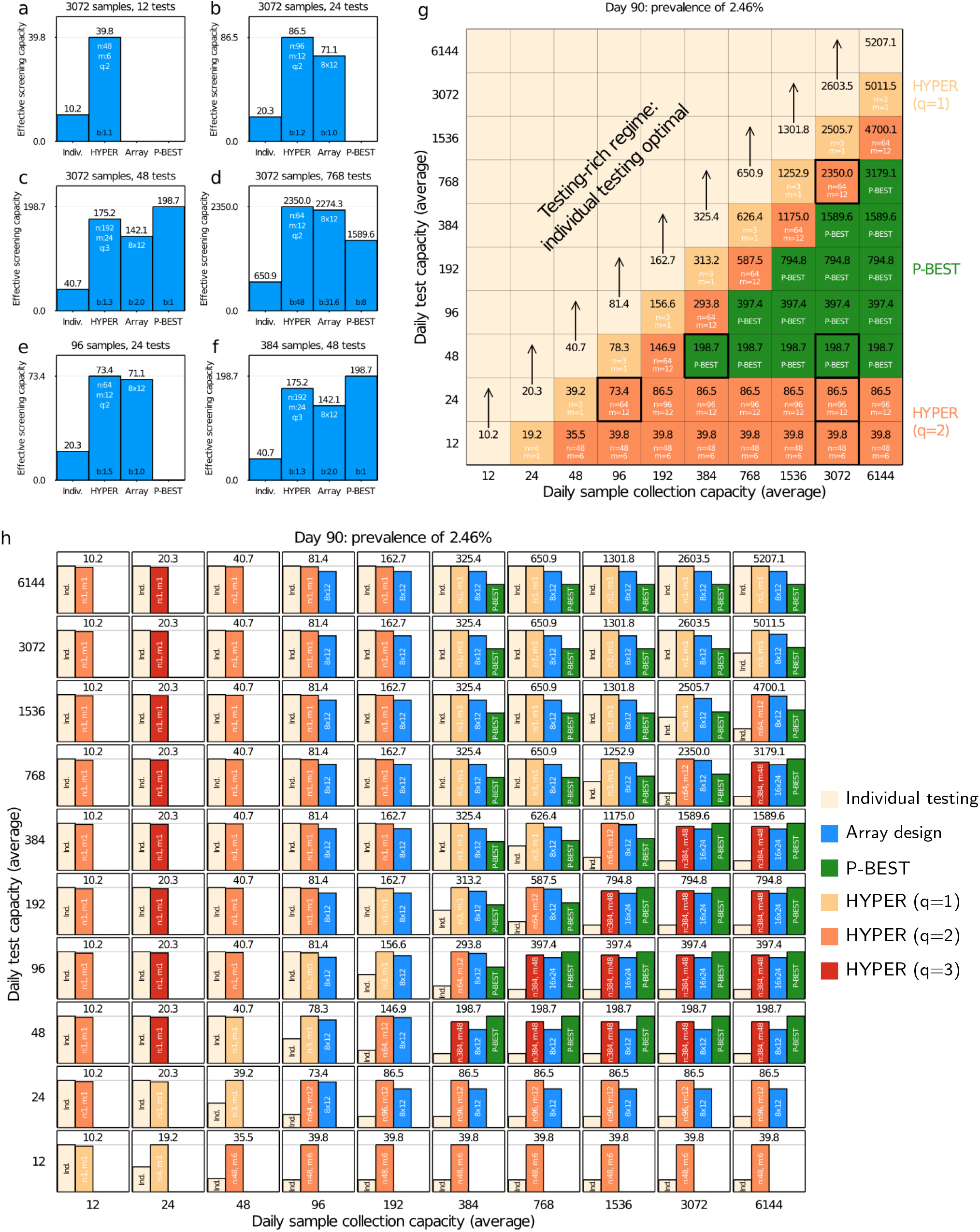
Comparison of pooling methods under resource constraints (Fig. 3 and Supplementary Fig. 10) repeated for day 90 (prevalence of 2.46%). At this prevalence, P-BEST continues to outperform HYPER on a subset of scenarios, with a larger gap than in Supplementary Fig. 14. Outside these settings P-BEST is still either not viable or substantially under-performs HYPER.

**Supplementary Figure 16:**
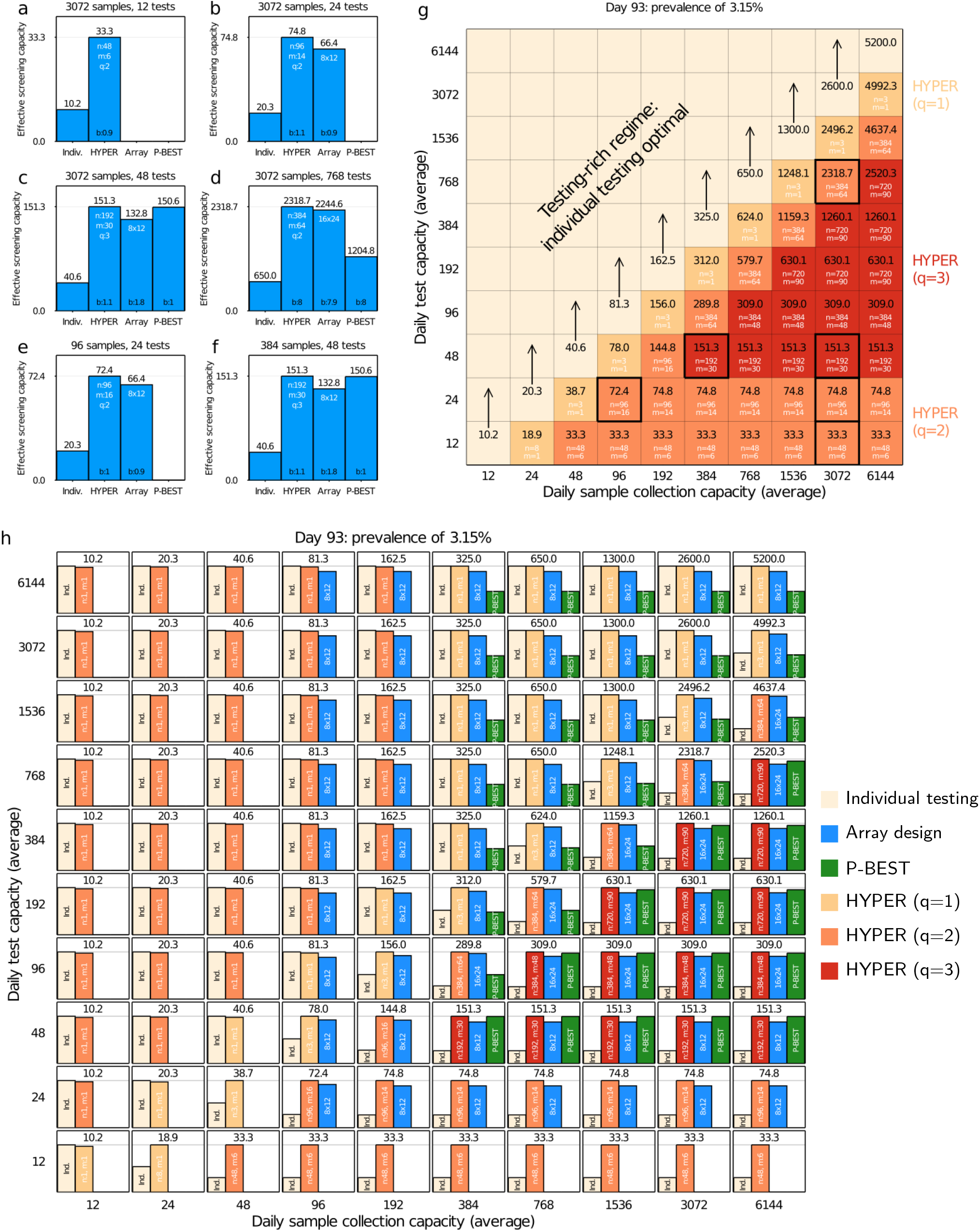
Comparison of pooling methods under resource constraints (Fig. 3 and Supplementary Fig. 10) repeated for day 93 (prevalence of 3.15%). At this higher prevalence, HYPER again performs best across all scenarios.

**Supplementary Figure 17:**
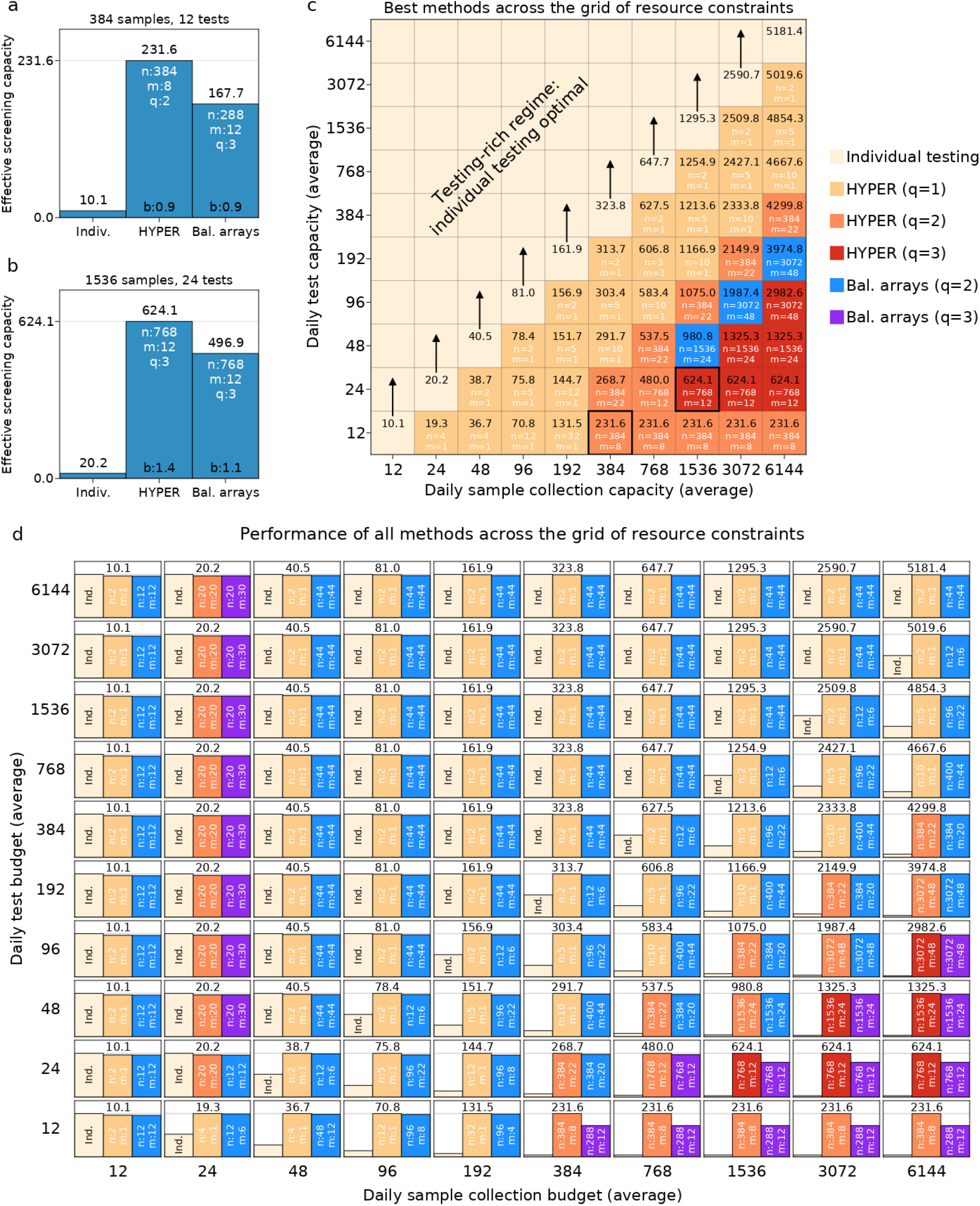
Comparison with balanced array designs under resource constraints for day 53 (prevalence of 0.1%). HYPER was about as effective as or substantially more effective than balanced array designs (Supplementary Methods, pg. S2) across the entire grid of resource constraints (**c, d**). Both methods were optimized over the same sweep of *n, m* and *q* (Supplementary Table 1). In important testing-constrained scenarios (with sample collection capacity significantly outstripping testing capacity), HYPER was up to 38% more effective than balanced arrays (e.g., for a budget of 384 samples and 12 tests; **a**). In some cases, HYPER was more effective because its greater efficiency enabled it to use more aggressive design parameters (**a**). In some other cases, the best design parameters for both methods was the same, but HYPER was more effective with the same parameters (e.g., for a budget of 1536 samples and 24 tests; **b**).

**Supplementary Figure 18:**
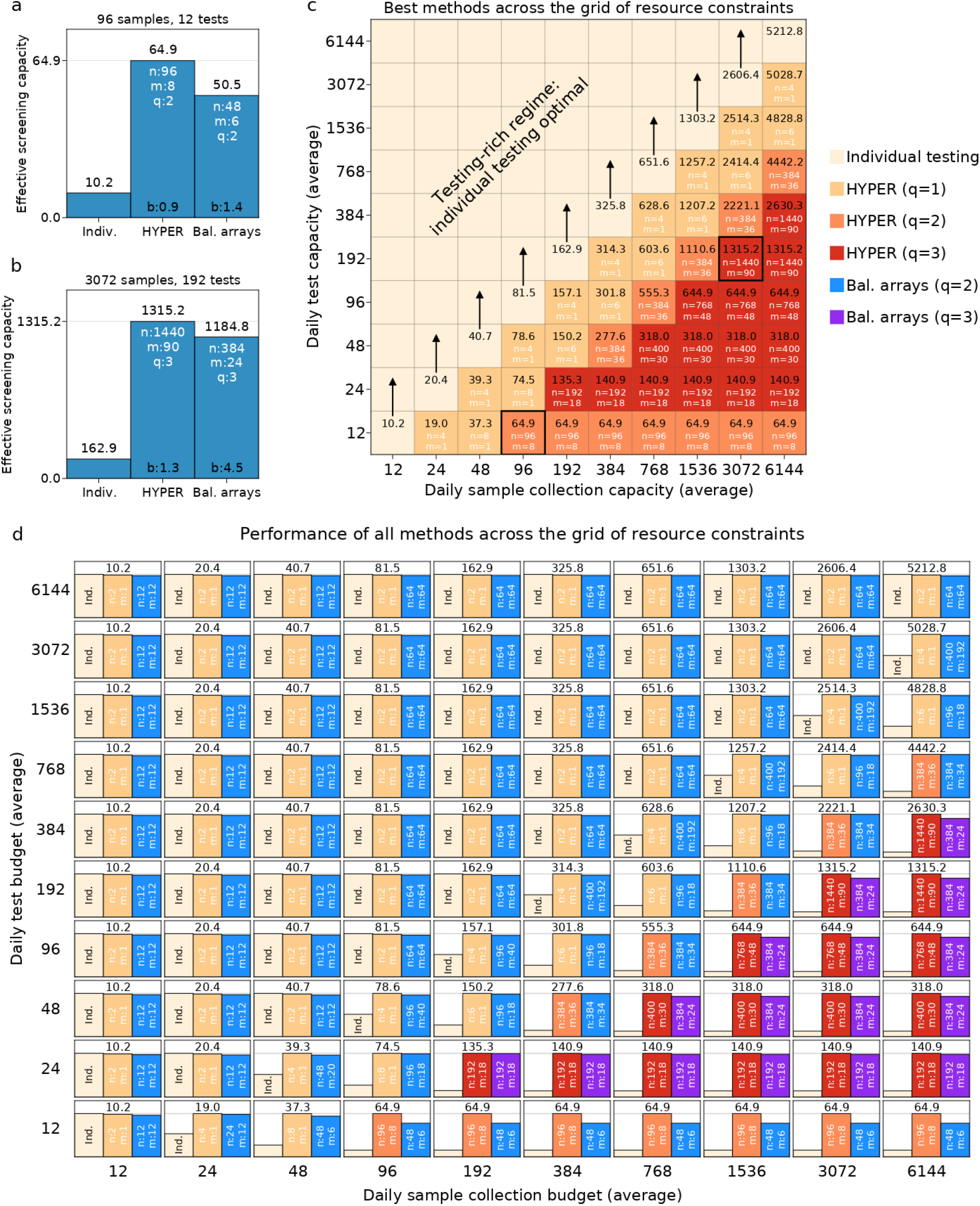
Comparison with balanced array designs under resource constraints for day 80 (prevalence of 1.06%). We repeated the comparison of Supplementary Fig. 17 for a later day with higher prevalence. As before, HYPER was more effective than balanced arrays across the entire grid of resource constraints (**c, d**), where both were optimized over the same set of design parameters (Supplementary Table 1). HYPER was also again significantly more effective than balanced arrays in important testing-constrained scenarios (with sample collection capacity significantly outstripping testing capacity). For example, HYPER was around 28% more effective than balanced arrays for a daily budget of 96 samples and 12 tests (**a**) and around 11% more effective for a daily budget of 3072 samples and 192 tests (**b**).

**Supplementary Figure 19:**
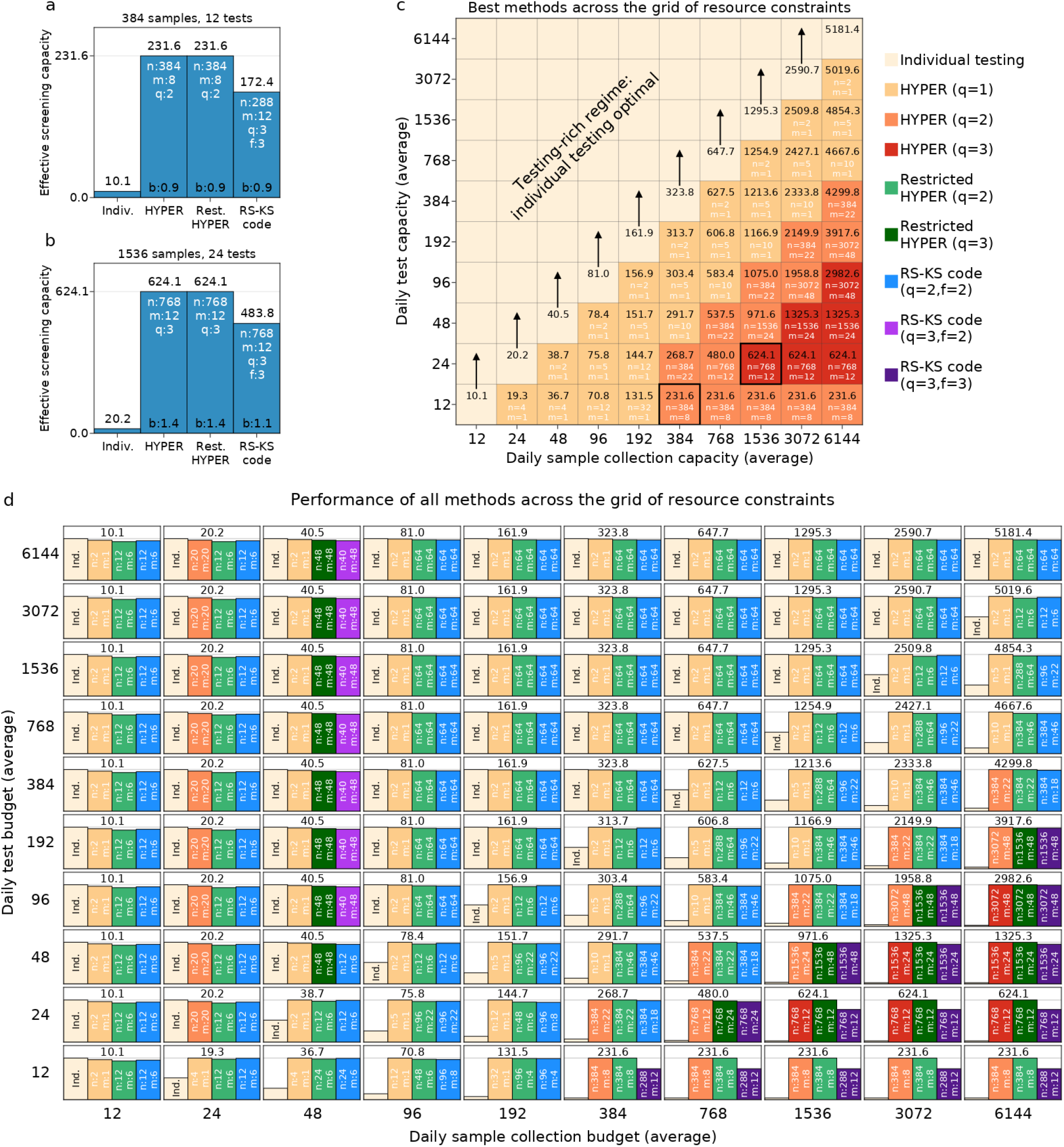
Comparison with Reed-Solomon Kautz-Singleton (RS-KS) code-based designs under resource constraints for day 53 (prevalence of 0.1%). HYPER was about as effective as or substantially more effective than RS-KS (Supplementary Methods, pg. S3) across the entire grid of resource constraints (**c, d**), even when restricted to only design parameters allowed by RS-KS (Supplementary Table 1 gives both the set used by HYPER and the restricted set used by Restricted HYPER and RS-KS). In important testing-constrained scenarios (with sample collection capacity significantly outstripping testing capacity), both HYPER and Restricted HYPER were up to 34% more effective than RS-KS (e.g., for a budget of 384 samples and 12 tests; **a**). In some cases, they were more effective because the greater efficiency of HYPER enabled them to use more aggressive design parameters (**a**). In some other cases, the best design parameters for all methods was the same, but HYPER and Restricted HYPER were more effective with the same parameters (e.g., for a budget of 1536 samples and 24 tests; **b**).

**Supplementary Figure 20:**
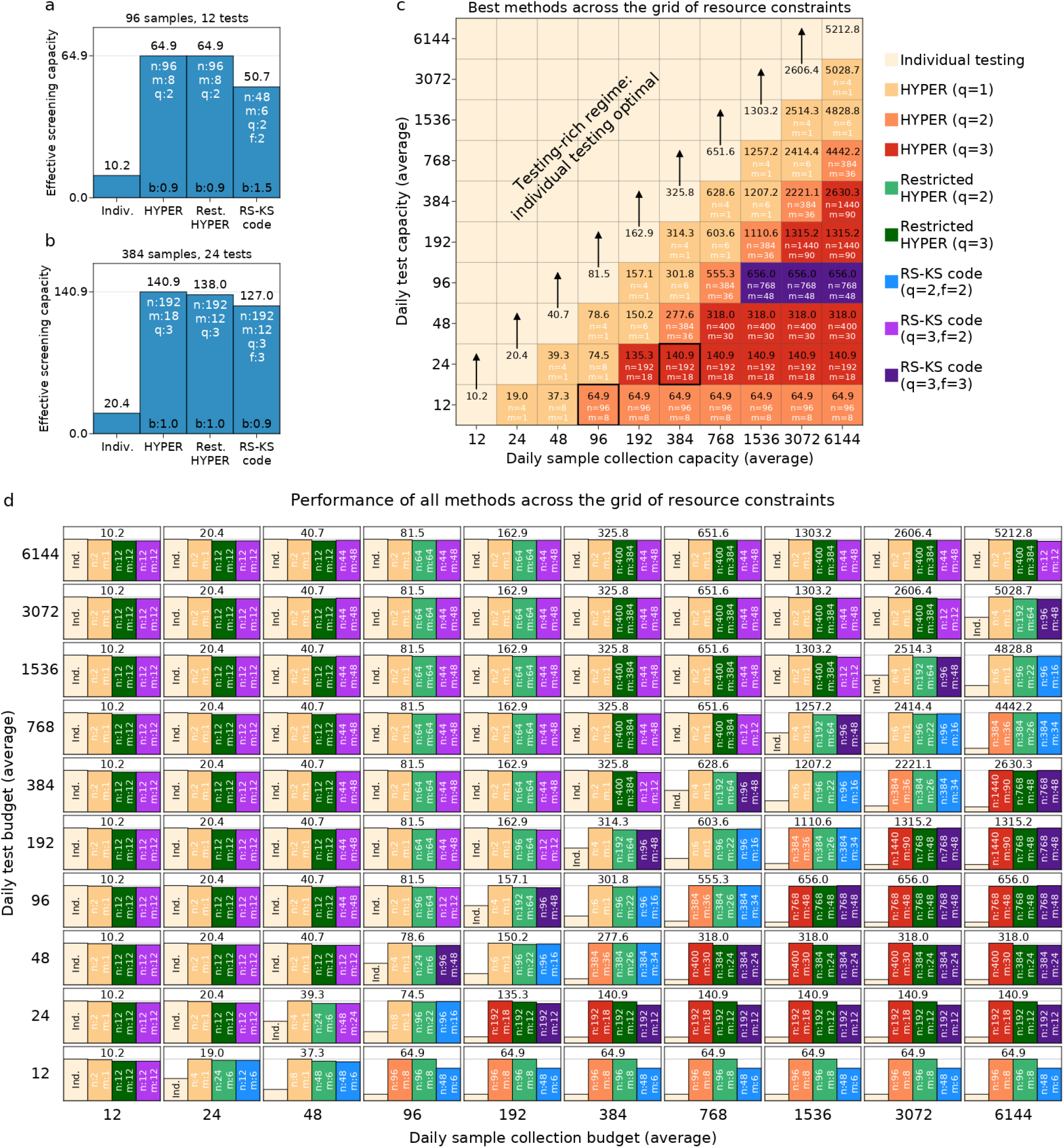
Comparison with Reed-Solomon Kautz-Singleton (RS-KS) code-based designs under resource constraints for day 80 (prevalence of 1.06%). We repeated the comparison of Supplementary Fig. 19 for a later day with higher prevalence. As before, HYPER and Restricted HYPER were more effective than RS-KS across the entire grid of resource constraints (**c, d**), where Restricted HYPER was optimized over the same set of design parameters as RS-KS (Supplementary Table 1). HYPER and Restricted HYPER were also again significantly more effective than RS-KS in important testing-constrained scenarios (with sample collection capacity significantly outstripping testing capacity). For example, both were around 28% more effective for a daily budget of 96 samples and 12 tests (**a**) and over 8% more effective for a daily budget of 384 samples and 24 tests (**b**).

**Supplementary Table 1:**
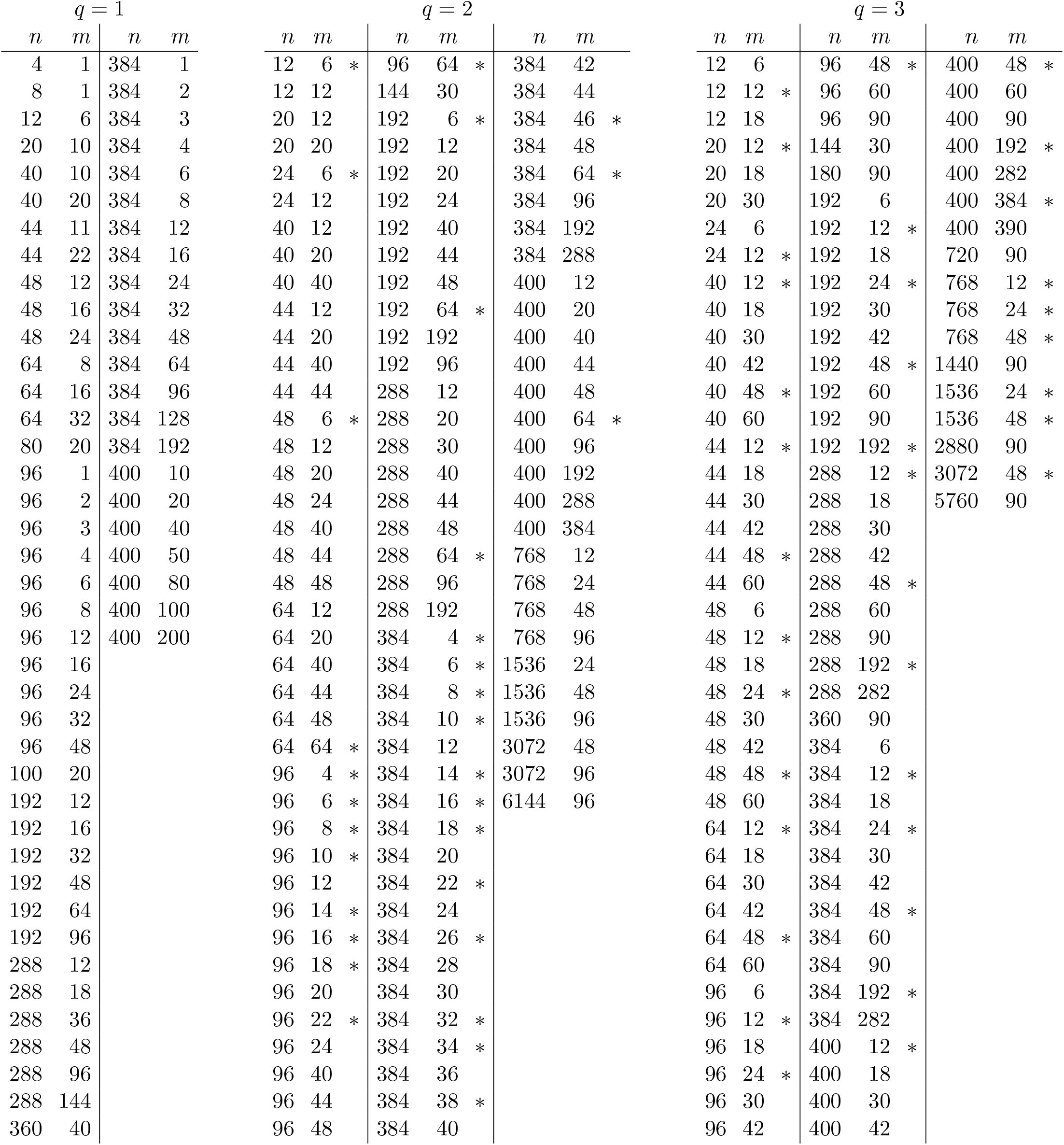
List of HYPER design H_*n,m,q*_ considered. We simulated the following HYPER configurations and selected the most effective for each scenario of the constrained-resource analyses shown in Fig. 3 and Supplementary Figs. 10 to 16. The same parameters were used in Supplementary Figs. 17 and 18, as well as Supplementary Figs. 19 and 20. Asterisks (*) denote the restricted set of parameters that are allowable for Reed-Solomon Kautz-Singleton (RS-KS) code-based designs (described in Supplementary Methods, pg. S3).

